# RD-Embed: Unified representations of rare-disease knowledge from clinical records

**DOI:** 10.64898/2026.04.02.26350083

**Authors:** Tudor Groza, Freddie Tan, Noah Tian Run Lim, Maheshwaran Windersalam Shanmugasundar, Jhanvi Kappaganthu, Jane Andrea Lieviant, Neerja Karnani, Haichao Chen, Tien Y Wong, Saumya Shekhar Jamuar

## Abstract

Rare diseases often present with incomplete, evolving symptoms and signs scattered across clinical notes and coded records, making diagnosis and gene discovery difficult even when genomic data are available. Existing approaches either depend on curated phenotype profiles or use general biomedical language models that are not aligned to rare-disease knowledge, limiting performance in early or ambiguous clinical presentations. Here, we show that RD-Embed – a three-stage representation framework that builds a base space that preserves domain knowledge, aligns clinical text and SNOMED-derived signals, and refines relationships with graph-based learning – enables robust rare-disease retrieval from heterogeneous clinical records. Across ten rare-disease datasets, RD-Embed attains up to >50% top-ten diagnostic retrieval using combined text and phenotype features, compared with ∼30% on average for other embedding models and similarly sized large language models. On an EHR stress test, clinical alignment substantially improves text-based retrieval compared with ontology-only representations, supporting use in routine EHR data. We suggest RD-Embed is lightweight model that can be incorporated into existing hospital systems that supports rare disease identification and diagnosis, and gene prioritization.

## Introduction

Rare diseases affect more than 300 million individuals worldwide, yet many patients experience a “diagnostic odyssey” characterized by years of uncertainty, repeated investigations, and misdiagnoses before receiving a definitive answer. Clinical presentations are often heterogeneous, age-dependent, and incomplete – particularly in mitochondrial disorders, neurodevelopmental syndromes, and connective tissue diseases – so clinicians must reason over sparse, fragmented symptoms and signs and clinical information that evolves over time, frequently without a clear genotype-phenotype match even in the era of widespread genomic sequencing [1, 2]. Although large-scale sequencing has transformed rare disease medicine, 40-60% of patients undergoing clinical exome or genome sequencing remain undiagnosed, reflecting persistent limitations in gene variant interpretation when phenotypic knowledge is partial, inconsistently captured, or missing altogether.

A central obstacle is that rare-disease information in hospital systems is simultaneously structured and unstructured. Resources such as the Human Phenotype Ontology (HPO) [3, 4] and curated databases including OMIM [5], Orphanet [6], and Monarch [7] have standardized disease and phenotype concepts and enabled computational phenotyping. Yet real-world electronic health records (EHR) are captured and arrives to the clinicians as a mix of fragments in frequently incomplete free-text notes, partial phenotype descriptions, and inconsistent disease coding. Mapping narratives to standardized phenotype terms remains labour-intensive, error-prone, and incomplete, limiting the practical impact of ontology-based approaches in routine care [8, 9].

To address these challenges, phenotype-driven tools – such as Phenomizer [10], LIRICAL [11], Phen2Gene [12], and Exomiser/Genomiser [13, 14] – rank candidate diseases or genes from explicit phenotype profiles but also rely strictly on structured HPO-described representations. They therefore degrade when patient descriptions are imprecise or expressed only in narrative form. Even recent AI systems [15, 16] that extract phenotypes from EHR inherit some of these constraints, remaining dependent on predefined phenotype vectors and manually engineered scoring rules [17]. Clinicians are thus left balancing structured tools requiring clean inputs against workflows where the most informative signals often reside in text, codes, and partially observed features.

In parallel, large language models (LLMs) [18–25] have shown strong performance on medical text tasks including summarization, concept recognition, and information retrieval. Yet for rare disease diagnosis, these conditions are underrepresented in current training data of LLMs, terminology is specialized, and reasoning depends on subtle phenotype relationships not explicitly grounded in rare-disease knowledge structures. Consequently, LLMs have performed inconsistently when retrieving plausible diagnoses from sparse phenotypes or notes, with recent evaluations showing only modest diagnostic accuracy even for advanced foundation models [26, 27]. These limitations reveal a critical gap: we lack representations that unify structured ontologies with unstructured clinical text and codes for rare diseases, in a way that is robust to missing data, supports gene-centric interpretation, and can plug into (rather than compete with) modern LLM-based clinical AI. Fig 1 summarizes this workflow: structured signals (HPO terms, SNOMED concepts) and unstructured evidence (clinical notes, case reports) are mapped into a shared space that supports clinician-facing inference tasks and embedding-based integration with LLMs.

**Figure 1.**
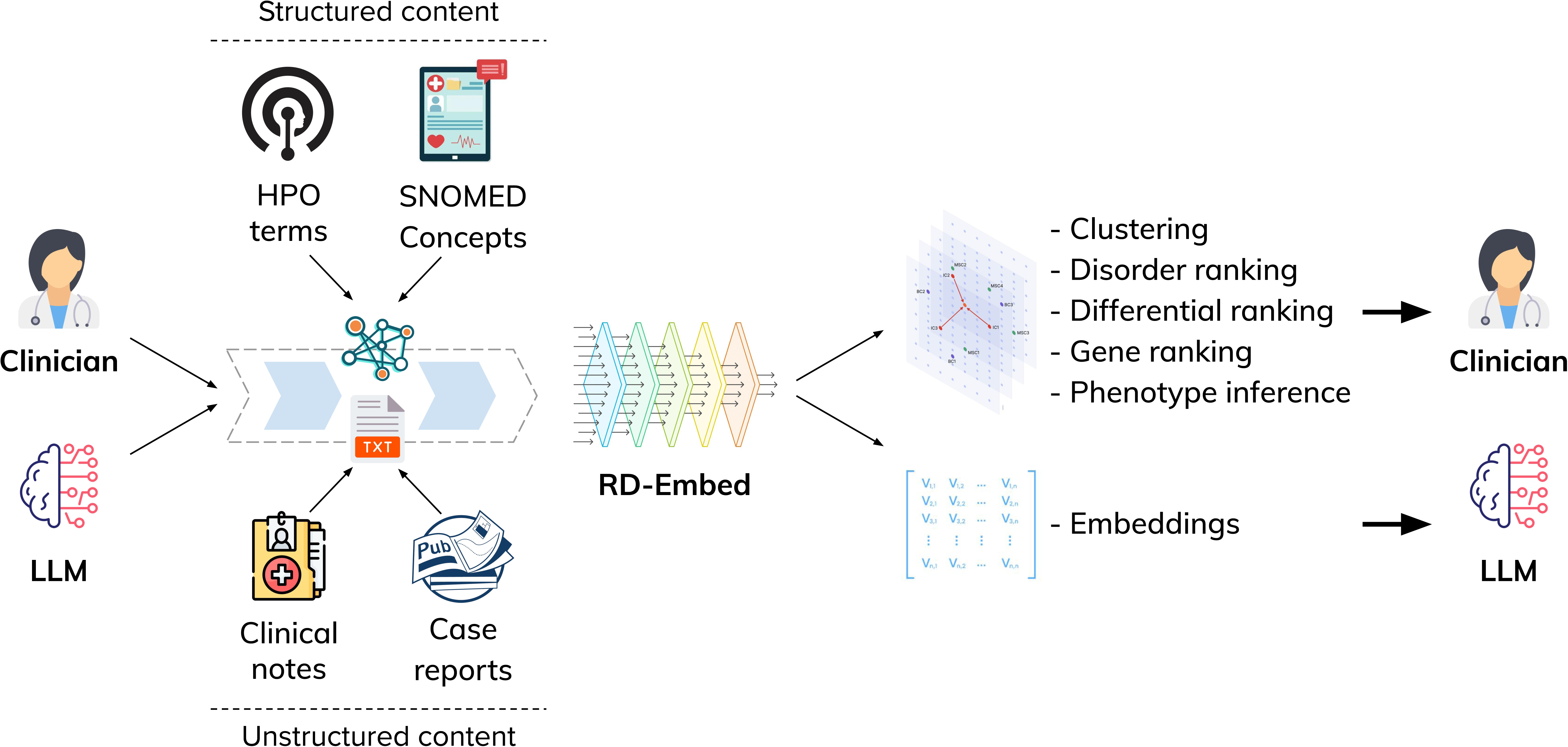
RD-Embed workflow for rare-disease retrieval and LLM grounding. Structured signals (HPO terms and SNOMED concepts) and unstructured evidence (clinical notes and published case reports) are encoded into a shared representation space by RD-Embed. The resulting embeddings support clinician-facing tasks – including disorder ranking, differential ranking, gene ranking, phenotype inference, and clustering – and can also be provided as embedding-based inputs to large language model (LLM) systems to ground downstream summarization and reasoning in rare-disease knowledge.

To address this gap, we introduce RD-Embed, a unified representation framework for rare disease diagnosis and clinical decision support. RD-Embed learns shared numerical representations of diseases, genes, phenotypes, and clinical descriptions by jointly modelling ontology structure, curated rare-disease knowledge, and real-world clinical data. The framework follows a three-stage design – a base space preserving domain knowledge, a clinical alignment bridge, and a graph-based refinement – ensuring that representations remain faithful to curated rare-disease structure while adapting to how phenotypes and diagnoses appear in notes and coded EHR data. Rather than relying on exact phenotype matches, RD-Embed captures graded clinical similarity, enabling comparison between incomplete patient presentations and known rare disease patterns.

Our contributions are threefold. (1) We develop a rare-disease-focused representation space integrating ontological phenotype structure with disease-gene-phenotype knowledge, enabling coherent similarity comparisons across entities. (2) We align this space to clinical reality by learning from free-text descriptions and coded EHR signals, allowing retrieval and ranking even when phenotypes are missing, noisy, or partially observed. (3) We provide a practical clinical AI interface: RD-Embed supports differential diagnosis ranking, gene prioritization, phenotype inference, and case-based retrieval, and can also serve as structured input to LLMs for context-aware reasoning without retraining on rare-disease-specific data. Across evaluations mirroring clinical workflows – retrieving plausible differentials, operating on EHR data, and supporting genomic interpretation – RD-Embed demonstrates consistent improvements over strong embedding and phenotype-based baselines, supporting scalable and actionable rare disease identification, diagnosis for clinicians in hospital systems.

## Results

### RD-Embed workflow and evaluation map

RD-Embed is engineered to solve a clinically constrained input problem: rare disease reasoning must work when evidence is incomplete, inconsistently coded, or available only as narrative text. Using RD-Embed’s stage-wise training (1. ontology preservation → 2. clinical alignment bridge → 3. knowledge graph refinement), we represent diseases, genes, phenotypes, and clinical descriptions in a shared embedding space so that similarity search and ranking can be performed across the same interface regardless of input type.

We evaluated RD-Embed with tasks aligned to common workflow questions: (i) **retrieving candidate diagnoses** across multiple rare-disease corpora, (ii) **scaling retrieval to EHR-style data** and SNOMED-coded histories, (iii) **prioritizing candidate genes** to support genomic interpretation, and (iv) **re-ranking differentials** to refine an initial shortlist into a clinically actionable ordering. We additionally compared RD-Embed to two large language models on difficult cohorts to contextualize performance against general-purpose clinical AI.

To mirror real-world intake variability, we tested three input settings: narrative notes or narrative-style descriptions of phenotypes (TEXT), structured phenotypes from HPO (and, in EHR analyses, SNOMED) (HPO | SNOMED), and both together (COMBINED). The test corpora span seven publicly available curated benchmarks (emerged from the rare disease community), HPO-only registry profiles (DECIPHER, Care4Rare), an EHR-derived rare-disease cohort from longitudinal SNOMED histories, and a text-only clinical genetics referral cohort (UDPS), covering inputs from specialist documentation to early referral and routine care (Table 1; Methods).

**Table 1.** Overall statistics of the corpora used for training and evaluation in RD-Embed. The training corpus, consisting of case reports, was compiled and verified by the authors. Six evaluation corpora were adopted from [15] and one from [40] – to provide results comparable to existing approaches. Two corpora (Care4Rare and DECIPHER) were provided by the organizations maintaining the underpinning registries following a data access request process. Two corpora were compiled from the Singapore Health System EHR – one focused on rare diseases, from the Undiagnosed Diseases Program Singapore and one filtered from a population-scale corpus of structured EHR data introduced in [41].

| Dataset | Case reports | Care4Rare | DECIPHER | HMS | LIRICAL | MME | PUMCH-ADM | PUMCH-L | PXF | RAMEDIS | UDPS | EHR |
| --- | --- | --- | --- | --- | --- | --- | --- | --- | --- | --- | --- | --- |
| Usage | Train | Eval | Eval | Eval | Eval | Eval | Eval | Eval | Eval | Eval | Eval | Eval |
| Nature | Pubs | Registry | Registry | Case data | Case reports | Case data | Case data | Case data | Case reports | Case data | EHR Clinical notes | EHR structured data |
| Modalities | TEXT, HPO | HPO | HPO | HPO | HPO | HPO | HPO | HPO | HPO | HPO | TEXT | SNOMED |
| Source | Pubmed | Care4Rare Canada | [39] | [15] | [15] | [15] | [15] | [15] | [40] | [15] | Local | [41] |
| # cases | 26,293 | 237 | 4,164 | 88 | 315 | 36 | 74 | 929 | 6,974 | 329 | 64 | 8,963 |
| # diseases | 4,410 | 376 | 1,360 | 67 | 221 | 16 | 32 | 158 | 519 | 65 | 43 | 634 |
| # genes | 2,992 | 203 | 967 | 49 | 232 | 15 | 70 | 226 | 433 | 66 | 48 | 1,217 |
| Cases per disease |  |  |  |  |  |  |  |  |  |  |  |  |
| Min | 1 | 1 | 1 | 1 | 1 | 1 | 3 | 1 | 1 | 1 | 1 | 1 |
| Median | 7 | 1 | 2 | 2 | 1 | 1 | 5 | 2 | 5 | 2 | 1 | 3 |
| Max | 59 | 14 | 244 | 11 | 18 | 10 | 8 | 255 | 592 | 67 | 7 | 1,723 |
| Cases per terminology |  |  |  |  |  |  |  |  |  |  |  |  |
| OMIM | 3,462 | 403 | 1,667 | 53 | 221 | 15 | 68 | 249 | 489 | 62 | 49 | 1,510 |
| Orphanet | 2,324 | 358 | 1,293 | 55 | 138 | 15 | 32 | 145 | 386 | 65 | 43 | 634 |
| # HPO terms per case |  |  |  |  |  |  |  |  |  |  |  |  |
| Min | 0 | 5 | 5 | 5 | 5 | 6 | 5 | 5 | 5 | 5 | - | 5 |
| Median | 12 | 8 | 8 | 17 | 12 | 11 | 16 | 30 | 15 | 10 | - | 9 |
| Max | 76 | 46 | 36 | 53 | 49 | 26 | 46 | 60 | 61 | 46 | - | 535 |

### Diagnosis retrieval across rare-disease corpora

Across the 10 rare-disease corpora and two target disease-coding ontologies (OMIM and Orphanet), RD-Embed’s later stages consistently improved diagnosis retrieval relative to general biomedical embedding baselines (Fig. 2). The overall pattern was stable across datasets with very different case styles (specialist genetics write-ups vs. fragmented or ambiguous referrals): rare-disease-specialized representations retrieved the correct diagnosis more often and at clinically meaningful shortlist sizes, whereas broad biomedical encoders frequently failed to surface the true diagnosis near the top.

**Figure 2.**
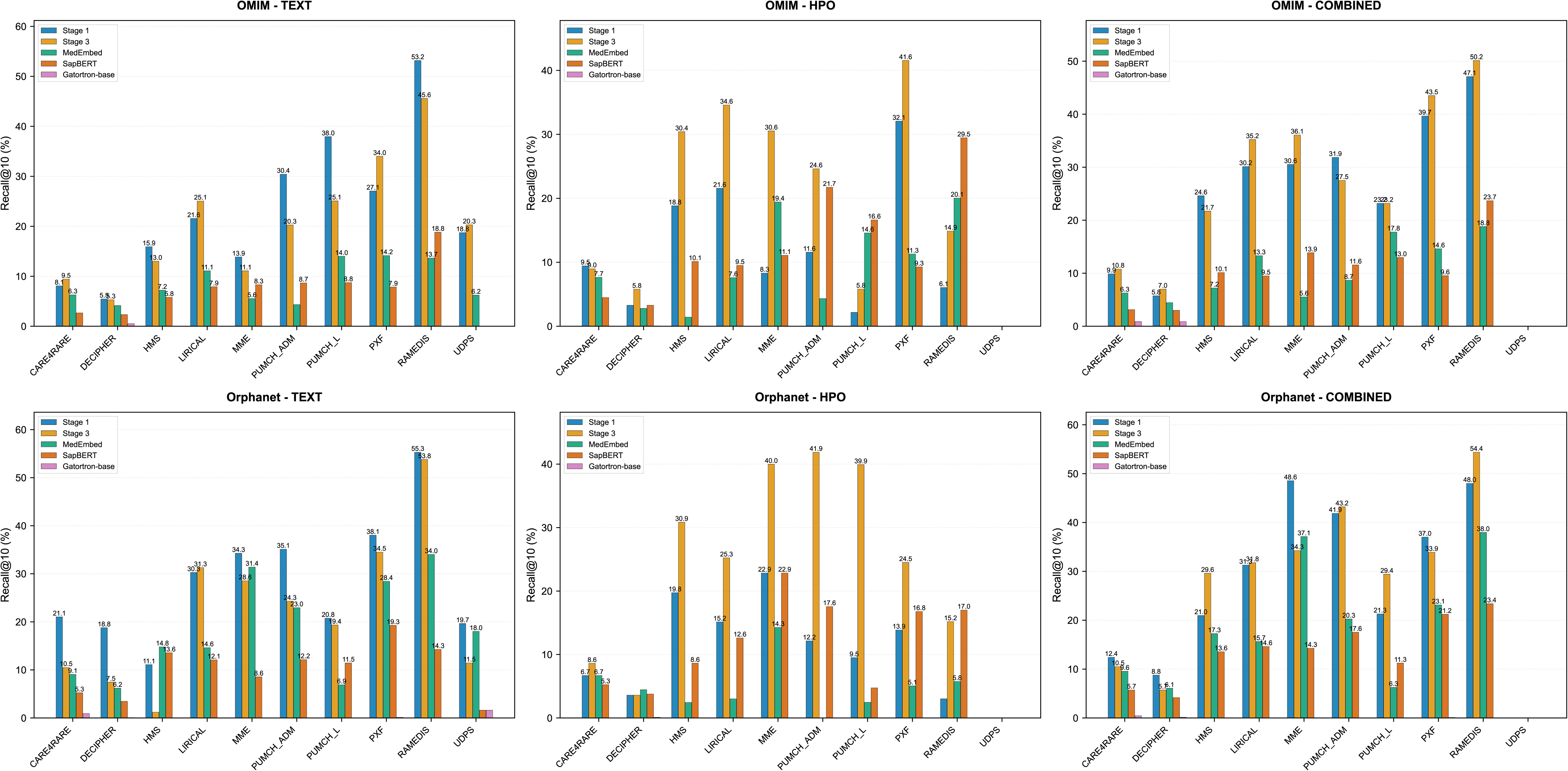
Diagnosis retrieval across rare-disease corpora and inputs. Recall@10 for diagnosis retrieval across 10 corpora evaluated against OMIM (top) and Orphanet (bottom). We test three realistic inputs: narrative notes or narrative-style representations of phenotypes (TEXT), structured phenotypes (HPO), and both (COMBINED). Models include RD-Embed Stage 1 and Stage 3 and three biomedical embedding baselines. Bars indicate the fraction of cases where the correct diagnosis appears in the top 10. UDPS includes TEXT only.

As a concrete illustration of the operating range achieved on specialist-like documentation, the best-performing setting reached ∼52% Recall@10 (Stage 3, LIRICAL corpus, OMIM, COMBINED). In contrast, strong general-purpose baselines peaked far lower in the same evaluation grid (e.g., MedEmbed ∼19% Recall@10 at its best), and other biomedical encoders were frequently near zero on difficult conditions.

Input modality had a clear – but dataset-dependent – effect. When both a narrative and structured phenotypes were used (i.e., COMBINED inputs) retrieval was often improved substantially against either modality alone (e.g., HMS under OMIM improved from ∼26% (TEXT) to ∼41% (COMBINED) for Stage 3). However, not all cohorts benefited equally, and occasional degradations were consistent with variable phenotype annotation quality and heterogeneous text informativeness. Practically, this indicates that multimodal retrieval is most valuable when the structured phenotype layer is both present and specific, but robust retrieval still requires strong performance when it is absent or noisy.

Text-only cohorts provide a clinically important stress test because early referrals often contain narrative notes without standardized phenotyping. In UDPS (text-only), Stage 3 maintained non-trivial performance (∼20% Recall@10 under OMIM), while several general biomedical baselines collapsed to near-zero recall. Full results across Recall@5/10/20/50 and all corpora are provided in the Supplementary Figs. S1-S24.

### EHR-based diagnosis retrieval

We next evaluated diagnosis retrieval on an EHR-derived corpus designed to reflect routine health-system documentation and SNOMED-aligned signals (Fig. 3; Supplementary Fig. S25). This setting directly tests whether a rare-disease representation is deployable beyond curated case reports.

**Figure 3.**
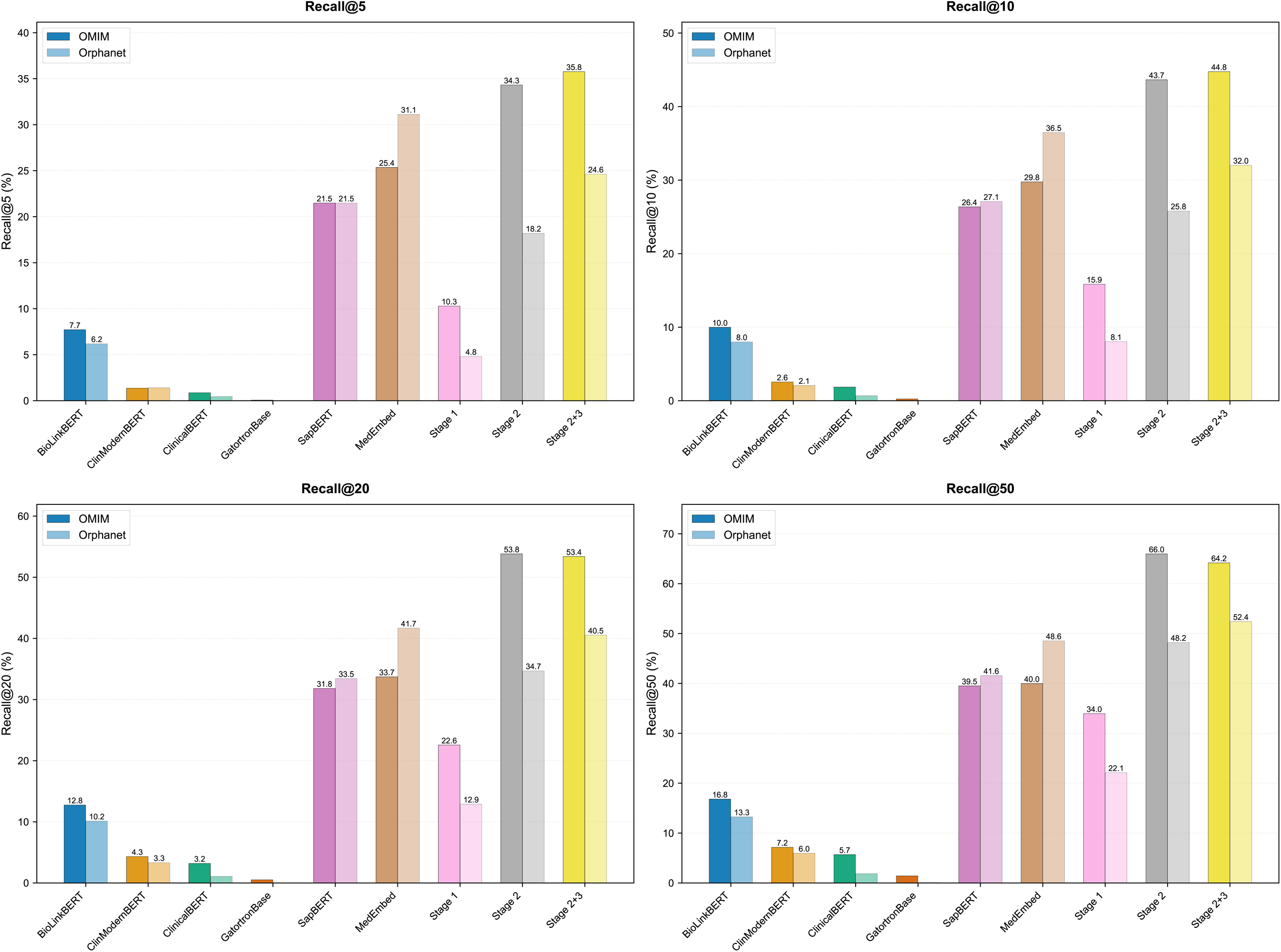
EHR-based diagnosis retrieval using routine health-system signals. Recall@10 on the EHR corpus for RD-Embed stages (Stage 1; Stage 2 clinical alignment; Stage 2+3 with graph refinement) and baseline models, evaluated on OMIM and Orphanet. Inputs are derived from routine documentation and SNOMED-aligned signals as represented in this corpus.

In this EHR context, early-stage ontology training alone was not sufficient: Stage 1 underperformed an EHR-aware baseline, as expected. After clinical alignment, performance increased markedly. As an anchor comparison, for OMIM diseases in TEXT mode, Stage 1 achieved ∼16 Recall@10, whereas the clinically aligned model reached ∼45 Recall@10 (Fig. 3), surpassing MedEmbed (∼30 Recall@10). This step change supports the engineering premise that explicit clinical alignment is required to translate curated rare-disease structure into health-system documentation.

Modality breakdown further clarifies how signals contribute in EHR-scale workflows. SNOMED codes alone carried useful diagnostic information but were consistently weaker than text alone; combining them produced the strongest results (e.g., OMIM COMBINED achieved ∼45 Recall@10). At broader shortlist sizes relevant to triage and downstream review, Recall@50 reached ∼64% (OMIM) in this EHR setting, indicating that the correct diagnosis can often be surfaced within a reviewable set even when documentation is imperfect.

### Gene identification to support genomic interpretation

To support downstream genomic interpretation, we evaluated gene identification across corpora and input settings (Fig. 4; Supplementary Figs. S26-S37). Gene prioritization is inherently more difficult than diagnosis retrieval because the candidate space is larger and the mapping from phenotype text to genes is less direct; clinical utility depends heavily on early precision.

**Figure 4.**
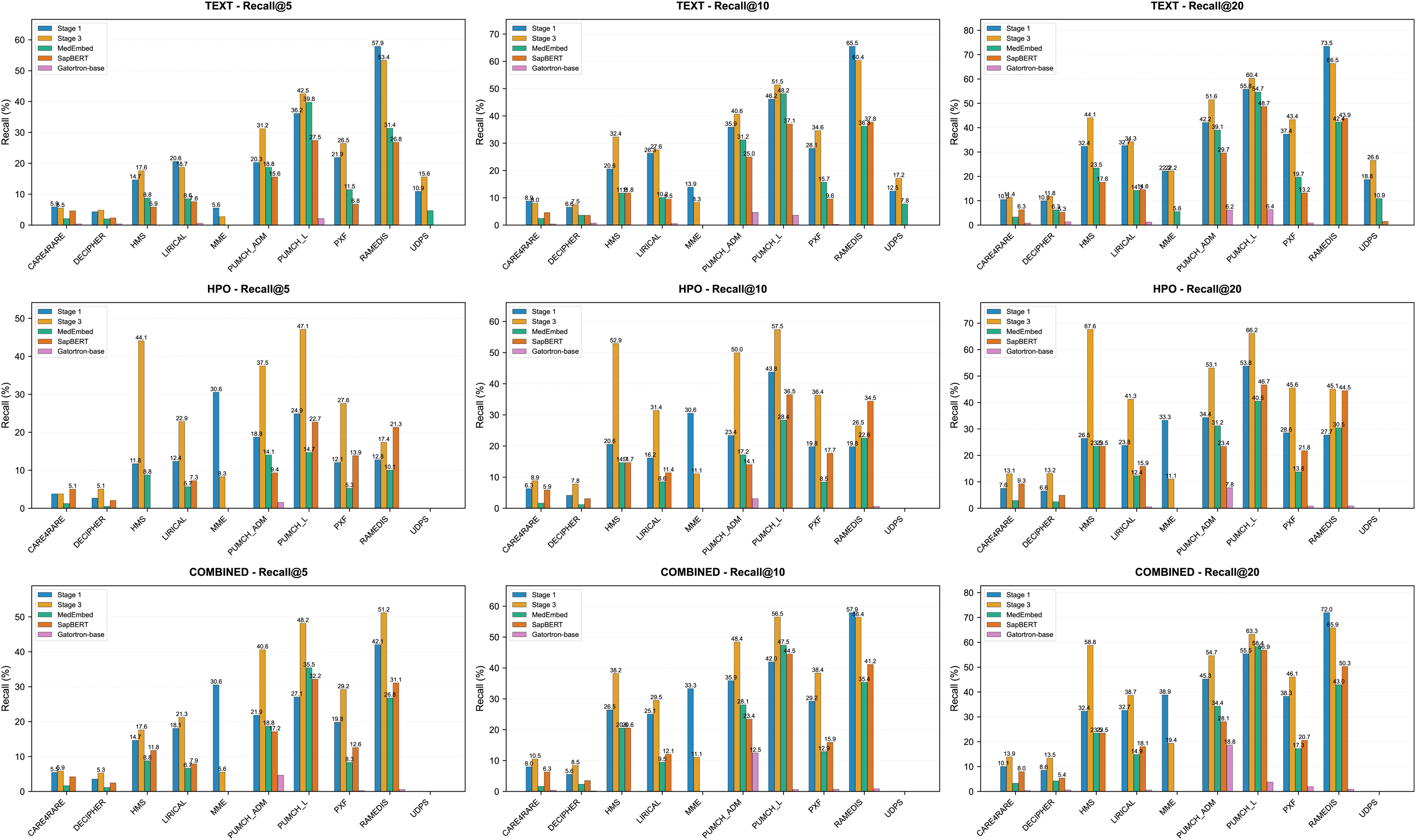
Gene prioritization across corpora and inputs. Recall@K (K=5,10,20) for identifying the causative gene across 10 corpora using TEXT, HPO, or COMBINED inputs. Models include RD-Embed Stage 1 and Stage 3 and biomedical embedding baselines. Bars indicate the fraction of cases where the causative gene is retrieved within the top K from the full gene candidate set. UDPS includes TEXT only.

Despite this difficulty, RD-Embed’s later stages improved gene recall across most corpora, with the strongest gains in cohorts where phenotypes were captured with reasonable specificity. As an anchor example, Stage 3 reached ∼45% Recall@10 for gene identification in MME (COMBINED). More broadly, COMBINED inputs frequently improved gene identification over single-modality inputs, and HPO-only inputs could be competitive in well-phenotyped datasets – consistent with curated phenotype-gene association structure being highly informative when present.

In text-only stress tests (UDPS), performance decreased but remained non-trivial: Stage 3 achieved ∼17% Recall@10 and improved further at larger shortlist sizes, supporting its use as an early, narrative-driven gene narrowing signal when structured phenotypes are unavailable.

We also evaluated two related capabilities that support gene-centric workflows when phenotyping is incomplete and when cases fall outside previously observed disease-gene-phenotype contexts. In phenotype inference (predicting missing HPO terms from partial profiles), Stage 3 improved ranking quality on 776,100 synthetic profiles (MRR 5.33, Recall@20 22.90) and remained stable across difficulty tiers (Supplementary Results S2). In zero-shot transfer to unseen diseases, Stage 3 again performed best overall but remained below standard (in-distribution) retrieval performance, reflecting the intrinsic difficulty of long-tail generalization. For example, on PXF it achieved Recall@50 49.62 for unseen-disease identification, and on DECIPHER MRR increased from 2.52 (Stage 1) to 5.31 (Stage 3) additional analyses are provided in the Supplementary Results S3.

### Differential diagnosis re-ranking on difficult cohorts

To better approximate the clinical act of refining an initial shortlist, we evaluated differential diagnosis re-ranking on four difficult cohorts (DECIPHER, LIRICAL, PXF, RAMEDIS) with candidate list sizes of k=10, 20, and 50 (Fig. 5; Supplementary Figs. S38-S39). Across corpora, RD-Embed improved ranking quality over baseline embeddings, and its advantage grew with larger differentials – an important property when clinicians must choose among many plausible rare conditions.

**Figure 5.**
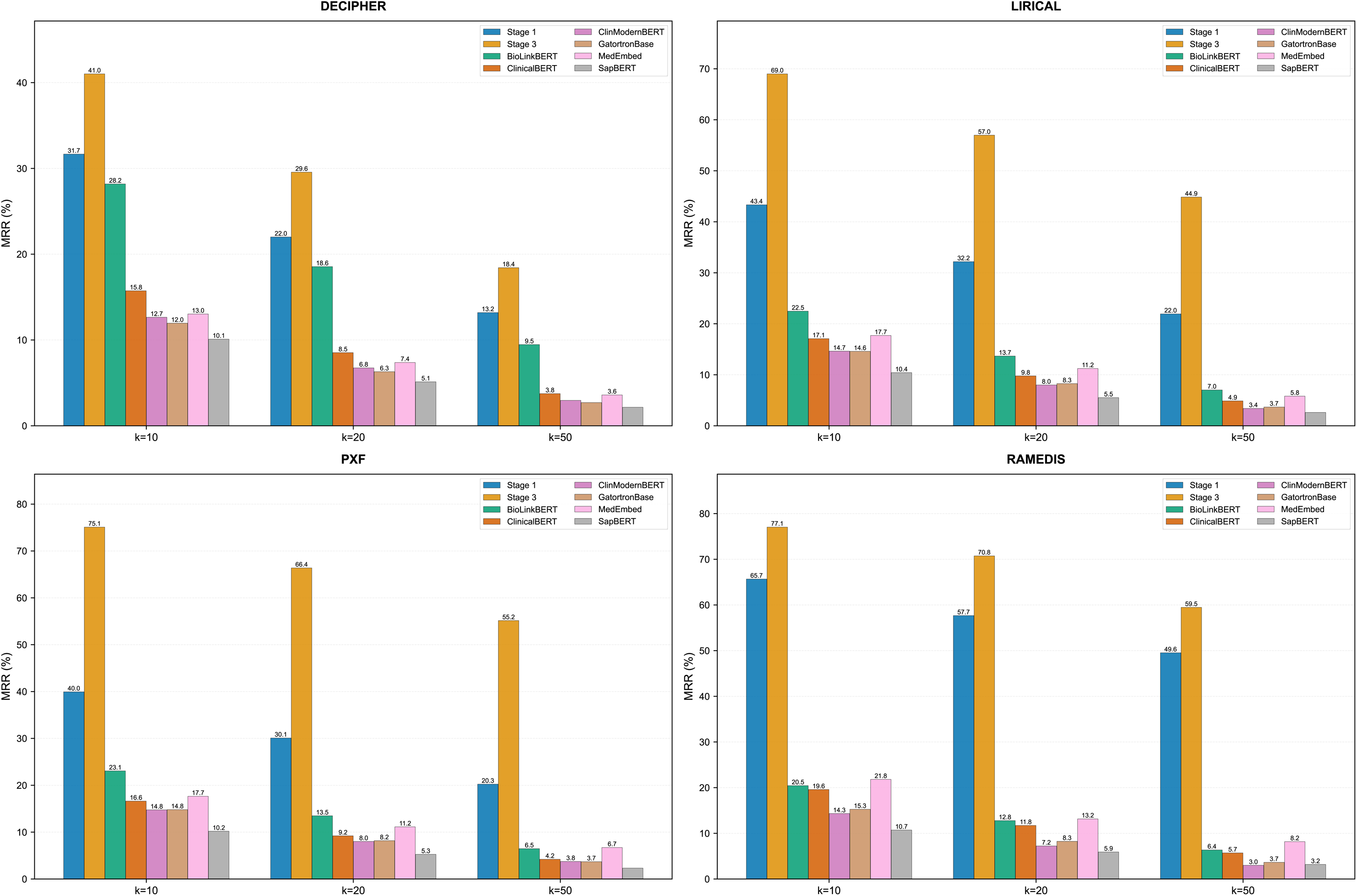
Differential diagnosis re-ranking on difficult cohorts. Mean reciprocal rank (MRR) for re-ranking candidate differentials in DECIPHER, LIRICAL, PXF, and RAMEDIS. Models re-rank candidates drawn from the initial retrieval step at k=10, 20, and 50. Higher MRR indicates that the correct diagnosis is ranked closer to the top of the re-ranked list.

As an anchor result, Stage 3 achieved a mean MRR ∼55 across the four cohorts, compared with ∼36 for Stage 1, indicating that later-stage training materially improves the ordering of candidates once a plausible set has been surfaced. In RAMEDIS at k=10, Stage 3 placed the correct diagnosis in the top three for ∼82% of cases, supporting its use as a re-ranking layer to sharpen actionable differentials.

### Comparison against large language models on difficult corpora

To contextualize RD-Embed relative to modern LLM-based clinical AI, we compared it against two on-premise open-weight models chosen to reflect realistic deployment constraints: GPT-OSS-120B run at full precision and DeepSeek-R1 run in 8-bit quantized form, representing a trade-off between quality and resource requirements. In addition, because the difficult cohorts used here impose data-use and licensing constraints, we were unable to benchmark large commercial LLMs on these datasets. Across four cohorts (Care4Rare, DECIPHER, EHR, UDPS) and two ontologies, RD-Embed was competitive and frequently superior under Orphanet, particularly in EHR-like retrieval. For example, on EHR under Orphanet, RD-Embed achieved ∼39% Recall@10 compared to ∼26% (GPT-OSS-120B) and ∼23% (DeepSeek-R1), with the gap widening at larger K. Under OMIM, the larger model more often led, but RD-Embed remained competitive and exceeded GPT-OSS-120B in selected settings at larger shortlist sizes. Together, these results illustrate that compute scale alone does not guarantee rare-disease diagnostic utility: purpose-built representations can match or exceed general models on difficult cohorts, while providing a lightweight interface that can be integrated into downstream LLM reasoning as structured retrieval context.

### Engineering-to-clinic takeaway

Taken together, the evaluations support three practical deployment modes. First, RD-Embed enables early triage from narrative-only referrals, where structured phenotyping is absent and baselines often fail. Second, it supports EHR-scale candidate surfacing from notes and codes, allowing health systems to retrieve a plausible differential from routine documentation rather than curated case reports. Third, it provides gene shortlist narrowing to a reviewable set that can complement molecular laboratory workflows and downstream interpretation. In combination, these capabilities make RD-Embed a practical bridge between curated rare-disease knowledge and real-world clinical evidence, and a compact retrieval substrate that can be consumed by LLM-based clinical systems.

## Discussion

RD-Embed addresses a practical real-world bottleneck in rare disease medicine: clinicians must retrieve plausible diagnoses and genes from fragmented, messy clinical EHR records, with partial disease phenotypes, evolving clinical symptoms and signs, and incomplete coded histories, rather than from idealized, fully curated phenotype profiles. Across evaluations that mirror this workflow, we show that RD-Embed consistently improves retrieval and ranking when compared with general biomedical embeddings, and RD-Embed remains usable in clinical settings where structured phenotyping is incomplete or absent.

Conceptually, RD-Embed learns a shared representation space that links diseases, genes, phenotypes, and clinical descriptions through a stage-wise design, from ontology-preserving base space, clinical alignment bridge to graph refinement, so that curated rare-disease structure is retained while adapting to real-world clinical evidence.

A key finding is that RD-Embed is most valuable when it supports the way rare disease workups actually unfold. When both narrative descriptions and structured phenotypes are available, multimodal inputs generally improve diagnosis and gene retrieval, consistent with the clinician’s practice of combining a story with a phenotype profile. When phenotypes are sparse or noisy, performance depends more strongly on the informativeness of the text, and gains from combining modalities are smaller or inconsistent, underscoring that the limiting factor becomes upstream documentation and annotation quality rather than model capacity. This also helps explain why specialist-like corpora yield substantially higher recall than cohorts that better resemble early, fragmented encounters. Importantly, RD-Embed retains meaningful signal in text-only settings, supporting triage and early differential generation without requiring a full HPO extraction pipeline as a prerequisite.

We conducted a EHR stress test to highlight the most deployment-relevant contribution: clinical alignment is necessary for health-system data. Ontology-only representations underperform in EHR-derived inputs, but aligning clinical text and SNOMED-linked signals to the rare-disease space (Stage 2) yields large gains and can exceed strong text-based baselines. Practically, this suggests that a rare-disease representation intended for routine care cannot rely on ontology structure alone; it must explicitly learn the mapping between how clinicians write and code and how rare-disease knowledge is curated. The relatively smaller incremental gains from graph refinement on top of Stage 2 are consistent with Stage 3 acting as a “knowledge propagation” step – improving neighbourhood structure and long-tail generalization once clinical alignment is established.

Gene prioritization results further support RD-Embed as an enabling layer for genomic interpretation, not just a disease label retriever. Although gene identification is inherently harder – because the search space is larger and clinical narratives often omit gene-discriminating details – the model can reduce thousands of possible genes to a shortlist that is closer to what clinical molecular laboratories and genetics teams can feasibly review. The consistent improvement from small to moderate candidate list sizes (for example, from top 5 to top 20) is particularly relevant for downstream variant interpretation workflows, where practical utility depends on early precision but also benefits from capturing additional true positives in a manageable review set. The same representation also supports differential diagnosis re-ranking: once a plausible candidate set is retrieved, RD-Embed improves ordering quality, with increasing advantage as the differential grows – an operating regime that reflects real-world rare disease workups, where clinicians often start with broad possibilities and iteratively narrow them.

Positioned against existing phenotype-driven tools, RD-Embed should be viewed as complementary rather than competitive. Systems such as LIRICAL, Phen2Gene, and Exomiser have demonstrated clear value when explicit, well-curated HPO profiles and high-quality disease-phenotype frequency information are available; in curated benchmarks, they can achieve very high top-k performance (e.g., LIRICAL reporting the correct gene in the top three for ∼90% of a 384-case benchmark). However, those conditions are often met only late in the diagnostic process or within specialized settings, whereas early consultations, emergency presentations, and longitudinal EHR records frequently contain partial, evolving, or narrative-only descriptions. RD-Embed explicitly targets this gap by enabling similarity-based retrieval directly from text and codes, while still benefiting from structured phenotypes when they exist. Compared with general biomedical and clinical embeddings (e.g., ClinicalBERT, GatorTron, SapBERT, MedEmbed), the gains observed here suggest that broad linguistic competence and concept retrieval are not sufficient for rare disease reasoning; preserving phenotype structure and modelling disease-gene-phenotype relationships improves both retrieval and downstream inference. In this sense, RD-Embed can be used alongside phenotype tools as an upstream retrieval layer (to propose differentials from incomplete evidence) or as a unifying representation that makes heterogeneous signals comparable.

The comparison against LLMs offers a pragmatic interpretation rather than a size contest. On difficult datasets and in EHR-aligned retrieval, a lightweight embedding model can be competitive with, and in several settings outperform LLMs, despite orders-of-magnitude differences in parameters and compute requirements. This supports a realistic deployment model in which RD-Embed provides a fast, stable retrieval substrate that can run without expensive infrastructure, while LLMs are reserved for tasks where generation, summarization, and narrative reasoning are needed. Importantly, the same embeddings can also serve as structured inputs to LLM-based clinical systems, grounding downstream reasoning in a rare-disease-focused space rather than relying solely on general-purpose priors.

Several limitations remain for this work. The evaluated datasets are retrospective and geographically biased, with limited representation of non-English settings, low-resource contexts, and diverse age groups. Errors or inconsistencies in ontology mappings – particularly between HPO, OMIM, Orphanet, and SNOMED CT – may disproportionately affect ultra-rare conditions with sparse or conflicting annotations. Zero-shot evaluations are constrained by the availability of held-out entities with clean alignments, and while relative improvements are substantial, absolute performance in long-tail generalization and phenotype inference indicates room for improvement. Finally, we do not report prospective clinical trials, human-AI comparisons, or formal uncertainty calibration; these are essential next steps to establish safety, interpretability, and impact in multidisciplinary workflows in clinical environments [15]. A convincing path forward is a staged evaluation aligned to deployment: retrospective silent-mode studies measuring whether RD-Embed improves time-to-suspicion or reduces missed candidates in real EHR trajectories, followed by prospective decision-support pilots that assess how clinicians use retrieved differentials and gene shortlists, and where the system fails. Together, these steps would determine whether unified rare-disease representations can shorten the diagnostic odyssey by making rare-disease knowledge searchable from the data clinicians actually have.

## Methods

### Data sources and knowledge resources

The training dataset comprises 26,293 manually selected published case reports (PubMed), covering 4,410 diseases and 2,992 genes (Table 1). Table 1 also summarizes the evaluation datasets, their modality availability (TEXT/HPO/SNOMED), and key long-tail properties (cases per disease; terms per case), which vary substantially across cohorts. Case selection aimed for broad disease coverage rather than concentrating on well-documented conditions, to reduce bias toward commoner rare disorders. Five authors (FT, NLTR, MWS, JK, JAL) manually assigned and verified Orphanet and OMIM identifiers for each case report.

To support unified representations across clinical text, ontologies, and biology, we augmented the case report corpus with rare-disease and biomedical knowledge sources (versions available in Oct 2025): (i) Nomenclatures and ontologies: OMIM [5], Orphanet [6], HGNC [28], HPO [3,4], Gene Ontology (GO) [29], UBERON [30], and SNOMED CT [31]; (ii) Curated mappings: Orphanet↔OMIM, Orphanet↔SNOMED, and HPO↔SNOMED cross-references; (iii) Curated relational annotations: Monarch [7], GO and Reactome [32], providing disease-phenotype, disease-gene, gene-GO, and pathway-gene links.

### RD-Embed overview

RD-Embed learns a shared embedding space for diseases, genes, phenotypes, and clinical descriptions using a three-stage progression designed to match clinical reality: ontology-preserving base space (Stage 1) → clinical alignment bridge (Stage 2) → graph refinement (Stage 3) (Fig. 6). Each stage is trained sequentially with selective freezing so that later training improves clinical alignment or relational consistency without overwriting curated semantics learned earlier. The model outputs 512-dimensional L2-normalized embeddings so that retrieval can be performed efficiently using cosine similarity.

**Figure 6.**
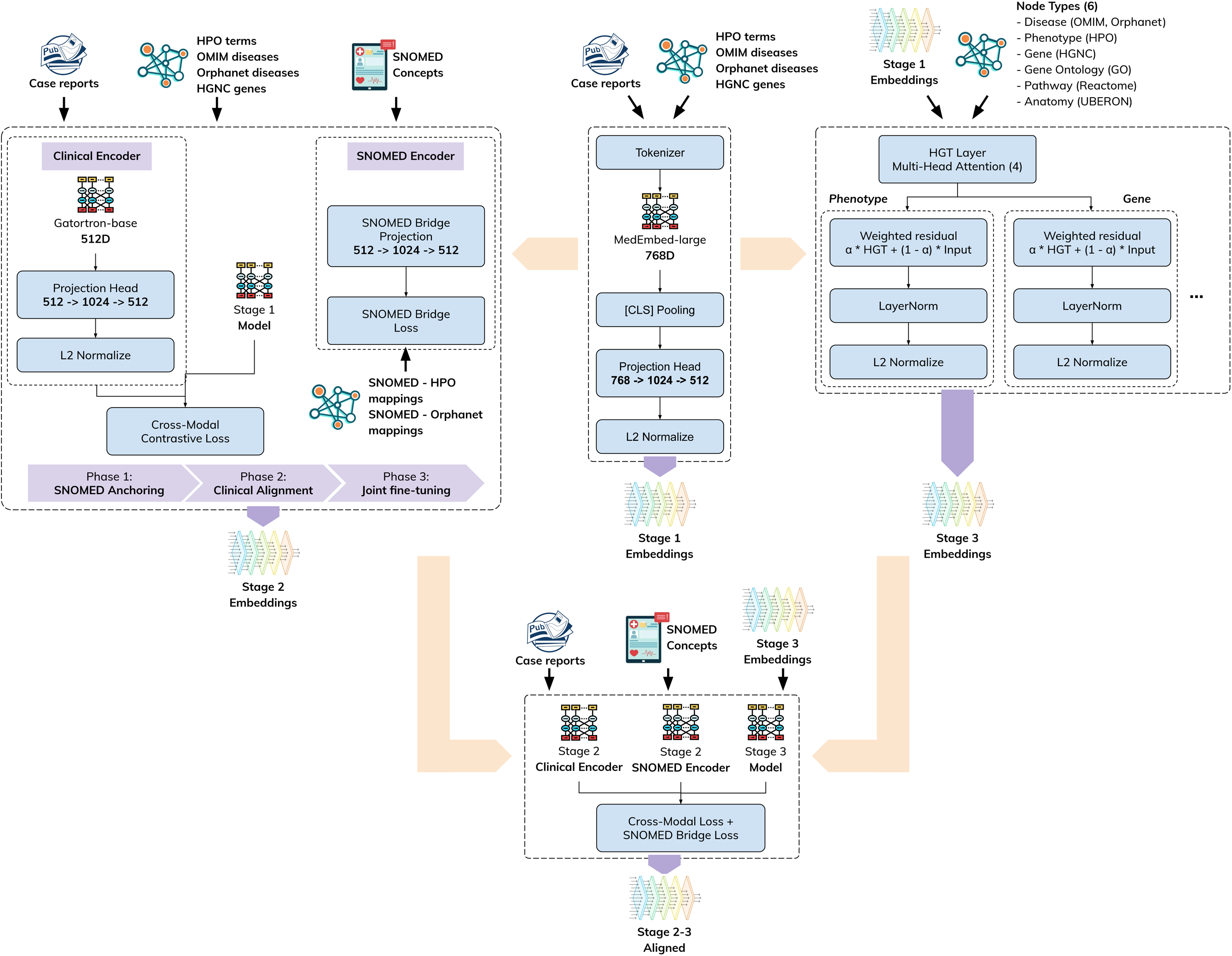
RD-Embed overview and staged training. RD-Embed maps diseases (OMIM/Orphanet), phenotypes (HPO), genes (HGNC), and clinical descriptions into a shared 512-dimensional L2-normalized embedding space using a three-stage progression: Stage 1 learns an ontology-preserving entity space from curated rare-disease resources (MedEmbed-large encoder + projection head); Stage 2 aligns clinical text and SNOMED CT concepts to this space via a dual encoder (GatorTron-base) and a SNOMED bridge trained in three phases (anchoring, clinical alignment, joint fine-tuning); and Stage 3 refines entity embeddings with a heterogeneous graph transformer (HGT) over a multi-type biomedical network (e.g., disease-phenotype, disease-gene, gene-GO/pathway/anatomy relations). A final Stage 2-3 alignment step fine-tunes the clinical encoder so clinical-text embeddings are directly comparable to the Stage 3-enhanced entity embeddings.

Unless otherwise stated, entity names, synonyms, and short textual definitions from the underlying resources (e.g., HPO labels/synonyms, OMIM and Orphanet names) are encoded using transformer encoders followed by shallow projection heads to map into the shared 512D space.

Code and configuration will be released upon acceptance.

### Stage 1: Ontology-aware contrastive learning

#### Objective

Stage 1 initializes the rare-disease embedding space from curated structure so that diseases, phenotypes, and genes occupy coherent neighbourhoods even before any EHR-specific alignment.

#### Encoder and projection

The base text encoder is MedEmbed-large-v0.1 [23]. The 768-dimensional pooled representation is mapped through a two-layer MLP projection head (768→1024→512, GELU activation) and L2-normalized.

#### Positive pairs

We construct cross-entity positive pairs from ontologies and databases including HPO, OMIM, Orphanet, and HGNC [3,4,5,6,28]. Pair types include disease-phenotype (127,568 OMIM-HPO and 421,673 Orphanet-HPO), disease-gene (4,279 OMIM-HGNC and 5,186 Orphanet-HGNC), and phenotype-phenotype positives via curated cross-references and ontology hierarchy relations, yielding 563,706 positive pairs across entity types.

#### Negatives

To encourage fine-grained discrimination among clinically confusable entities, we combine: (i) hard negatives sampled from differential diagnosis lists (n=2 per anchor), (ii) random negatives from the entity pool (n=1 per anchor), and (iii) in-batch negatives (all other samples in the batch).

#### Loss and optimization

We train using a symmetric contrastive objective (InfoNCE) over cosine similarity with temperature τ=0.05, computed in both anchor→positive and positive→anchor directions (Eq. 1; full form provided in Supplementary Methods). Optimization uses AdamW (weight decay 0.01), batch size 32, and differential learning rates (1×10⁻□ for the encoder; 2×10⁻□ for the projection head). Training runs for up to 30 epochs with early stopping on a 70/15/15 split stratified by pair type.

### Stage 2: Clinical text alignment using a SNOMED bridge

#### Objective

Stage 2 aligns how rare disease concepts appear in clinical documentation to the Stage 1 entity space. This stage targets settings where structured phenotyping is incomplete and evidence is expressed through narrative text and coded histories.

#### Dual-encoder setup

A clinical transformer encoder maps clinical text spans and SNOMED concept strings into the same 512D space, while Stage 1 remains fixed as the target representation for diseases/genes/phenotypes and ontology-derived mention strings. We use GatorTron-base [22] as the clinical encoder, with a projection head matching Stage 1’s output dimensionality and normalization.

### SNOMED initialization and bridge

SNOMED CT concept embeddings are initialized by averaging Stage 1 embeddings for mapped HPO and Orphanet entities using curated cross-references, then refined with hierarchical smoothing across SNOMED parent-child links. A lightweight projection network produces 512D SNOMED embeddings that serve as an intermediate “bridge” between EHR-coded concepts and the rare-disease entity space.

**Training phases.** Stage 2 training proceeds in three phases (Eqs. 2-4; full forms in Supplementary Methods):

1. SNOMED anchoring: fit the SNOMED projection network to Stage 1 targets using mean-squared error (Eq. 2).
2. Clinical alignment: train the clinical encoder using a bidirectional contrastive loss between clinical text segments and linked entities, with Stage 1 frozen (Eq. 3; in-batch negatives).
3. Joint fine-tuning: update the clinical and SNOMED encoders (and optionally Stage 1 projection layers) under a composite objective combining cross-modal alignment, code-bridge loss, and hierarchical consistency terms (Eq. 4; λ₁=0.3, λ₂=0.0 in this study).

We use 4,096-token windows with overlap for long notes and early stopping based on mean reciprocal rank for entity retrieval from clinical text.

### Stage 3: Graph refinement with a heterogeneous graph transformer

#### Objective

Stage 3 refines entity embeddings using biomedical relational structure so that the representation better approximates rare-disease knowledge graphs and supports propagation across diseases, phenotypes, genes, pathways, and anatomy.

#### Graph model

We use a heterogeneous graph transformer (HGT) [33], which applies type-and relation-specific attention to aggregate neighbour information for multi-type graphs. Prior rare-disease graph efforts (e.g., PhenoKG) support the utility of phenotype-genotype graphs for long-tail discovery and diagnostic ranking [34–38].

#### Graph construction

Nodes include diseases, phenotypes, genes, GO terms, pathways, and anatomical structures, initialized with frozen Stage 1 embeddings. Edge types include disease-phenotype and disease-gene links, disease-disease mappings/similarity, gene-phenotype and gene-pathway relations, gene-anatomy links, and ontology hierarchy edges across HPO, GO, and UBERON. A full specification of node/edge types, counts, and filtering is provided in Supplementary Methods.

### Training and residual mixing

We train a single HGT layer with four attention heads (selected to limit over-smoothing) using a contrastive objective over graph-linked triplets sampled within node types (Eq. 5; full form in Supplementary Methods). To preserve semantic content while allowing structure-driven refinement, Stage 3 learns a per-node-type residual mixing parameter that combines normalized HGT outputs with normalized input embeddings (Eq. 6; full form in Supplementary Methods). Edge sampling is weighted to emphasize high-information relationships (e.g., rarer phenotypes).

### Stage 2–3 alignment

Because Stage 3 modifies the entity space, we re-align the Stage 2 clinical encoder to the Stage 3-enhanced entity embeddings. Specifically, we load the trained Stage 2 model, substitute Stage 1 entity embeddings with Stage 3 embeddings, and fine-tune the clinical encoder for 10 epochs with a reduced learning rate (5×10⁻D) using the same cross-modal contrastive loss as Stage 2 clinical alignment. This produces a single pipeline in which clinical text embeddings are directly comparable to Stage 3 entity embeddings.

### Training pipeline and implementation

Stages are trained sequentially: Stage 1 on ontology-derived contrastive pairs; Stage 2 on narrative representations of SNOMED terms, case reports and SNOMED mappings conditioned on a frozen Stage 1 target space; Stage 3 on the heterogeneous biomedical graph using Stage 1 embeddings as fixed node features. Models are implemented in PyTorch, with PyTorch Geometric used for graph components. Training used mixed precision on H100/H200 GPUs with fixed random seeds and deterministic operations where available.

### Evaluation design

We evaluate RD-Embed with experiments that map to clinical workflow questions and representation properties, comparing Stage 1/2/3 to baselines. Unless otherwise stated, retrieval is performed by cosine similarity between a query embedding (case text and/or phenotype profile) and candidate embeddings (diseases or genes), with performance measured using mean reciprocal rank (MRR) and Recall@K.

### Core workflow evaluations

1. Disease ranking: retrieve the correct disease given TEXT, HPO, or COMBINED inputs; evaluate under OMIM and Orphanet candidate spaces.
2. EHR diagnosis retrieval: evaluate retrieval using EHR-derived inputs (SNOMED-aligned text and/or codes) to test robustness on routine health-system documentation.
3. Gene identification: retrieve causative genes given TEXT, HPO, or COMBINED inputs; for multiple true genes, use the best rank among positives.
4. Differential diagnosis ranking: re-rank a provided candidate differential list for each case (rather than the full ontology), using MRR/Recall@K and bootstrap confidence intervals.

Additional protocols are described in Supplementary Results and cover hierarchy preservation and clustering probes (Supplementary Results S1); synthetic masking for phenotype inference (Supplementary Results S2); “seen vs unseen” partitions for zero-shot (Supplementary Results S3) and compositional/perturbation tests for embedding arithmetic (Supplementary Results S4).

### Test datasets, modalities, and baselines

#### Evaluation datasets

Evaluation uses the datasets summarized in Table 1, spanning registries, published case collections, and health-system EHR data. Case-report/case-collection datasets include HMS, LIRICAL, MME, PUMCH-ADM, PUMCH-L, and RAMEDIS (assembled using the protocols in [15] and [40], as applicable). Registry phenotype-profile datasets include DECIPHER (HPO profiles; source [39]) and Care4Rare (HPO profiles; Care4Rare Canada). EHR datasets include UDPS (free-text notes from first genetics encounter; local cohort) and an EHR rare-disease subset derived from [41] (8,963 patients with structured longitudinal records summarized into a pre-diagnosis SNOMED problem-list profile).

#### Evaluation modalities

We evaluate three query modalities depending on dataset availability (Table 1).

- TEXT: raw clinical narratives when available (notably UDPS; Table 1). For profile-only datasets (HPO or SNOMED), we generate a text surrogate by concatenating concept labels (one per line).
- HPO (or SNOMED for EHR): mean pooling of embeddings for the provided HPO terms (or SNOMED concepts for EHR).
- COMBINED: mean pooling of the TEXT representation and the HPO/SNOMED profile representation when both are available.

#### Baselines

MedEmbed [23] underpins Stage 1 and is also evaluated directly. Additional baselines include SapBERT [24], ClinicalBERT [20], ClinModernBERT [21], BioLinkBERT [19], and GatorTron [22] (also used as the Stage 2 clinical encoder).

## DATA AVAILABILITY

This study makes use of data generated by the DECIPHER community [39]. A full list of centres who contributed to the generation of the data is available from https://deciphergenomics.org/about/stats and via email from. DECIPHER is hosted by EMBL-EBI and funding for the DECIPHER project was provided by the Wellcome Trust [grant number WT223718/Z/21/Z]. DECIPHER and the original sources of the data displayed on DECIPHER bear no responsibility for the analysis or interpretation of the data that you are performing.

DECIPHER provides data in good faith as a research tool, but without verifying the accuracy, clinical validity or utility of the data. DECIPHER makes no warranty, express or implied, nor assumes any legal liability or responsibility for any purpose for which the data are used. The study also makes use of data collected by the Care4Rare Canada Consortium and shared through Genomics4RD, a rare disease data sharing platform. These initiatives are supported in part by Genome Canada and the Ontario Genomics Institute (OGI-147), the Canadian Institutes of Health Research, Ontario Research Fund, Genome Alberta, Genome British Columbia, Genome Quebec, and Children’s Hospital of Eastern Ontario Foundation.

Other data sources used in the study:

- The HMS, LIRICAL, MME, RAMEDIS, PUMCH-ADM and PUMCH-L corpora were retrieved from [15]
- The PXF corpus was retrieved from [40]
- The EHR corpus was used from [41]

## Acknowledgements

SSJ is supported by National Medical Research Council Clinician Scientist Award (NMRC/CSAINVJun21-0003) and (NMRC/CSAINV24jul-0001). TG is supported by NMRC/CSAINVJun21-0003 and by the National Medical Research Council (NMRC) Research, Innovation and Enterprise (RIE2025) Centre Grant seed funding [NMRC/CG1/006/2021-KKH]. The computational work for this article was partially performed on resources of the National Supercomputing Centre (NSCC), Singapore (https://www.nscc.sg).

## Authors’ contributions

Conceptualisation of research aims and goals (TG, HC, SSJ). Methodology development (TG). Validation and rigour (TG, NK, HC, TYW, SSJ). Analysis and interpretation (TG, HC, SSJ). Investigation and collection of data (TG). Data curation (TG, FT, NLTR, MWS, JK, JAL). Writing original draft (TG, NK, HC, TYW, SSJ). All authors read and approved the final manuscript. Funding acquisition (SSJ, NK). The authors would like to thank Ms Cecilia A. Chandra (KK Research Centre, KK Women’s and Children’s Hospital), Dr Julie Lecarpentier-Guillou-Keredan (DECIPHER) and Dr Magda Price (Care for Rare Canada) for their support in preparing the data sharing agreements.

## Ethical approval

The UDPS corpus consists of data that was collated from patients enrolled in the Singapore Undiagnosed Disease Programme (IRB approval 2019/2243).

