## Supplementary Material for "RD-Embed: Unified representations of rare-disease knowledge from clinical records"

### Supplementary Results

#### S1. Ontology preservation

**Task:** Ontology Preservation assesses whether the embedding space preserves the hierarchical and relational structure of biomedical ontologies (HPO, Orphanet, OMIM), a prerequisite for using embeddings as proxies for curated knowledge graphs. The evaluation encodes ontology terms and measures structural consistency through three complementary analyses: (1) hierarchy preservation, testing whether child nodes are closer to their parents than to random nodes; (2) transitivity, verifying that distances respect is-a chains in the ontology hierarchy; and (3) sibling similarity, confirming that nodes sharing a parent are more similar than random pairs. Additionally, disease and phenotype clustering is evaluated using Orphanet disease classifications and top-level HPO categories, with performance measured via silhouette score, adjusted mutual information (AMI), and within-class versus between-class cosine similarity. This mirrors ontology-preservation probes used in prior biomedical embedding studies and validates that the learned representations reflect biomedical semantics rather than superficial lexical patterns.

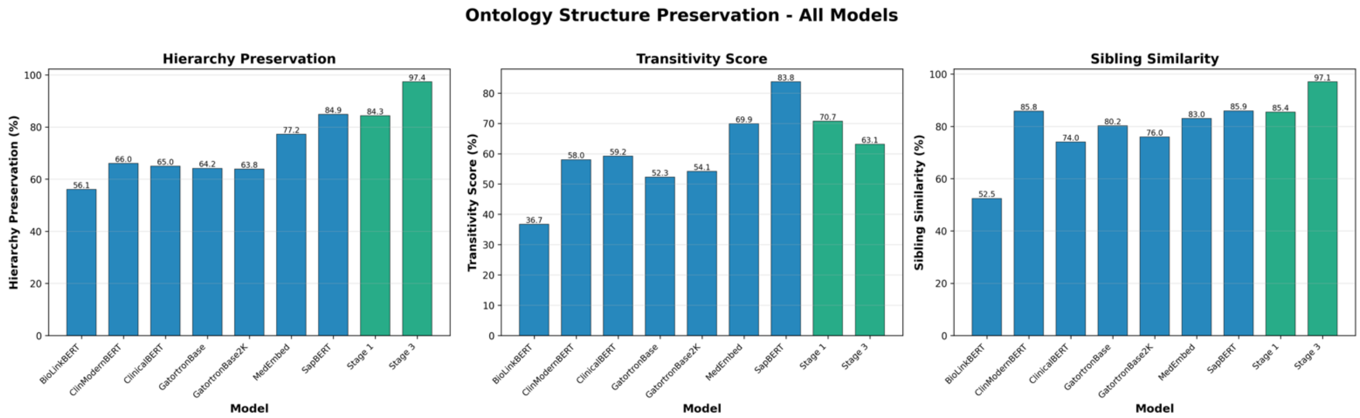


**Figure 1.** Comparative overview of the ontology structure preservation across all models

**Results:** Stage 3 substantially improves ontology structure preservation over both Stage 1 and SapBERT, achieving 97.39% hierarchy preservation versus 84.35% for Stage 1 and 84.86% for the best baseline SapBERT (Δ+13.0 and +12.5 percentage points, respectively) – depicted in Fig. 1. For disease-phenotype (*has_phenotype*) edges, Stage 3 reaches 94.16% hierarchy preservation compared with 85.67% for Stage 1 (Δ+8.49 points), and for disease-gene (*has_gene*) edges 98.63% versus 68.53% (Δ+30.10 points), indicating that the graph-enhanced Stage 3 embeddings best approximate curated rare disease knowledge graphs (see Fig. 2). Clinically, preserving these hierarchical relationships is important because patients rarely present with complete phenotype lists; models must recognize that related findings (for example, cardiomyopathy, arrhythmia, and sudden cardiac death) should map to nearby disease mechanisms and diagnostic candidates even when only a subset of features is documented.

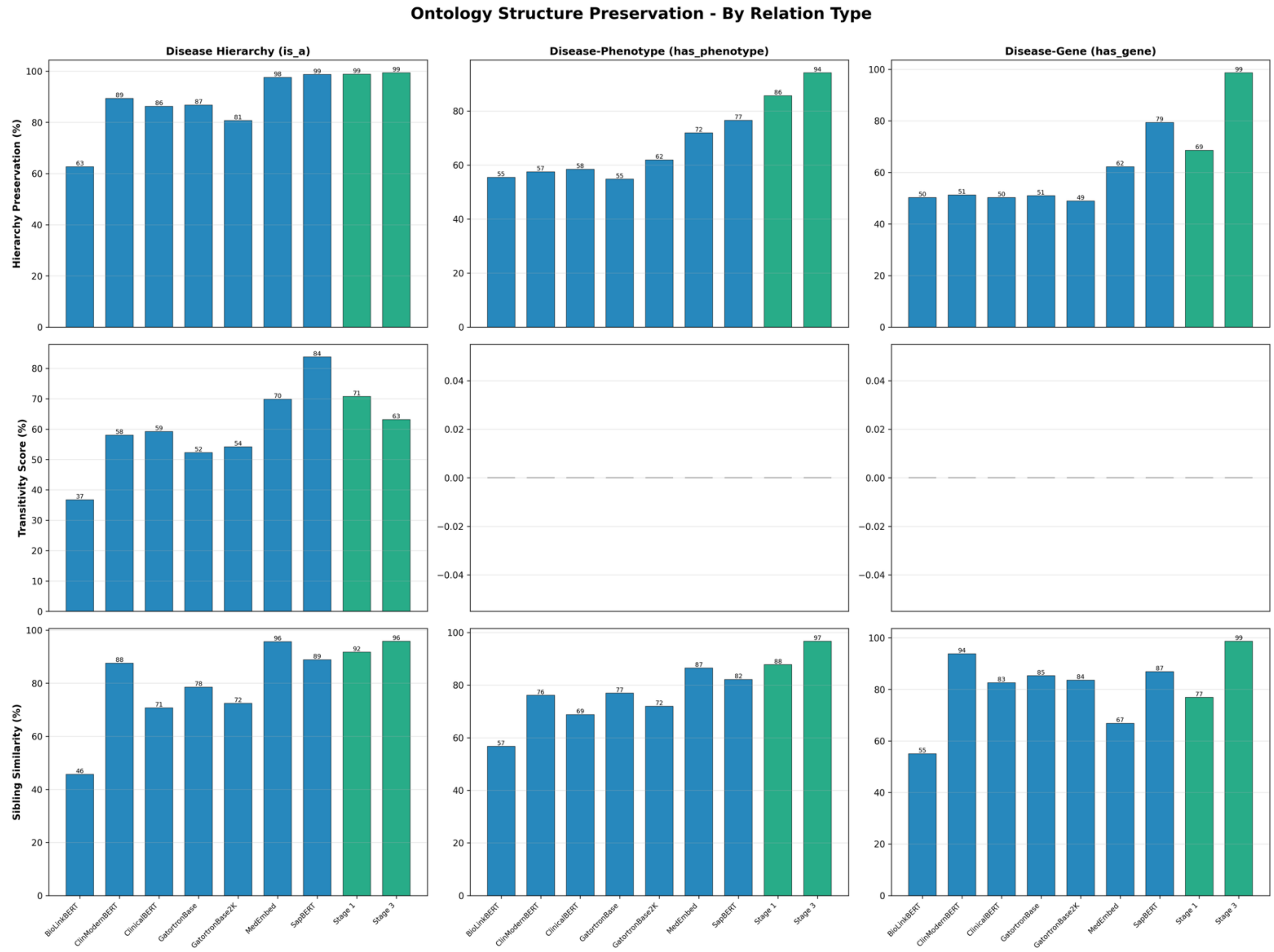


**Figure 2.** Comparative overview of the ontology structure preservation across all models, split by relation type

Sibling similarity shows a similar pattern: Stage 3 achieves 97.09% versus 85.44% for Stage 1 and 89.08% for MedEmbed, confirming that closely related diseases and phenotypes cluster tightly in the embedding space. Disease clustering over 10,016 Orphanet diseases across 35 clinical classes yields a within-between similarity gap of 0.094 for Stage 3 (see Fig. 3), 3-4× larger than the baseline mean (0.0217) and clearly higher than SapBERT (0.0652), while ClinicalBERT, ClinModernBERT, and GatorTron-base exhibit poor separation (within and between ≈0.77, gap ≈0.01). A t-SNE visualization of the outcome is presented in Fig. 4 with some groups showing tight, well-separated clusters while others exhibit more dispersed distributions. Notably, the largest category, “*Developmental anomalies during embryogenesis*” (3,090 diseases), forms multiple clusters across the embedding space, suggesting heterogeneity within this broad classification.  Several smaller categories like "*Cardiac*", "*Haematological*", and "*Genetic*" diseases form more compact, distinct clusters, indicating greater similarity within these disease groups. This pattern mirrors clinical reality: broad categories such as developmental anomalies aggregate many mechanistically distinct syndromes with overlapping features, whereas narrower domains (e.g., primary cardiac or haematological disorders) tend to have more consistent phenotype–gene structure and therefore form tighter diagnostic neighbourhoods.

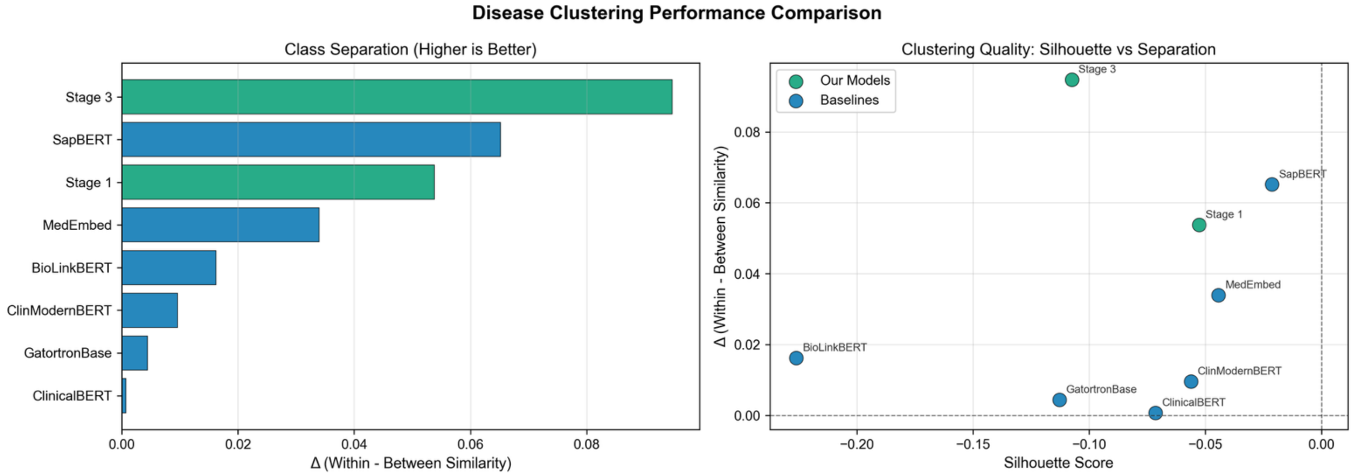


**Figure 3.** Orphanet disease clustering performance across all models using the Orphanet classification

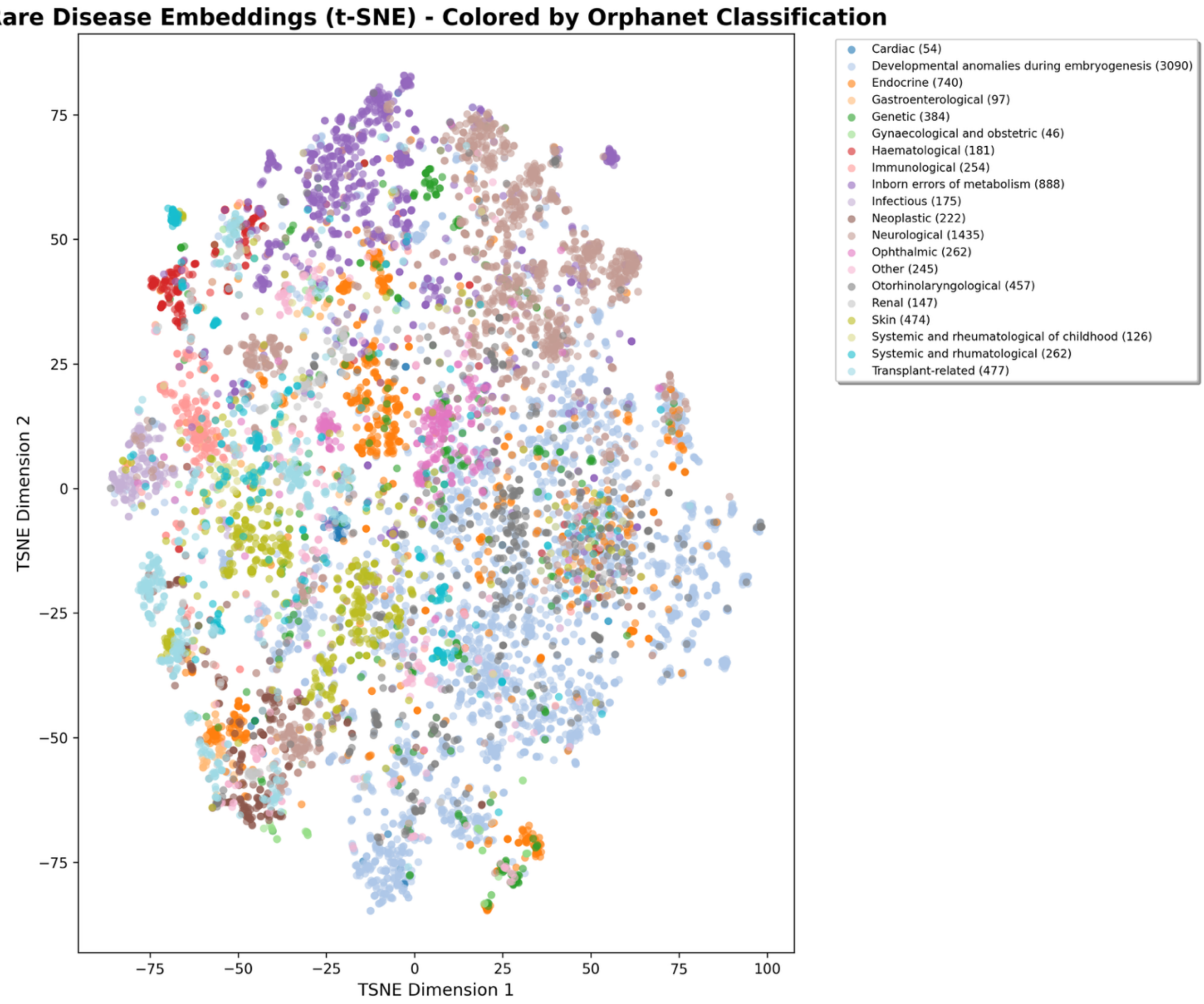


**Figure 4.** t-SNE visualization of the Orphanet clustering outcome of the Stage 3 RD-Embed model.

For 18,506 HPO terms across 23 top-level categories (Fig. 5), Stage 3 is the only model with a positive silhouette score (0.0059), a 14-fold improvement over the average baseline (−0.031) and markedly better than Stage 1 (−0.0016) and BioLinkBERT (−0.1466), highlighting that the Stage 3 heterogeneous graph transformer is critical for preserving phenotype category structure. Similar to disease clustering, Fig. 6 depicts a t-SNE visualization of the HPO clustering outcome, with categories, such as,

"*Abnormality of the cardiovascular system*", "*Abnormality of head or neck*" or "*Abnormality of metabolism/homeostasis*" forming well-delineated clusters in different regions of the space.

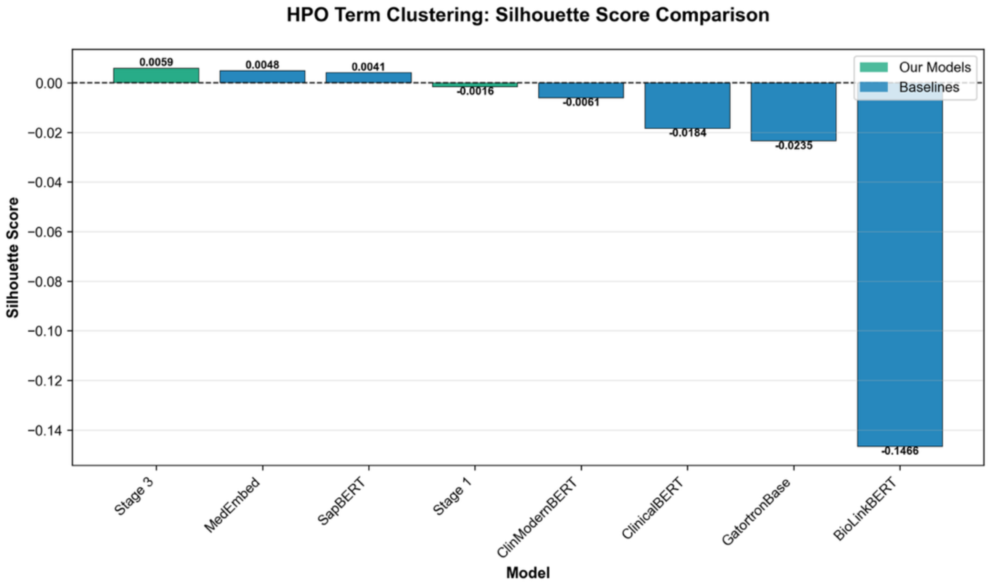


**Figure 5.** HPO term clustering performance across all models using the HPO top-level phenotypic abnormalities as categories

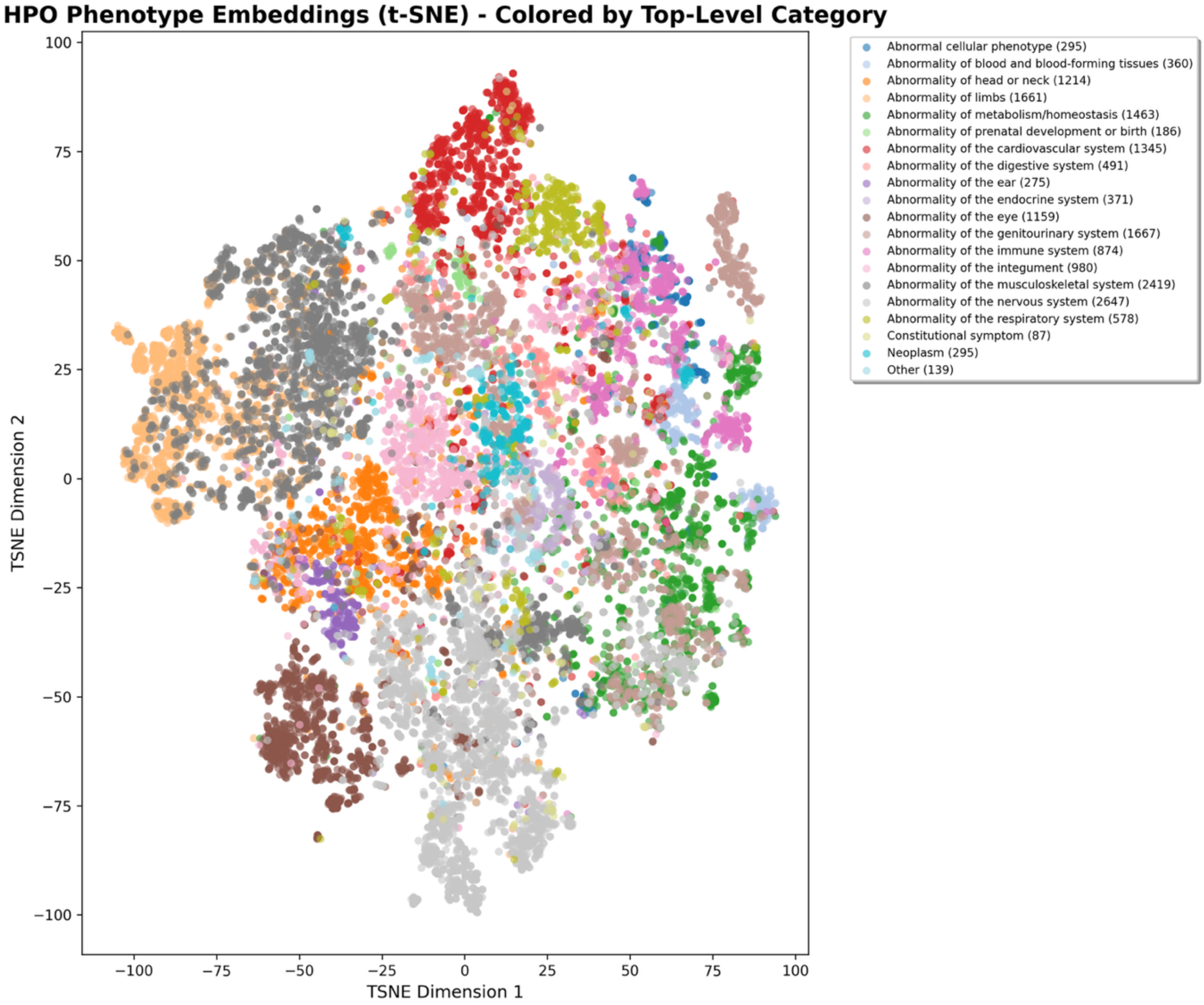


**Figure 6.** t-SNE visualization of the HPO clustering outcome of the Stage 3 RD-Embed model.

#### S2. Phenotype inference

**Task:** Measure phenotype inference performance by ranking missing phenotypes from partial HPO profiles using synthetically masked data and testing whether the model can generalize phenotype associations and recover clinically relevant findings not explicitly provided. For each disease, complete phenotype sets were derived from OMIM and Orphanet resources and randomly partitioned into "observed" and "missing" (held-out) subsets according to a tiered difficulty scheme, with easy, medium, and hard settings corresponding to 20%, 35%, and 50% of phenotypes held out, respectively, and with at least one phenotype always retained as observed. Multiple random splits were generated per disease and repeated across corpus iterations with distinct random seeds to ensure robustness of the evaluation. Model performance was quantified using Recall@K for the recovery of missing phenotypes, with bootstrap confidence intervals used to characterize statistical uncertainty in the estimates. Results were further stratified by difficulty tier (for example, by the number of observed versus missing phenotypes) and by corpus subsets, providing a nuanced assessment of phenotypic reasoning capabilities that is conceptually aligned with HPO-based phenotype association benchmarks developed by the Monarch Initiative.

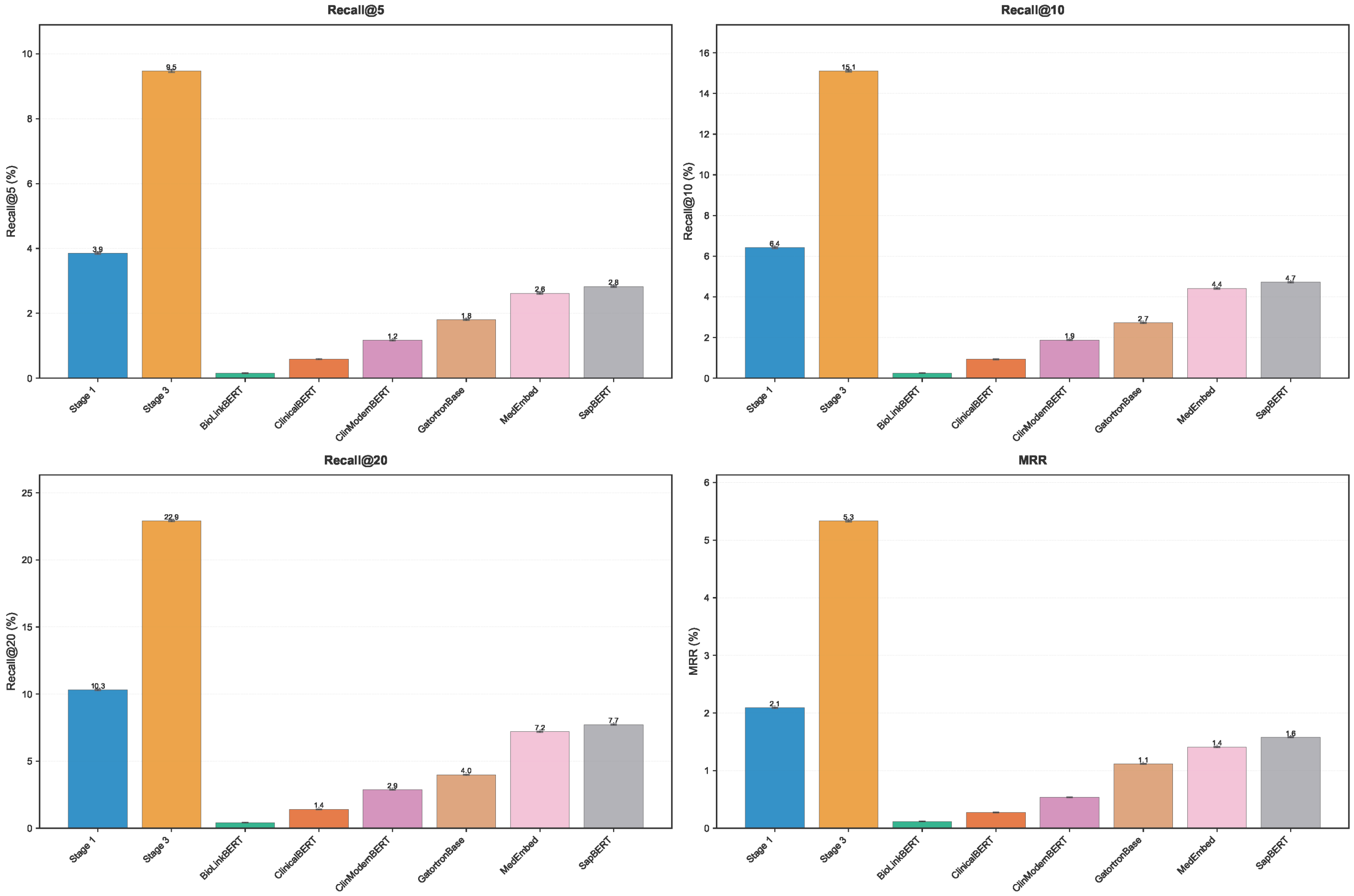


**Figure 7.** Phenotype inference from disease diagnosis. Performance metrics for the reverse phenotype inference task, where models predict HPO terms from disease diagnoses.

**Results:** On 776,100 synthetic HPO profiles (mean 15.6 observed and 8.4 missing phenotypes), Stage 1 shows modest gains over SapBERT (2.09 vs 1.58 MRR; 32.4% improvement), whereas Stage 3 dramatically improves performance, reaching 5.33 MRR and 22.90 Recall@20 (Δ+3.24 MRR and +15.18 Recall@20 over SapBERT, corresponding to 237.6% relative recall improvement; Fig. 7). Mean rank of missing phenotypes improves from 2,358 (SapBERT) to 419 (Stage 3), a 5.6-fold reduction, bringing many clinically plausible candidates into a manageable shortlist. Clinically, “missing phenotypes” can reflect features not yet elicited, under-documented, or not recognized as relevant; embedding-based suggestions may therefore support guided history-taking, targeted examination, and structured phenotyping during iterative diagnostic work-up.

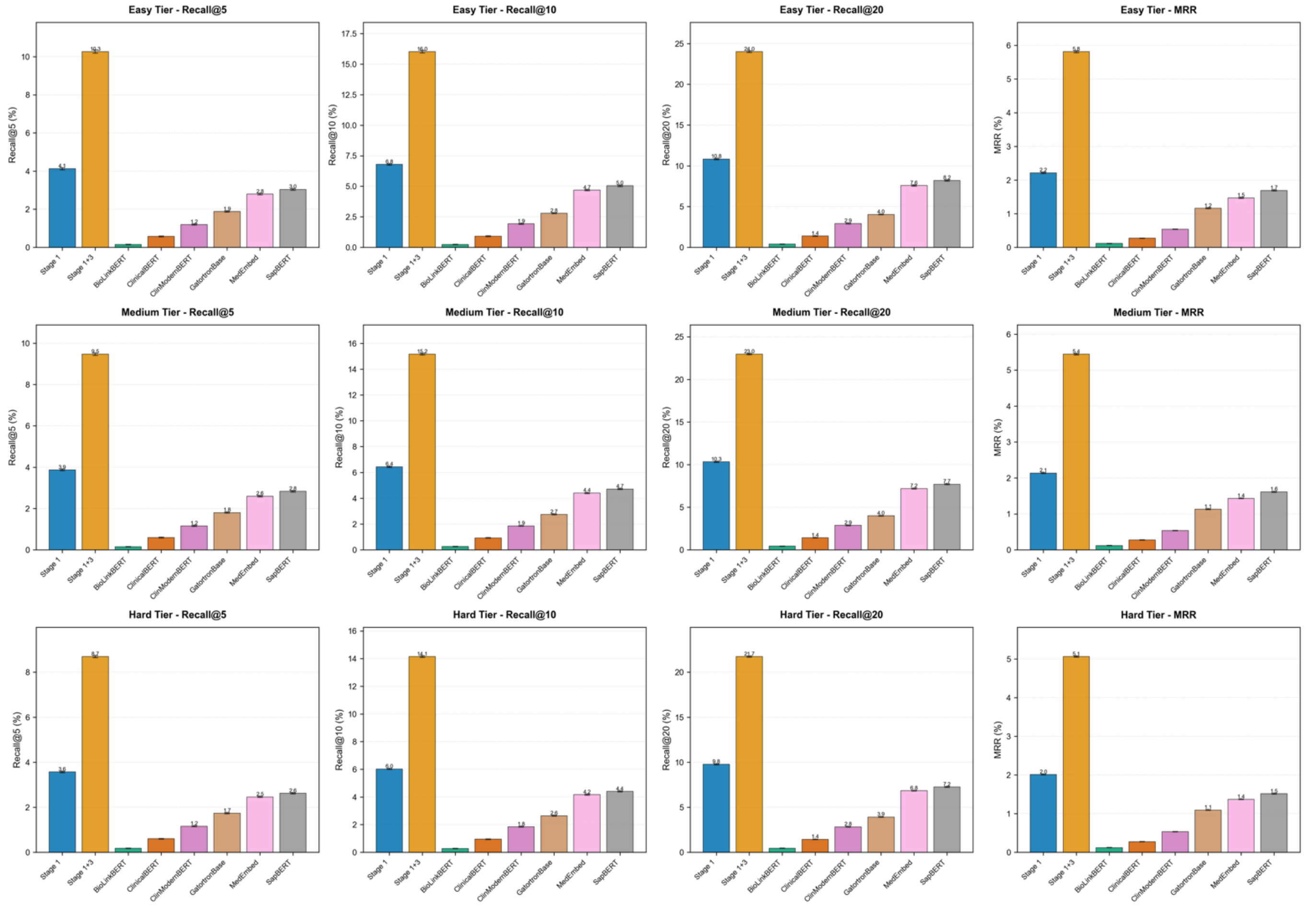


**Figure 8.** Phenotype inference results across all models and complexity tiers

Stage 3 maintains robust performance across difficulty tiers (Fig. 8): MRR is 5.81 (easy), 5.45 (medium), and 5.06 (hard), an 8% degradation from easiest to hardest profiles, while relative gains over SapBERT remain stable (≈234–244%). In clinical terms, for a typical profile, Stage 3’s 22.90 Recall@20 means ≈4.6 of 8-9 missing phenotypes appear among the top 20 suggestions, compared with ≈1.5 for SapBERT, supporting its use for guided deep phenotyping.

#### S3. Zero-shot transfer

**Task:** Zero-shot transfer evaluates a model’s capacity to generalize beyond previously seen disease-phenotype-gene data, testing whether learned representations can infer novel associations without additional training. This capability is particularly critical for rare and emerging disorders, where labelled examples are scarce and new gene-phenotype links frequently arise. More concretely, it measures retrieval performance on entities absent or underrepresented in training data. Test cases are partitioned into "seen" and "unseen" categories based on whether their associated diseases/genes appeared in the training corpus. The same retrieval setup as diagnosis and gene identification is used, with case embeddings generated from TEXT, HPO, or COMBINED inputs, and candidate entities ranked by cosine similarity. Performance is reported separately for all cases and unseen-only subsets using mean reciprocal rank (MRR) and Recall@K, with bootstrap confidence intervals to characterize uncertainty and variability. This evaluation tests a critical capability for rare disease models: generalization to novel or sparsely represented entities, which is essential given the long-tail distribution of rare diseases.

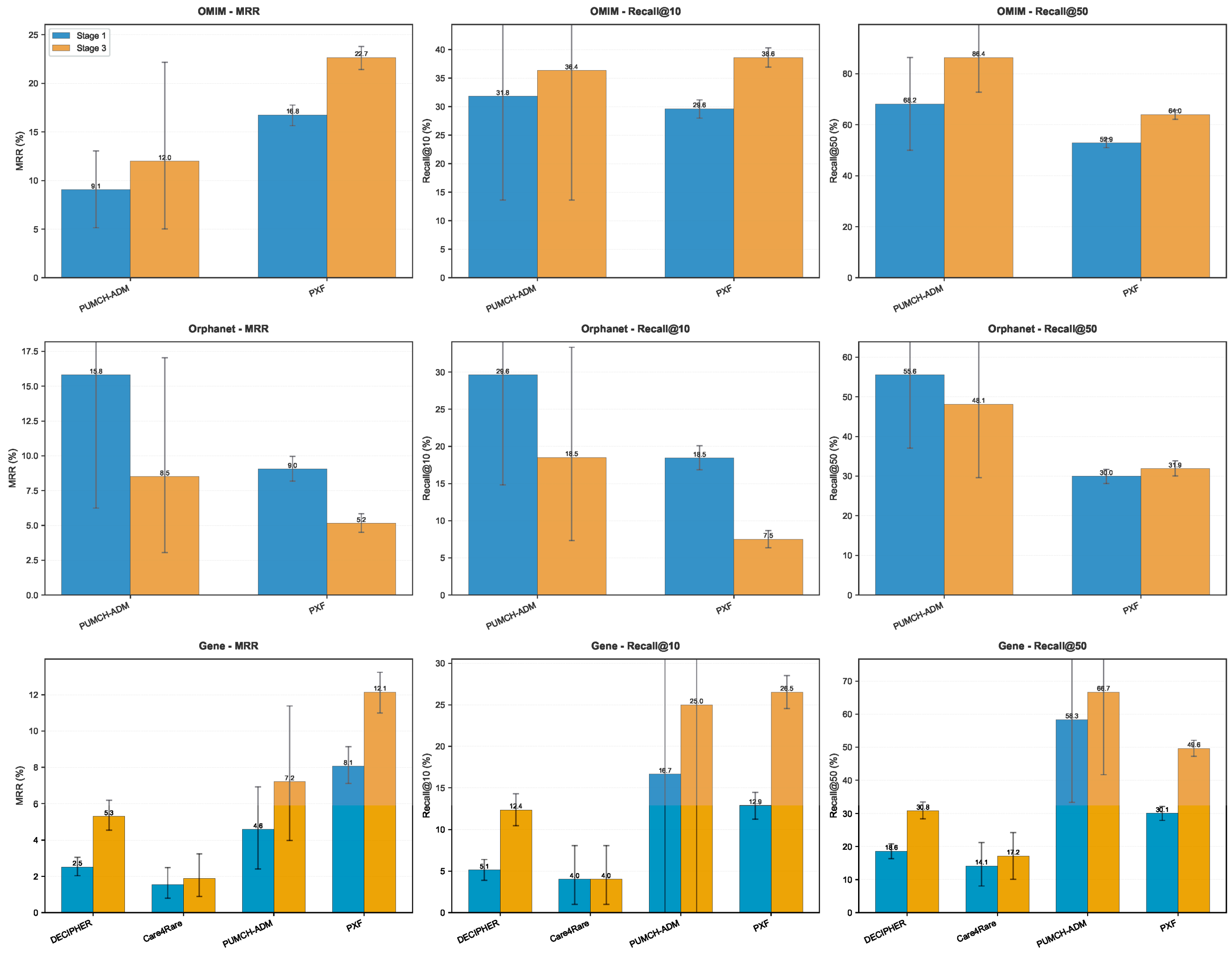


**Figure 9.** Zero-shot transfer performance across held-out rare disease corpora. Zero-shot transfer evaluation measuring model generalization to unseen rare disease cohorts without additional fine-tuning.

**Results:** Zero-shot transfer to unseen diseases and genes is substantially harder than standard retrieval, but Stage 3 again outperforms Stage 1 and embedding baselines (Fig. 9). On PXF (1,715 cases), Stage 1 achieves 8.06 MRR and Stage 3 12.15 MRR with 49.62 Recall@50 for unseen gene identification, meaning roughly half of unseen genes are retrieved within the top 50 candidates. On DECIPHER (1,204 cases), Stage 1 reaches 2.52 MRR and Stage 3 5.31 MRR, corresponding to a

110.9% relative gain, while smaller corpora (Care4Rare, PUMCH-ADM) show lower absolute performance and wider variability due to limited case counts.

The COMBINED modality (TEXT+HPO) inputs yield the best zero-shot performance in Stage 3: on PXF, combined mode attains 12.15 MRR versus 11.55 (HPO-only) and 11.23 (text-only), with some corpora (e.g. DECIPHER) exhibiting 5-10% relative gains for multimodality. Correlation analysis ($r \approx 0.87$) between corpus size and zero-shot MRR indicates that larger corpora provide more reliable and higher zero-shot performance, reflecting better coverage of phenotype patterns for the graph transformer to exploit. Despite these improvements, absolute zero-shot MRR values (≈1.9-12.2) remain well below standard retrieval (40-50 MRR on all-entity ranking in PXF), underscoring the inherent challenge of long-tail generalization.

#### S4. Embedding arithmetic and robustness

**Task:** Probe whether the embedding space supports phenotype aggregation and arithmetic operations, testing space interpretability and robustness to perturbations. Phenotype embeddings are progressively aggregated (via mean pooling), and cosine similarity to the target disease is tracked as additional phenotypes are added to the representation. The evaluation reports average similarity trajectories with bootstrap confidence intervals and the fraction of cases showing monotonically increasing similarity. Additional stress tests examine robustness to: (1) phenotype removal, measuring similarity degradation when phenotypes are dropped; (2) injected noise phenotypes, testing resilience when random unrelated HPO terms are added; and (3) ordering effects, quantifying sensitivity to the sequence in which phenotypes are aggregated. These analyses assess whether the embedding space behaves predictably under compositional operations and perturbations, which is important for model interpretability and clinical reliability.

**Results:** In contrast to the other tasks, Stage 1 performs best on embedding arithmetic and robustness tests, revealing an important trade-off between linear compositionality and graph-enhanced reasoning (Fig. 10-11). On DECIPHER, progressive phenotype aggregation yields a final disease-case cosine similarity of 0.7169 for Stage 1 versus 0.5282 for Stage 3 (35.7% advantage), and similar gaps of 29.7-37.9% are observed across PXF, Care4Rare, and PUMCH-ADM.

The monotonic increase ratio - fraction of cases where similarity increases smoothly as phenotypes are added - is 96.6% for Stage 1 versus 88.9% for Stage 3, indicating that Stage 1 provides more predictable evidence accumulation. Under 50% random phenotype removal, Stage 1's similarity drops from 0.7169 to 0.6841 (4.6% degradation), whereas Stage 3 drops 8.4%; with 30% noisy phenotypes, Stage 1 degrades only 1.1% and Stage 3 2.0%, while both models are effectively invariant to phenotype ordering (std ≈0). Compared with MedEmbed and SapBERT, Stage 1 achieves higher final similarity (0.7169 vs 0.6366 and 0.5135, respectively), although MedEmbed attains slightly higher monotonicity (99.4% vs 98.7%), suggesting different geometry trade-offs between absolute similarity and smoothness.

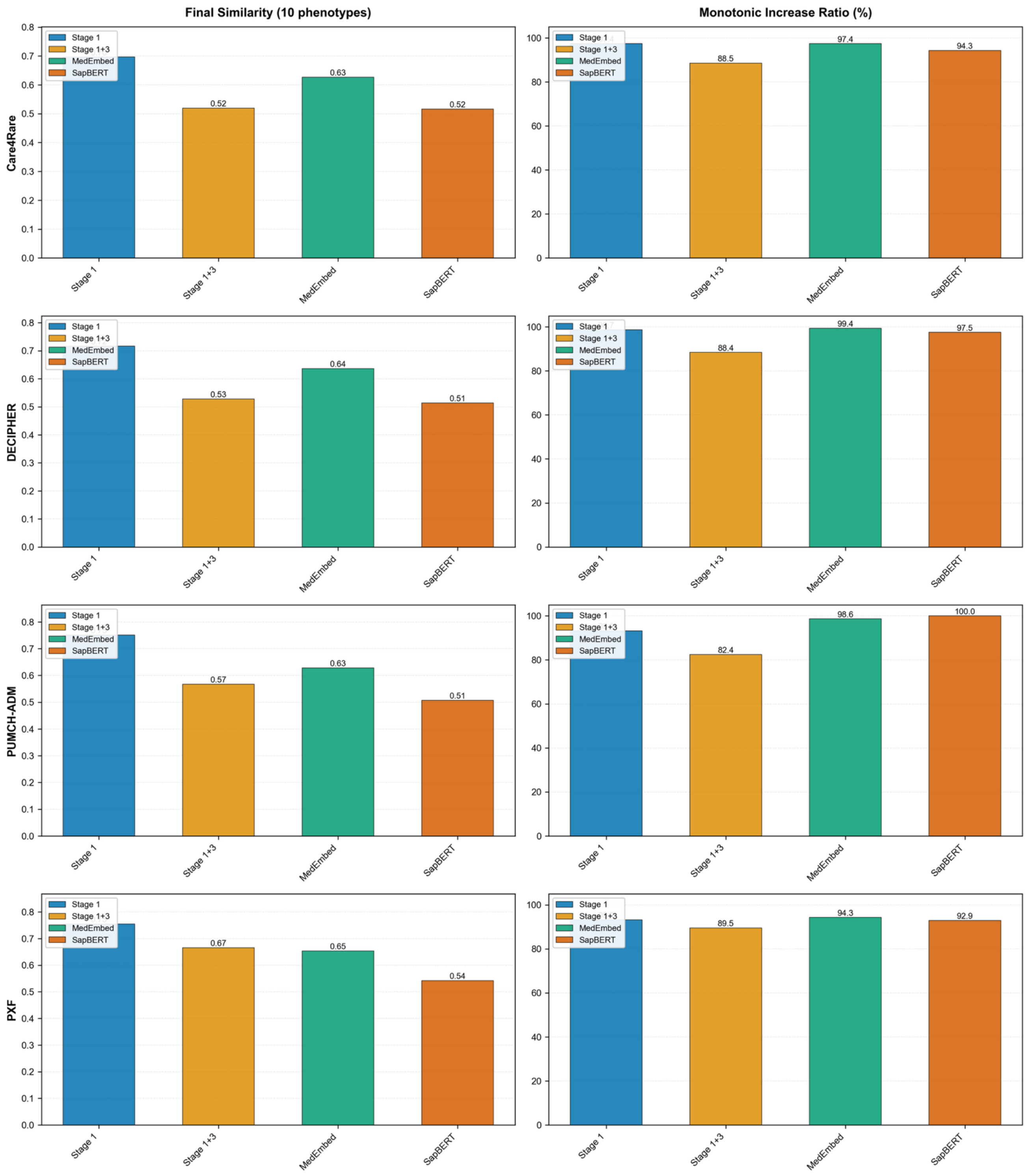


**Figure 10.** Similarity scores and monotonic increase ratio across a selection of corpora and models

Across evaluations, these findings justify task-dependent model selection: Stage 1 for workflows requiring interpretable, incremental phenotyping and robustness to missing or noisy data, and Stage 3 (optionally Stage 2+3 for EHR) for final high-accuracy ranking in diagnosis, gene identification, differential ranking, phenotype inference, and zero-shot scenarios. This stage-wise trade-off maps naturally to clinical practice: Stage 1 supports early, incremental evidence accumulation when phenotyping is sparse and evolving, whereas Stage 3 is best suited for final high-confidence ranking once richer phenotypic and longitudinal data are available.

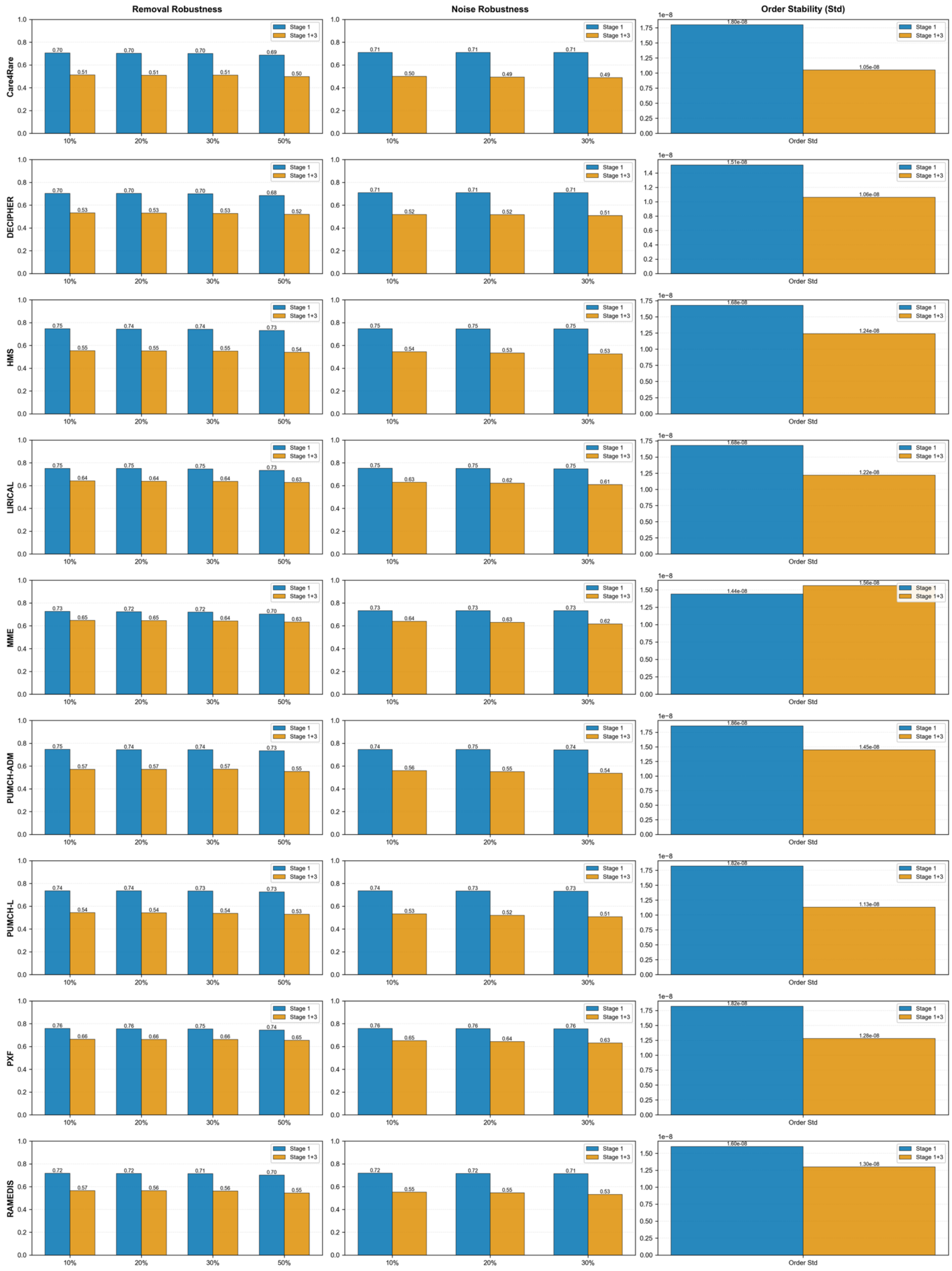


**Figure 11.** Comparative overview of robustness and order stability across all corpora

### Supplementary Methods

**Stage 1: Ontology-aware contrastive learning**

Stage 1 was optimized with a bidirectional InfoNCE loss over cosine similarities between anchor-positive pairs versus negatives at a temperature of 0.05, applied symmetrically in both directions - see Eq 1. Training used 32 samples per batch, AdamW (weight decay 0.01) with differential learning rates with $1\times10^{-5}$ for encoder and $2\times10^{-5}$ for projection head, automatic mixed precision optimization across 30 epochs with early stopping on 70/15/15 train-validation-test split balanced across pair types.

$$L_{\text{InfoNCE}} = -\log\left(\frac{\exp(\text{sim}(\mathbf{e}_\text{a}, \mathbf{e}_\text{p}) / \tau)}{\exp(\text{sim}(\mathbf{e}_\text{a}, \mathbf{e}_\text{p}) / \tau) + \sum_{i=1}^{K} \exp(\text{sim}(\mathbf{e}_\text{a}, \mathbf{e}_{n_i}) / \tau)}\right) \quad \text{Eq .1}$$

where $e_a$, $e_p$ and $e_{ni}$ are anchor, positive, and negative embeddings respectively, *sim* denotes cosine similarity, *tau=0.05* is the temperature parameter, and *N=3* is the number of hard negatives per anchor. Bidirectional loss computes both forward (*a* → *p*) and reverse (*p* → *a*) directions with equal weighting.

**Stage 2: Clinical text alignment with SNOMED bridge**

Stage 2 extends the embedding space to clinical concepts via a dual-encoder architecture that aligns clinical text to the fixed Stage 1 entity space, mediated by SNOMED CT concepts. A clinical transformer encoder with its own projection head maps clinical concepts represented as text into 512-dimensional L2-normalized embeddings, while a frozen copy of the Stage 1 model encodes disease, gene, phenotype, and ontology-derived mention strings. We used GatorTron-base, a 345M parameter clinical language model pretrained on 90 billion words of clinical text.

SNOMED CT concepts are initialized by averaging Stage 1 embeddings of mapped HPO and Orphanet entities, refined with hierarchical smoothing, and then passed through a projection network to produce 512-dimensional code embeddings that bridge clinical codes to the ontology-derived space. Stage 2 training proceeds in three phases:

- SNOMED anchoring, which fits the SNOMED projection network using mean-squared error against Stage 1 targets – Eq. 2;
- clinical alignment, which trains the clinical encoder with a bidirectional in-batch contrastive loss between clinical segments and linked entities while keeping Stage 1 frozen – Eq 3;
- joint fine-tuning, which updates the clinical and SNOMED encoders, and optionally the Stage 1 projection layers, under a composite loss combining alignment, code-bridge, and hierarchical consistency terms with tuned weighting coefficients – Eq 4.

$$\mathcal{L}_{\text{SNOMED}} = \frac{1}{K} \sum_{i=1}^{K} \| e_{\text{SNOMED},i} - e_{\text{target},\ i} \| \quad \text{Eq .2}$$

$$\mathcal{L}_{\text{align}} = \mathcal{L}_{\text{IBN}}(e_c, e_m) + \mathcal{L}_{\text{IBN}}(e_m, e_c) \qquad \text{Eq .3}$$

where $L_{IBN}$ is in-batch negatives contrastive loss treating all other samples in the batch as negatives.

$$\mathcal{L}_{\text{joint}} = \mathcal{L}_{\text{align}} + \lambda_1 \mathcal{L}_{\text{bridge}} + \lambda_2 \mathcal{L}_{\text{overlap}} \qquad \text{Eq .4}$$

where:

- $L_{bridge}$ = cross-modal loss between clinical text and SNOMED codes
- $L_{overlap}$ = hierarchical consistency loss for SNOMED parent-child relationships
- $\lambda_1 = 0.3$, $\lambda_2 = 0.0$ (tunable hyperparameters)

Hyperparameters include smaller learning rates for shared components, entity-type-specific weighting (higher for diseases than genes and phenotypes), 4,096-token windows with overlap, and early stopping based on mean reciprocal rank for entity retrieval from clinical text.

**Stage 3: Heterogeneous graph transformer enhancement**

Graph construction:

Node Types (6 types, 52,847 nodes total):

- Disease (16,005 nodes): OMIM and Orphanet rare disease entities
- Phenotype (16,725 nodes): HPO terms
- Gene (19,198 nodes): HGNC protein-coding genes
- Gene Ontology (28,547 nodes): GO biological process, molecular function, and cellular component terms
- Pathway (2,372 nodes): Reactome pathway annotations
- Anatomy (4,000 nodes): UBERON anatomical structures

Edge Types (17 types, 1,247,563 edges total)

Disease-centric edges:

- Disease → Phenotype (has_phenotype): 549,241 edges from OMIM-HPO and Orphanet-HPO
- Disease → Gene (has_gene): 9,465 edges from OMIM-HGNC and Orphanet-HGNC
- Disease ↔ Disease (similar_to): 23,847 edges from Resnik semantic similarity >0.7
- Disease ↔ Disease (mapped_to): 4,128 edges from OMIM-Orphanet cross-references

Gene-centric edges:

- Gene → Phenotype (has_phenotype): 87,134 edges from Gene2Phenotype

- Gene → Pathway (has_pathway): 41,256 edges from Reactome
- Gene → Anatomy (associated_with): 19,847 edges from GTEx tissue expression
- Gene → GO (enables, involved_in, located_in, acts_upstream_of_or_within): 287,451 edges from Gene Ontology annotations

Hierarchy edges:

- Phenotype → Phenotype (is_a): 18,947 edges from HPO hierarchy
- GO → GO (is_a, part_of, regulates, positively_regulates, negatively_regulates): 124,578 edges from GO hierarchy
- Anatomy → Anatomy (is_a, part_of): 81,669 edges from UBERON hierarchy

The HGT performs type-specific multi-head attention and aggregation over neighbours, followed by layer normalization and a learnable residual connection that mixes normalized HGT outputs with normalized input embeddings via a scalar parameter learned per node type. Training uses an InfoNCE-style loss over triplets sampled per node type, where anchors and positives are linked nodes and negatives are unconnected nodes of the same type, with weighted sampling emphasizing edges with high information content (e.g. rare phenotypes) – Eq. 5. A single HGT layer with four attention heads was chosen based on validation to avoid over-smoothing, and the Stage 1 encoder remains frozen to preserve semantic content while the graph layer learns structural refinements. Critically - we normalize embeddings before computing the residual connection to prevent gradient collapse – Eq. 6.

$$\boldsymbol{h}_t^{(l+1)} = \text{Aggregate}_{s \in N(t)} \left[ \text{Attention}^{(l)} \left( \boldsymbol{h}_t^{(l)}, \boldsymbol{h}_s^{(l)}, \tau(\text{t}), \tau(\text{s}), \varphi(e_{ts}) \right) \right] \quad \text{Eq .5}$$

where:

- $h_t^{(l)}$ – is the embedding of target node *t* at layer *l*
- *N(t)* – is the set of neighbours of *t*
- $\tau(.)$ – is the node type function
- $\varphi(.)$ – is the edge type function
- attention is type-specific multi-head attention with 4 heads

$$\boldsymbol{e}_{\text{enhanced}} = \text{Normalize}\left( \alpha \cdot \text{Normalize}(\boldsymbol{h}_{\text{HGT}}) + (1-\alpha) \cdot \text{Normalize}(\boldsymbol{h}_{\text{input}}) \right) \quad \text{Eq .6}$$

### Supplementary Figures

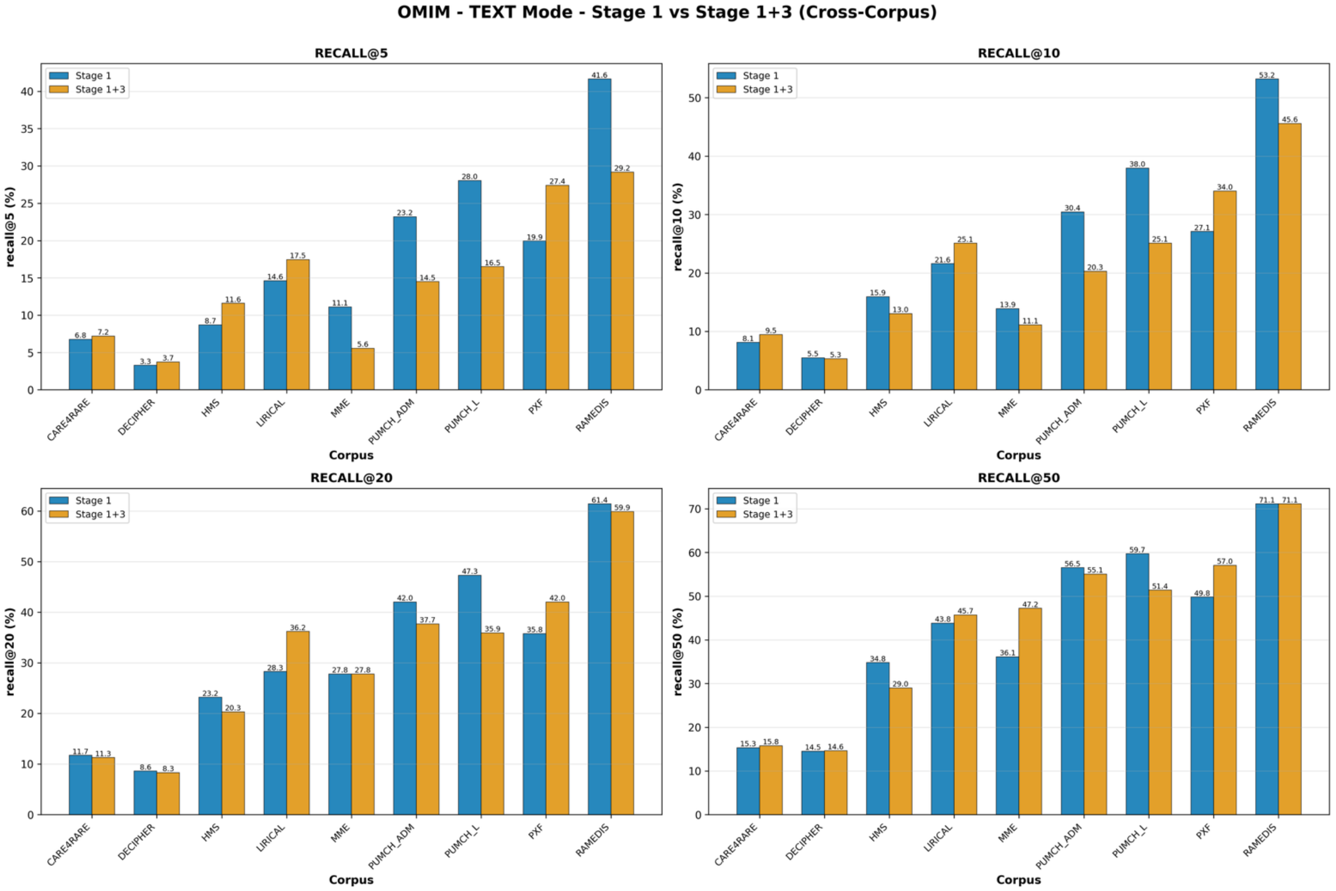


**Figure S1.** Disease ranking, cross-corpus comparison on OMIM using the TEXT modality

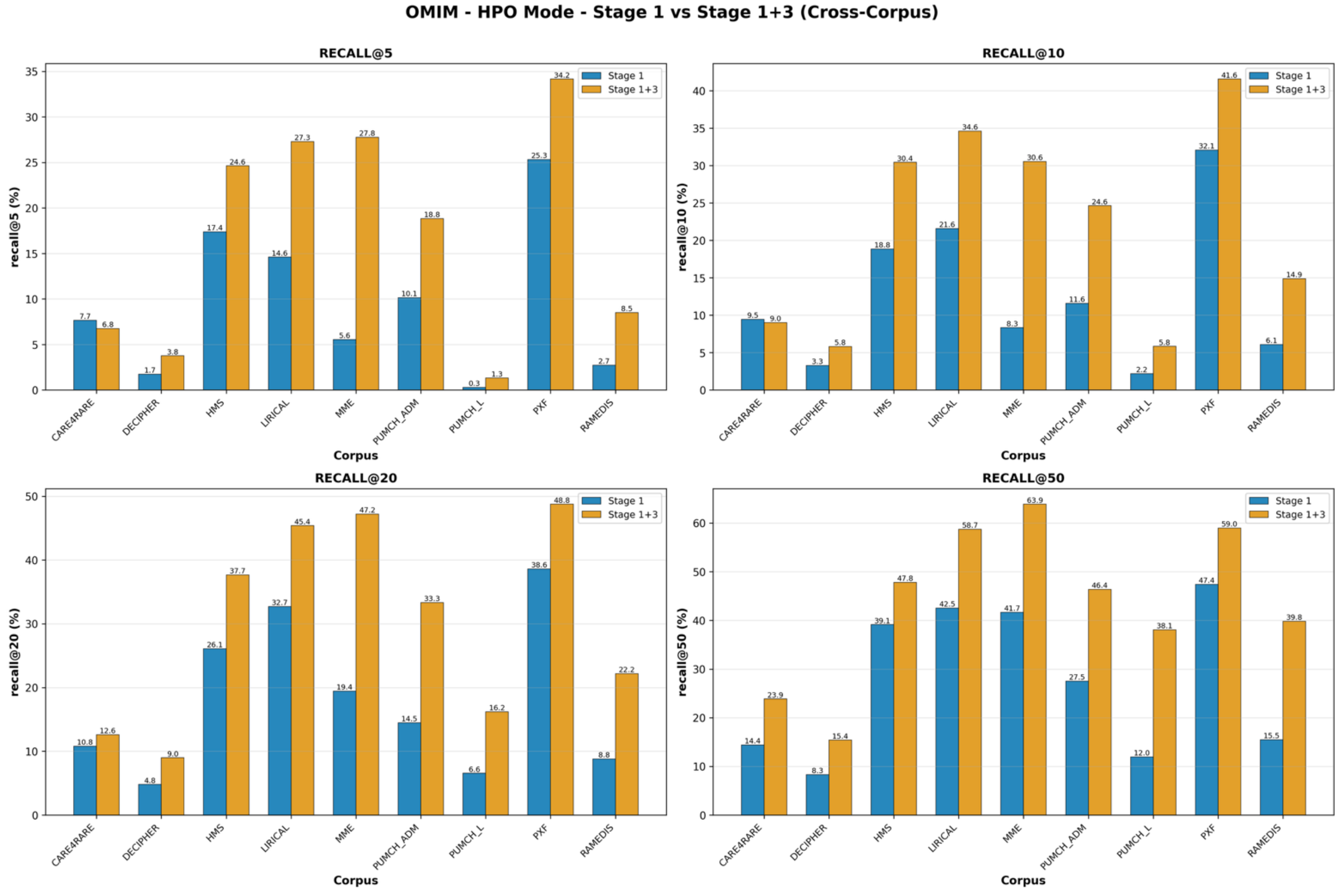


**Figure S2.** Disease ranking, cross-corpus comparison on OMIM using the HPO modality

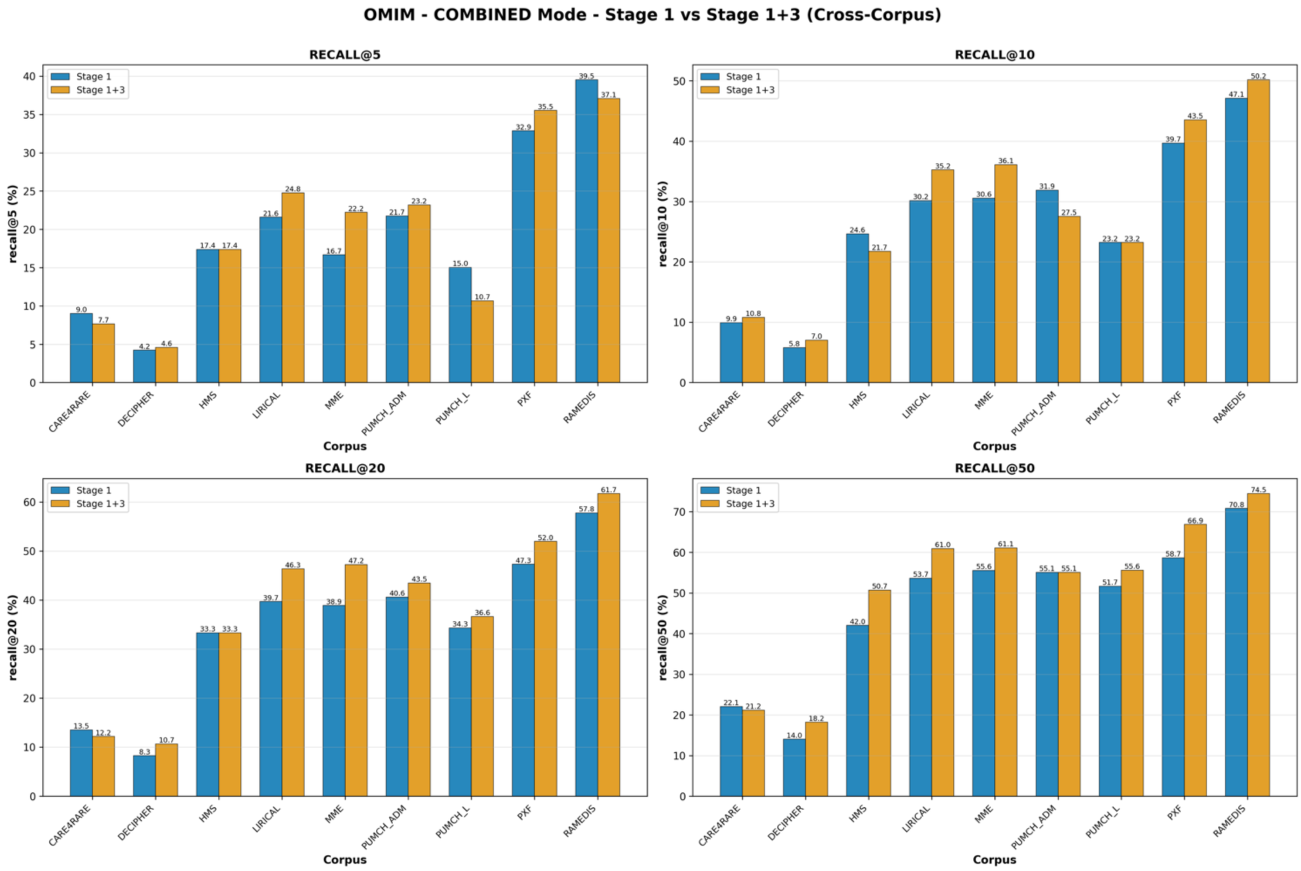


**Figure S3.** Disease ranking, cross-corpus comparison on OMIM using the COMBINED modality

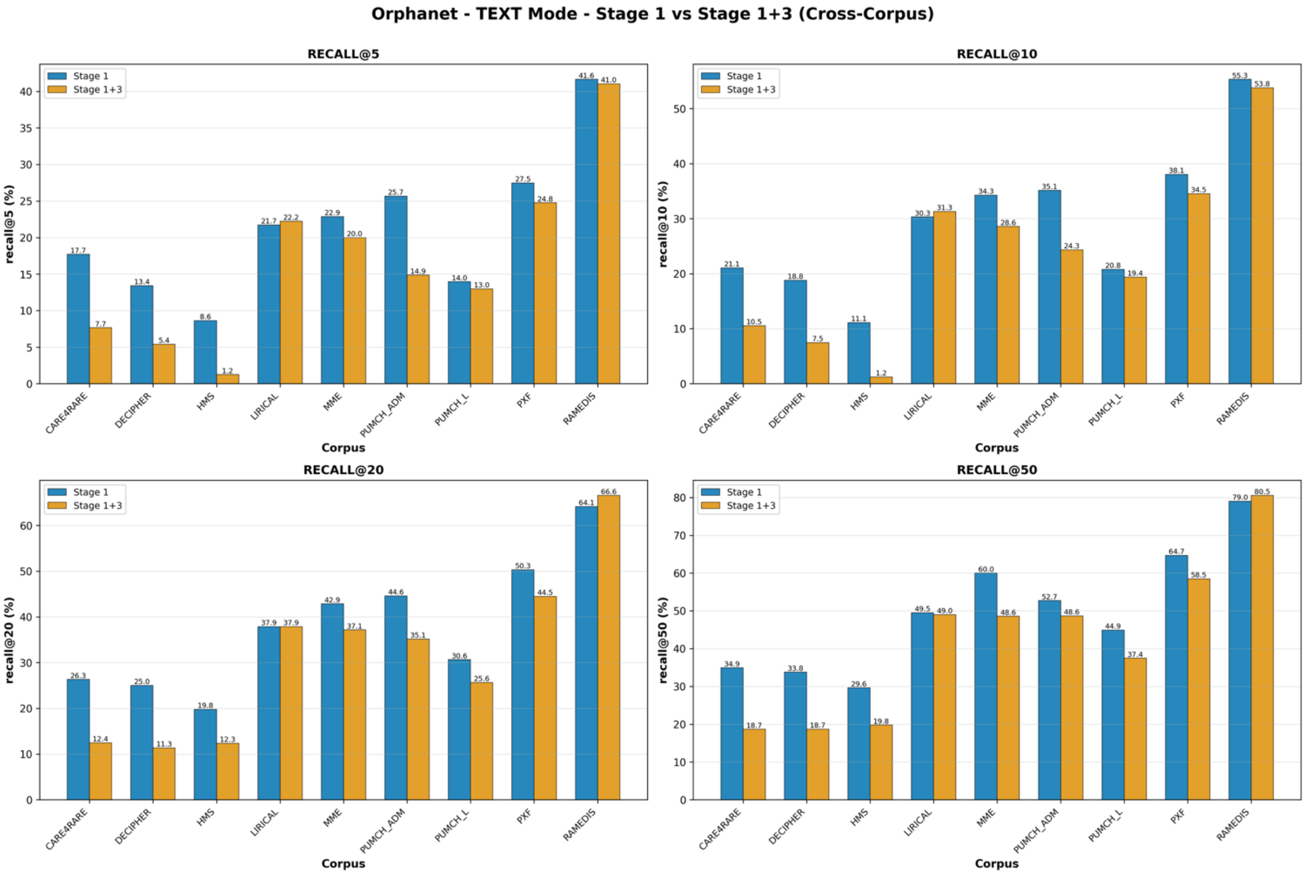


**Figure S4.** Disease ranking, cross-corpus comparison on Orphanet using the TEXT modality

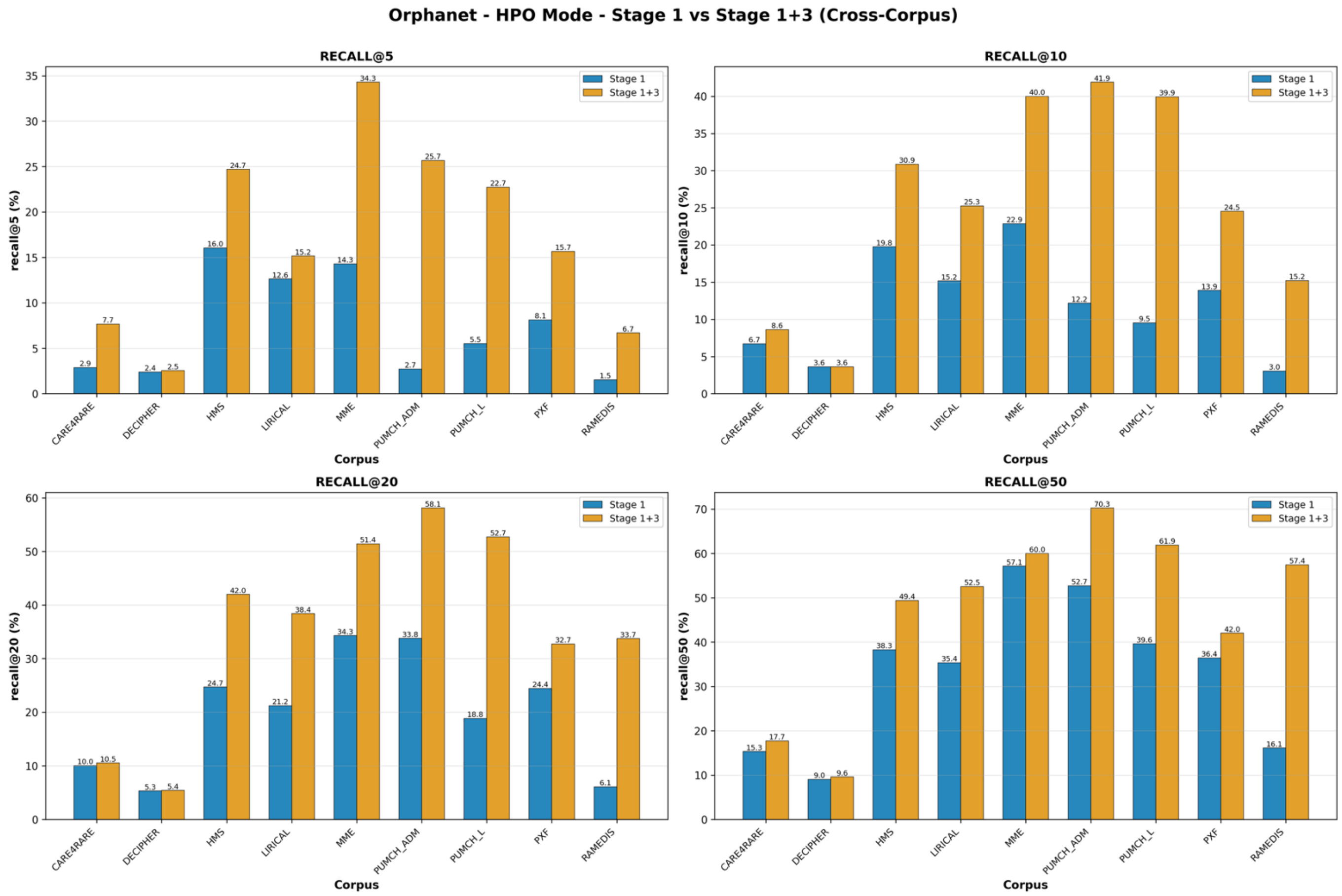


**Figure S5.** Disease ranking, cross-corpus comparison on Orphanet using the HPO modality

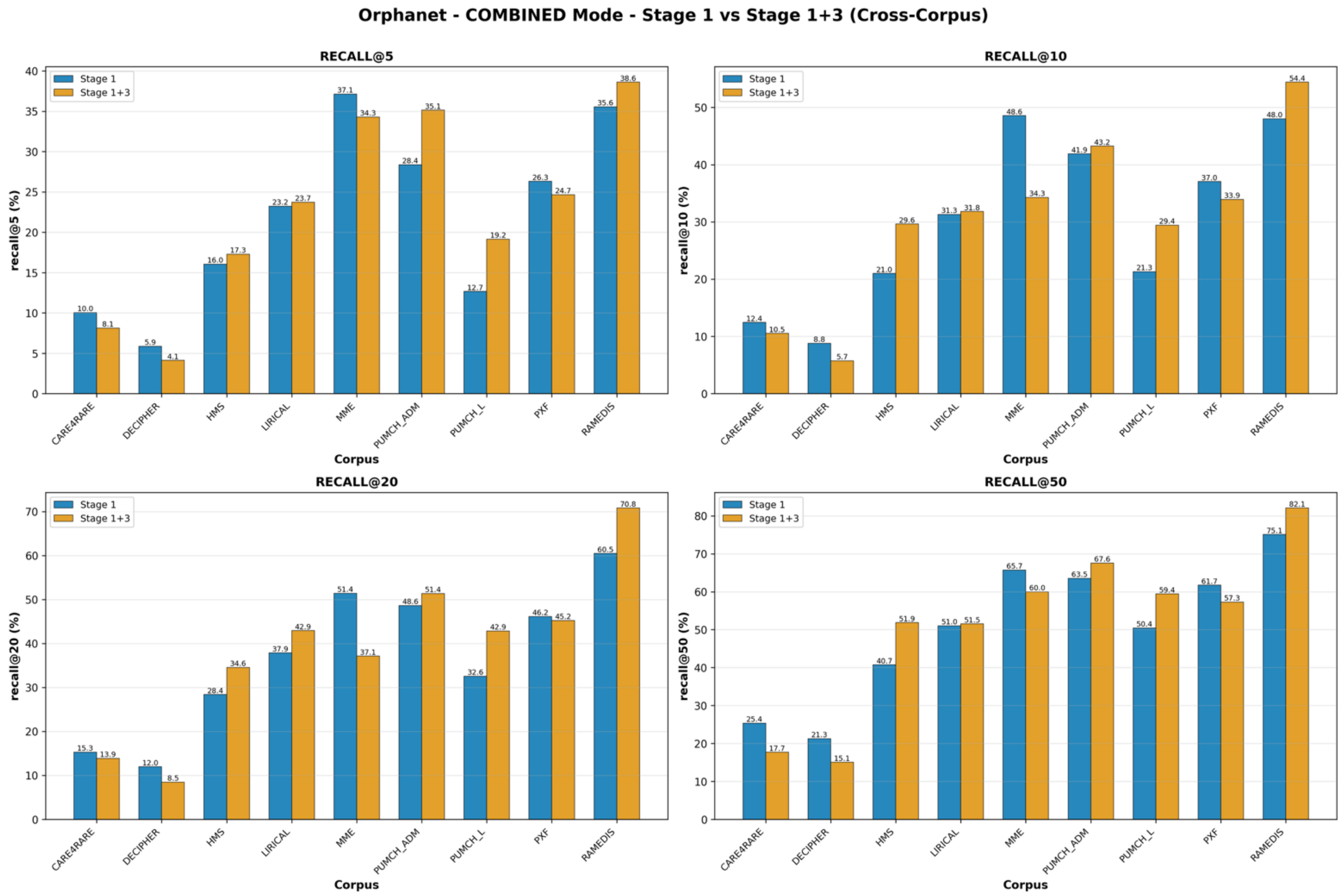


**Figure S6.** Disease ranking, cross-corpus comparison on Orphanet using the COMBINED modality

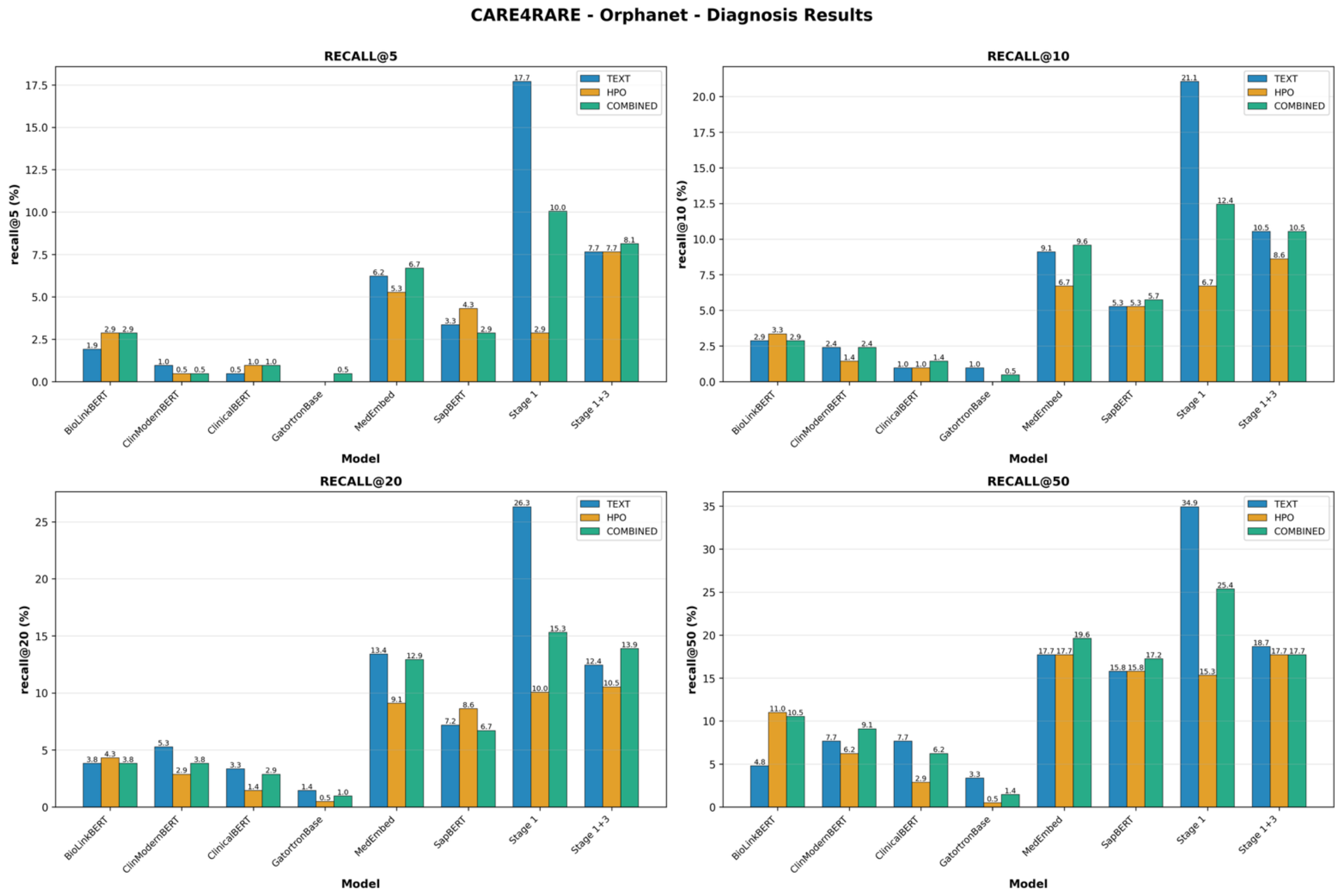


**Figure S7.** Orphanet-based disease ranking comparison across all modalities with a focus on the Care4Rare corpus

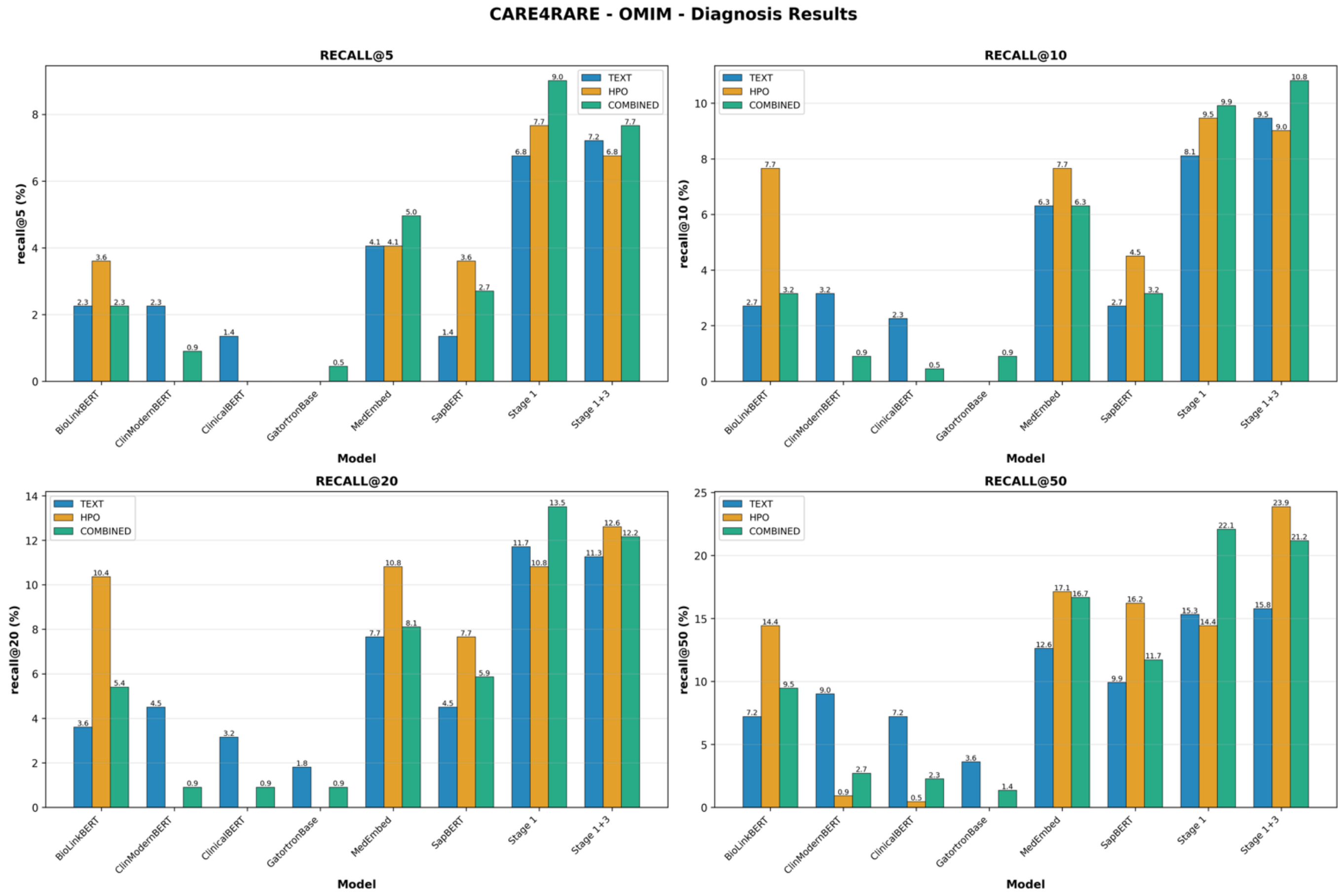


**Figure S8.** OMIM-based disease ranking comparison across all modalities with a focus on the Care4Rare corpus

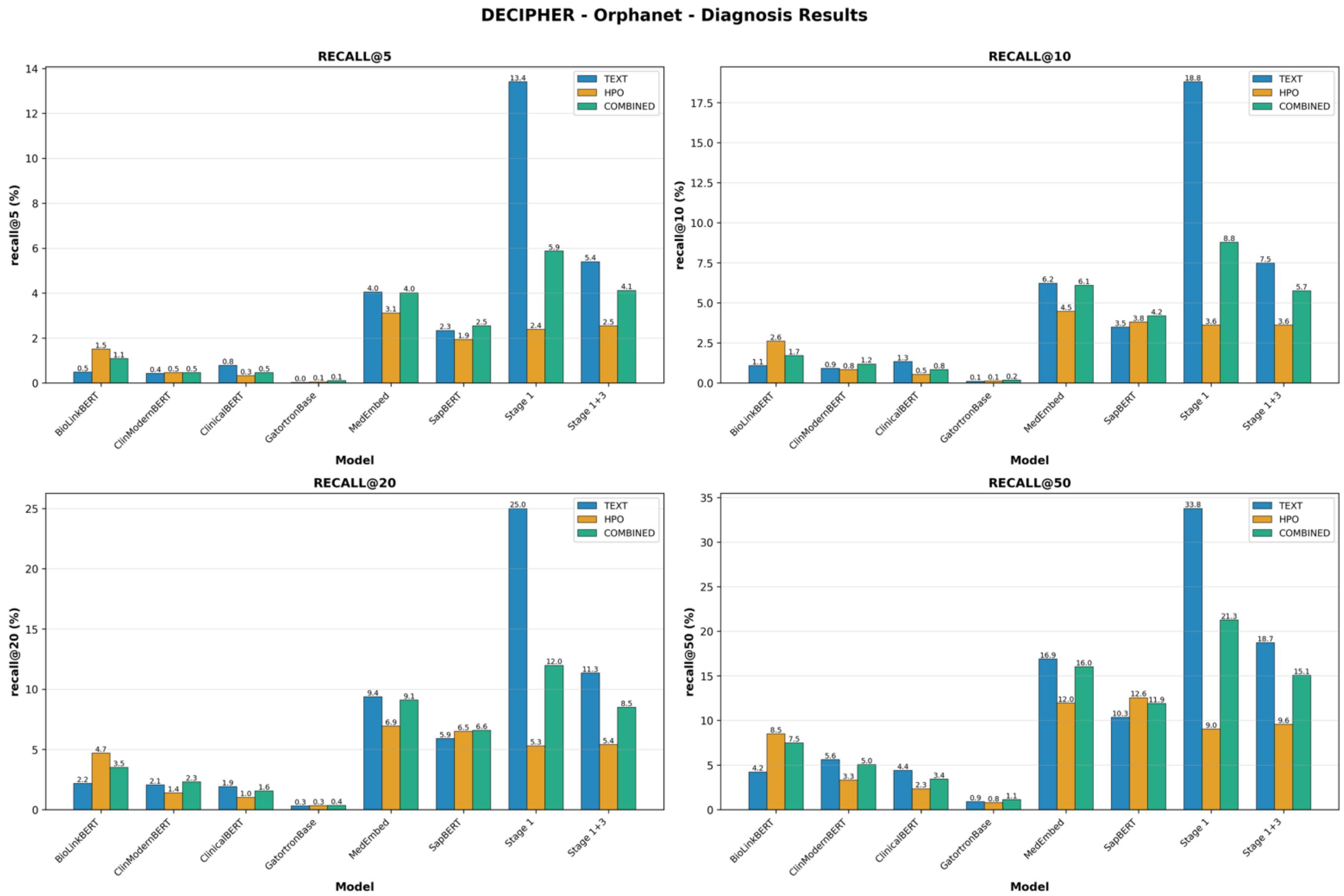


**Figure S9.** Orphanet-based disease ranking comparison across all modalities with a focus on the DECIPHER corpus

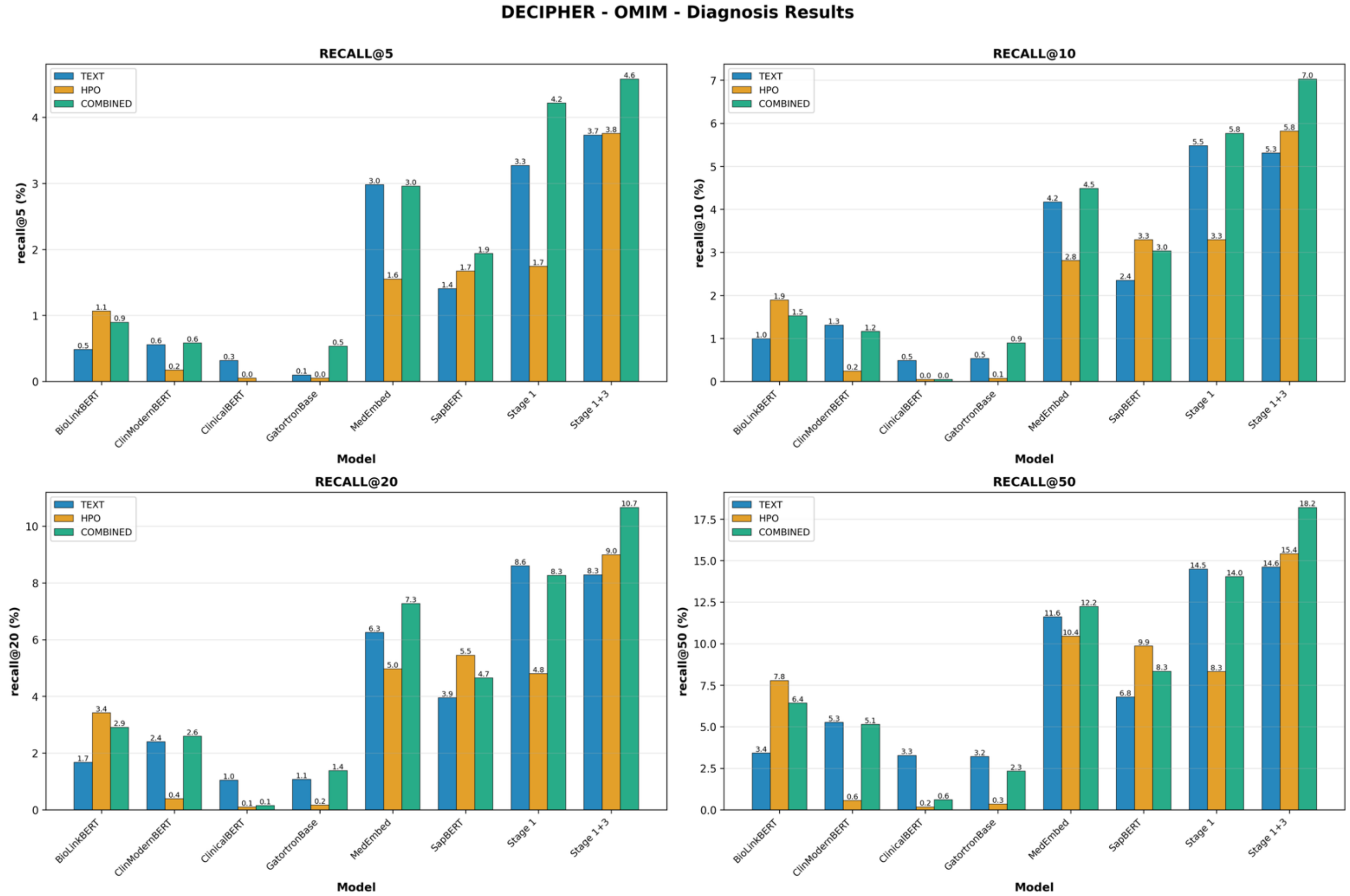


**Figure S10.** OMIM-based disease ranking comparison across all modalities with a focus on the DECIPHER corpus

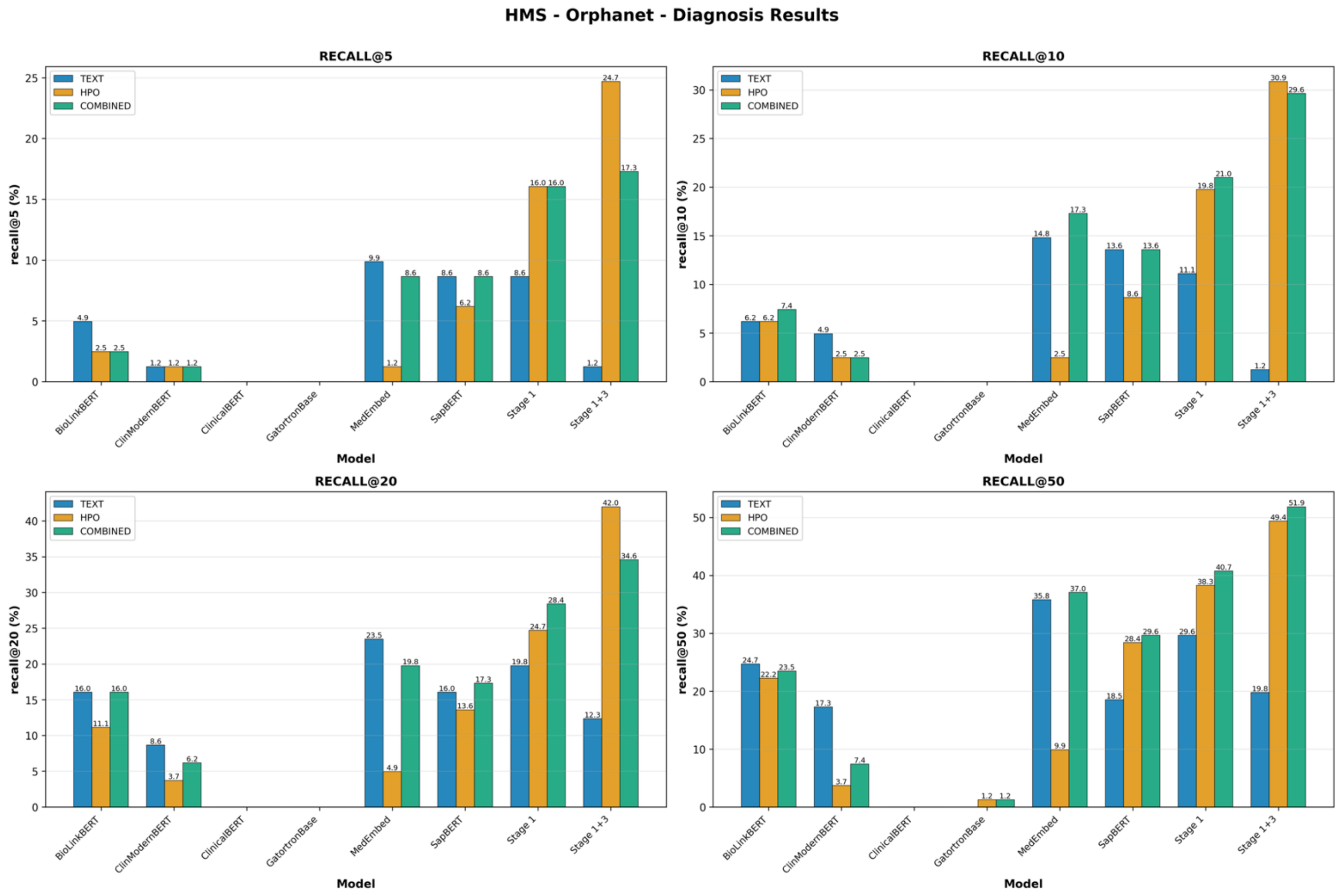


**Figure S11.** Orphanet-based disease ranking comparison across all modalities with a focus on the HMS corpus

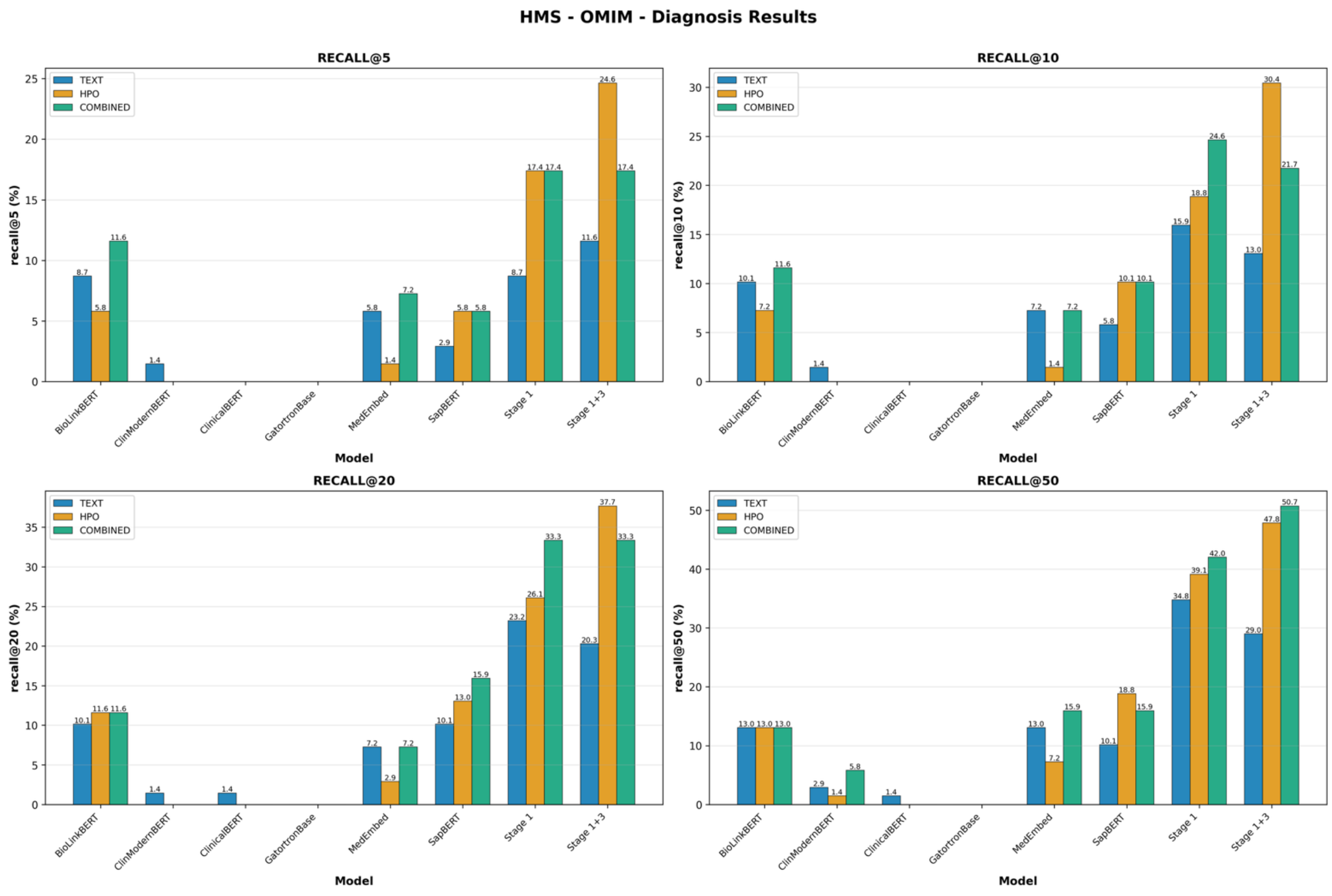


**Figure S12.** OMIM-based disease ranking comparison across all modalities with a focus on the HMS corpus

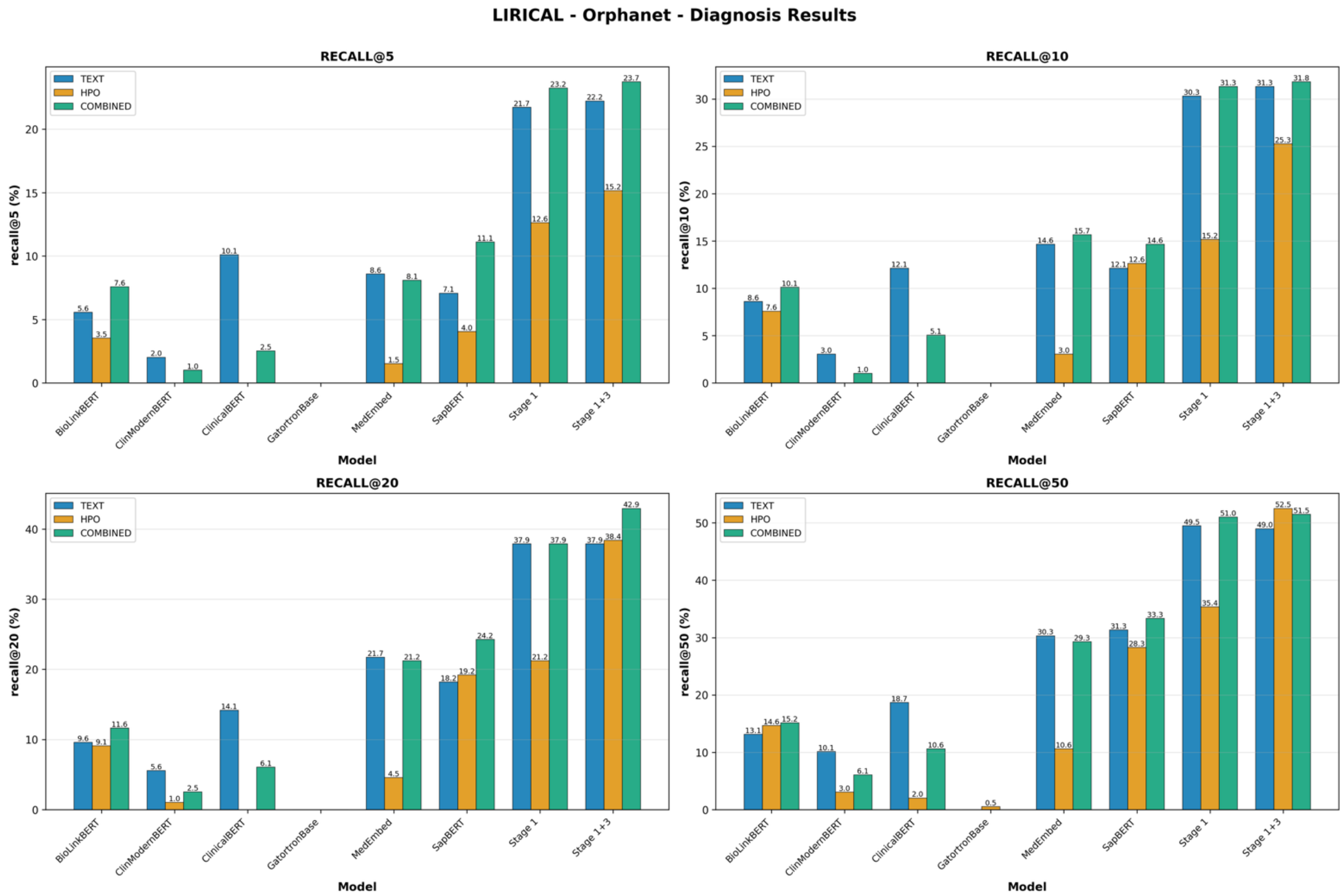


**Figure S13.** Orphanet-based disease ranking comparison across all modalities with a focus on the LIRICAL corpus

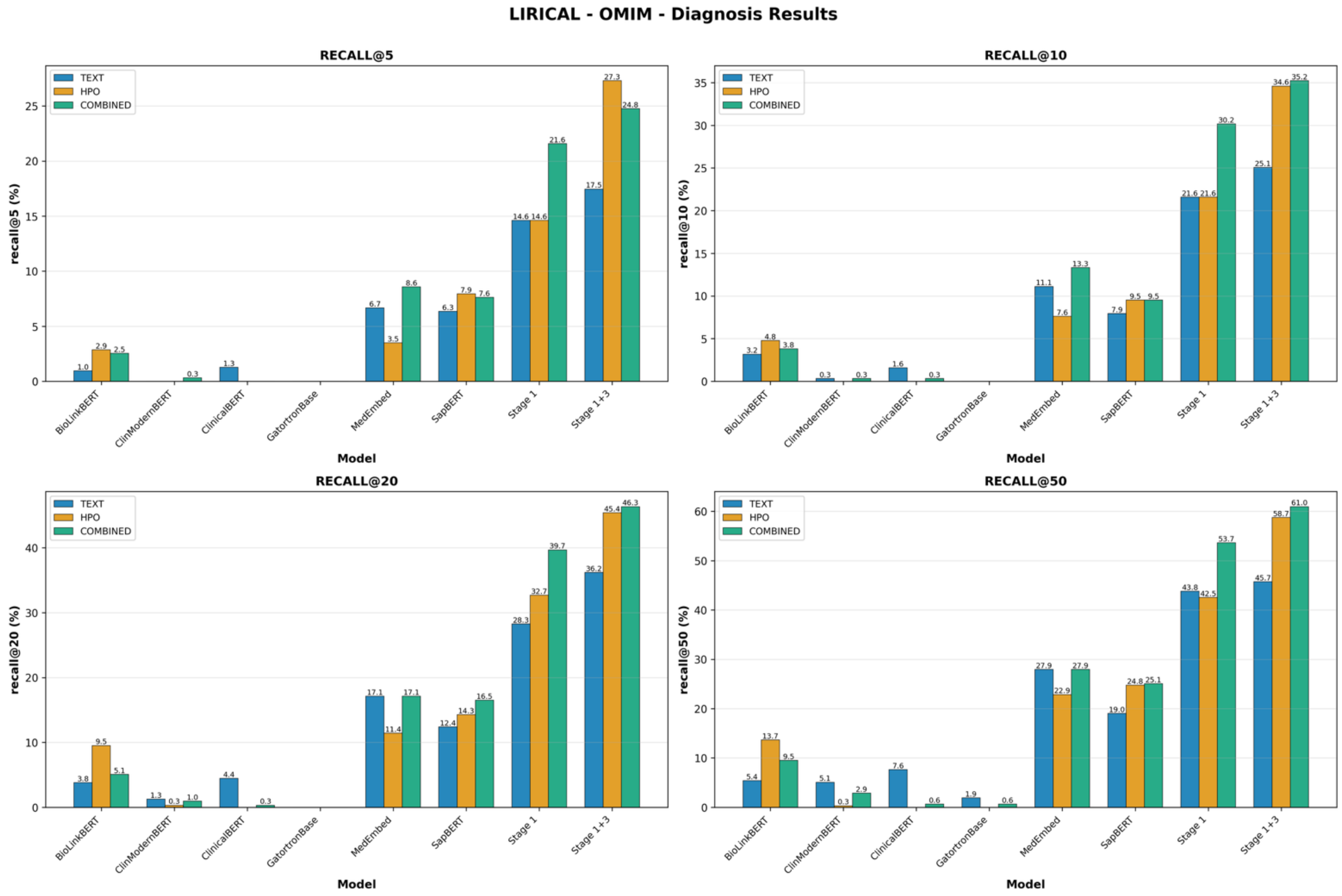


**Figure S14.** OMIM-based disease ranking comparison across all modalities with a focus on the LIRICAL corpus

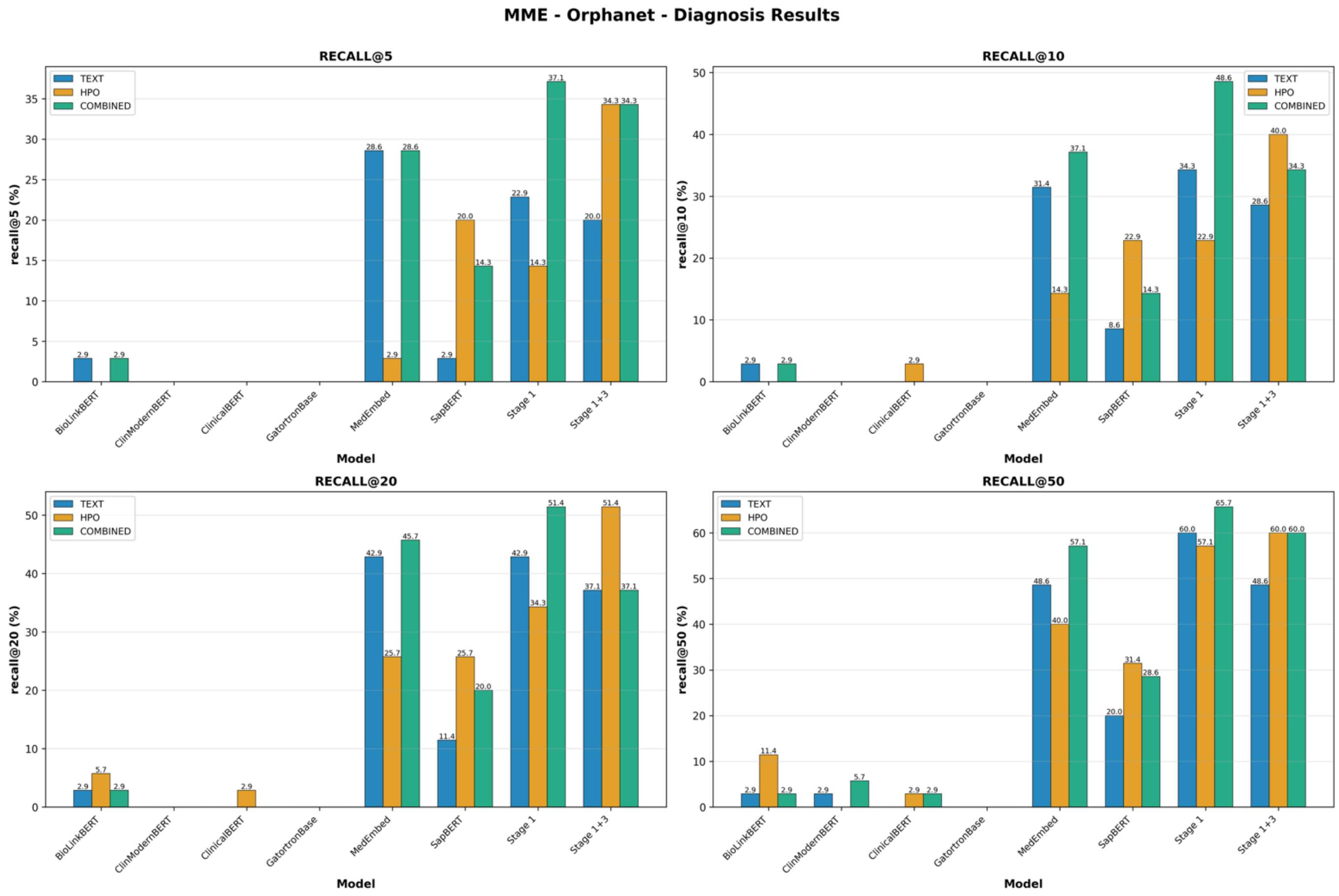


**Figure S15.** Orphanet-based disease ranking comparison across all modalities with a focus on the MME corpus

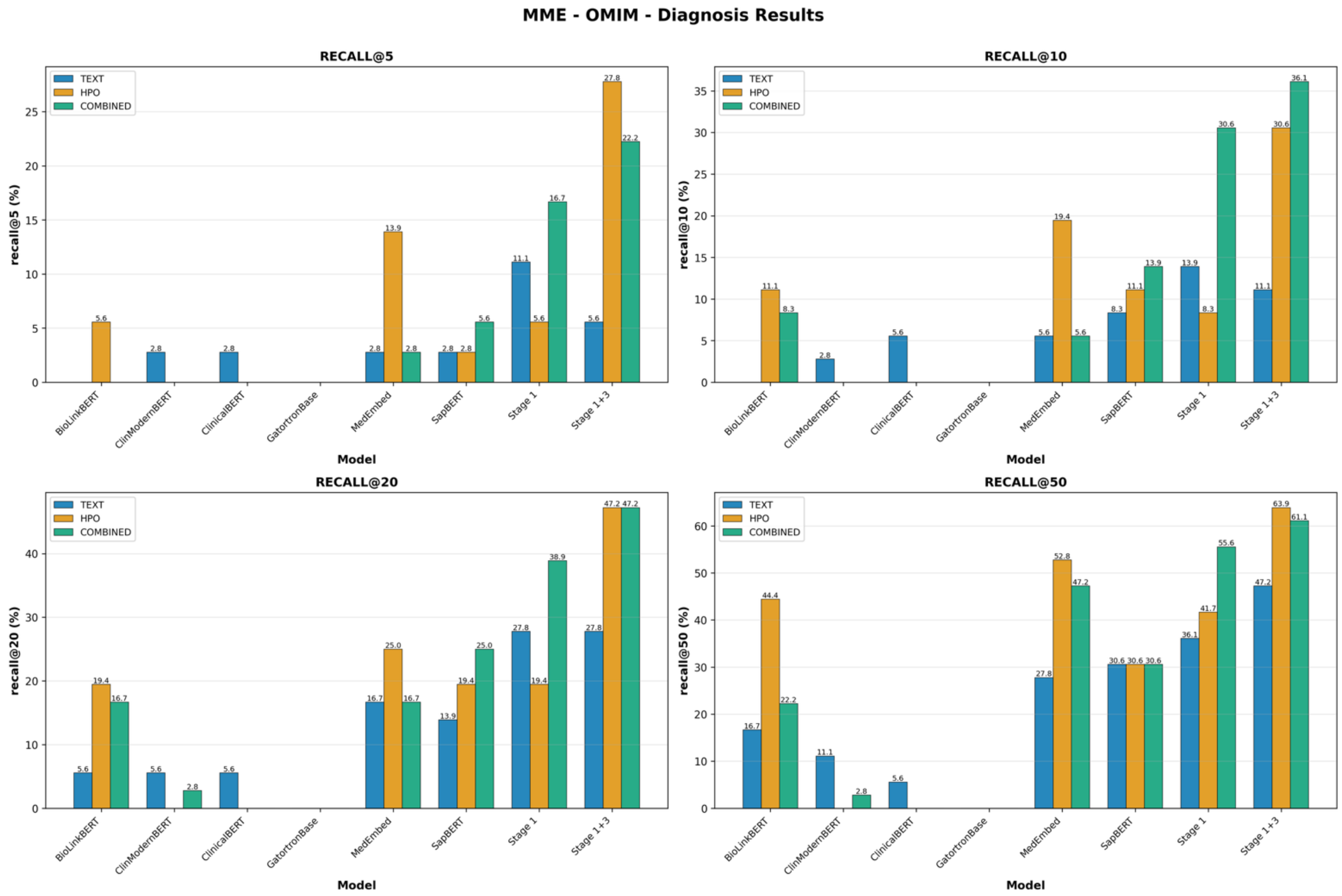


**Figure S16.** OMIM-based disease ranking comparison across all modalities with a focus on the MME corpus

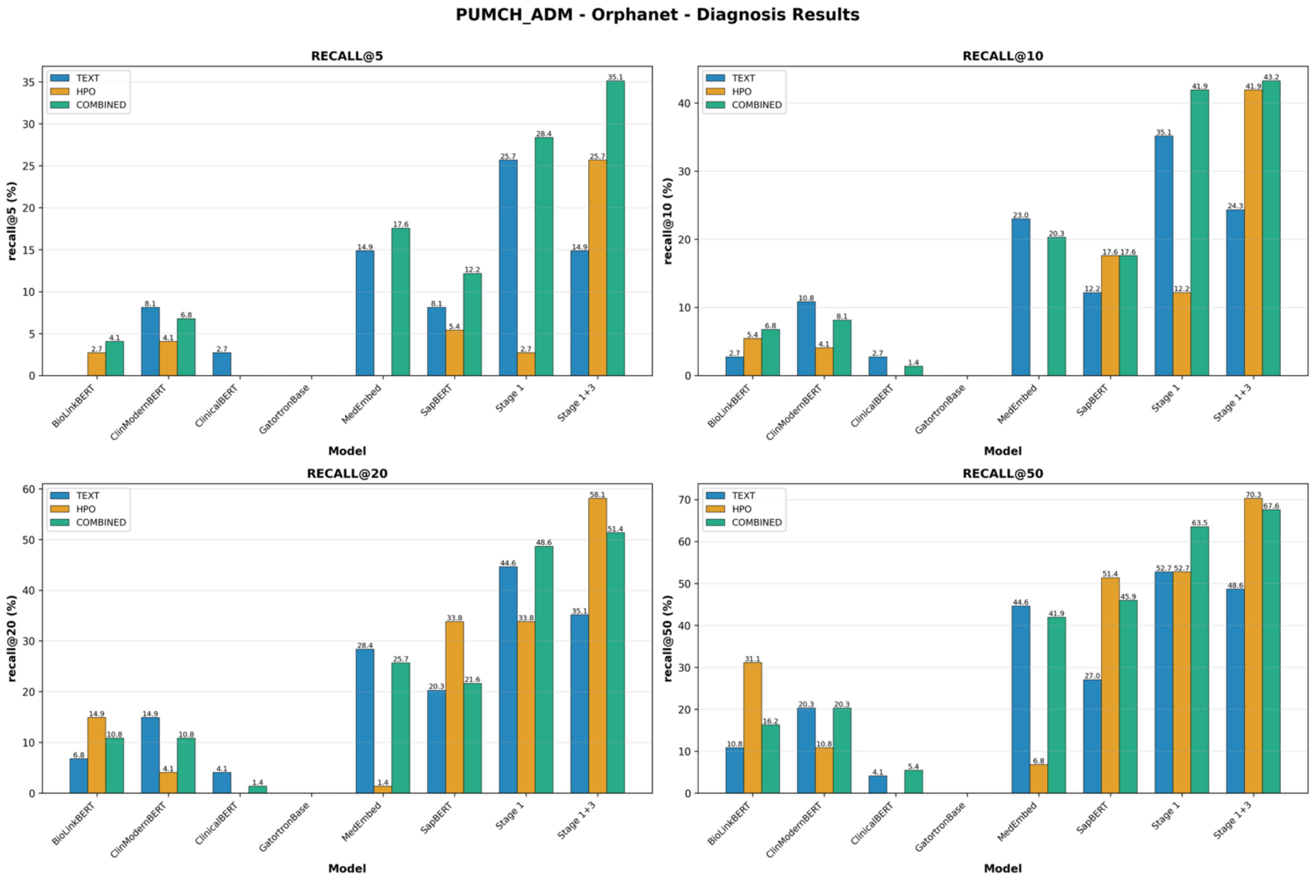


**Figure S17.** Orphanet-based disease ranking comparison across all modalities with a focus on the PUMCH-ADM corpus

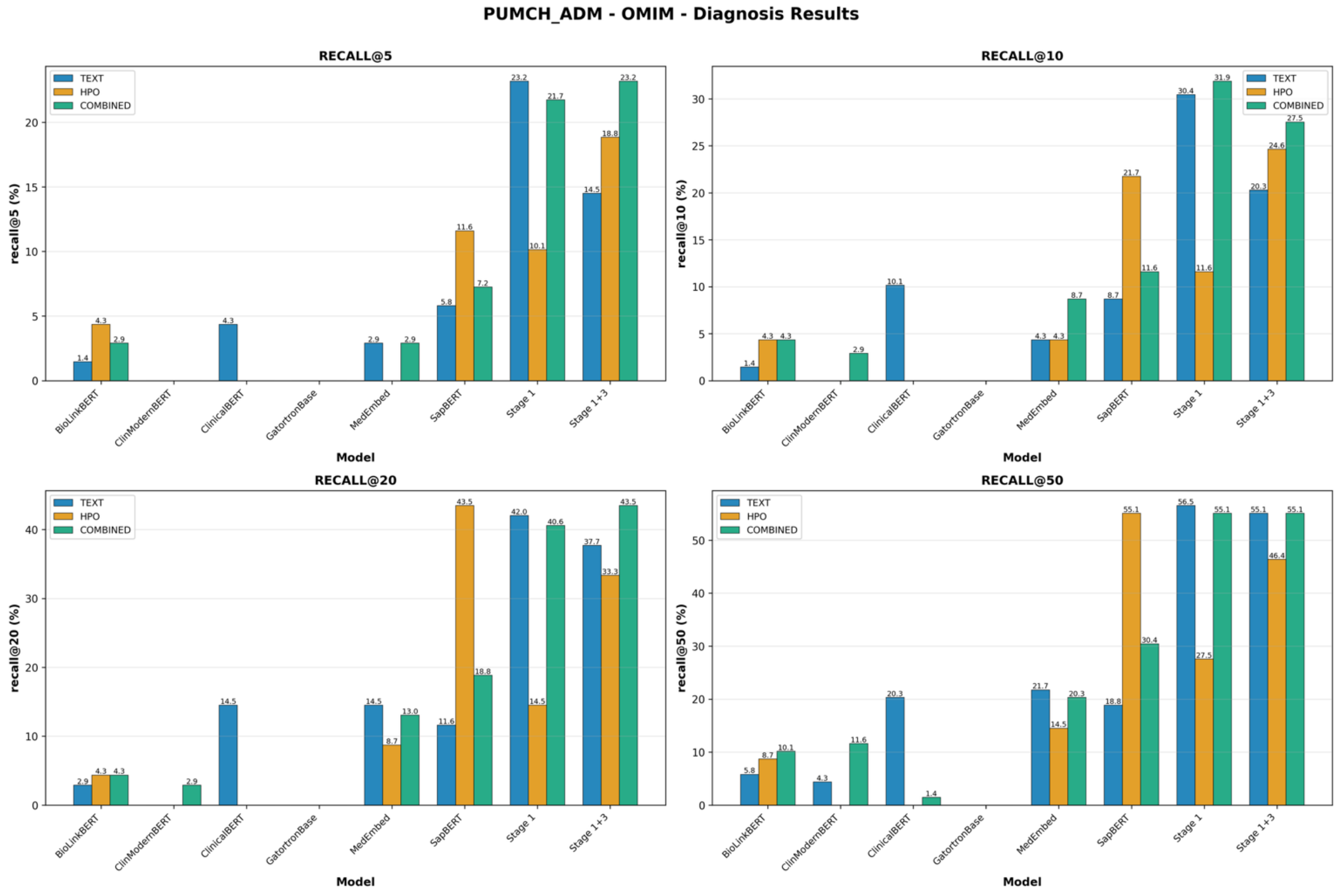


**Figure S18.** OMIM-based disease ranking comparison across all modalities with a focus on the PUMCH-ADM corpus

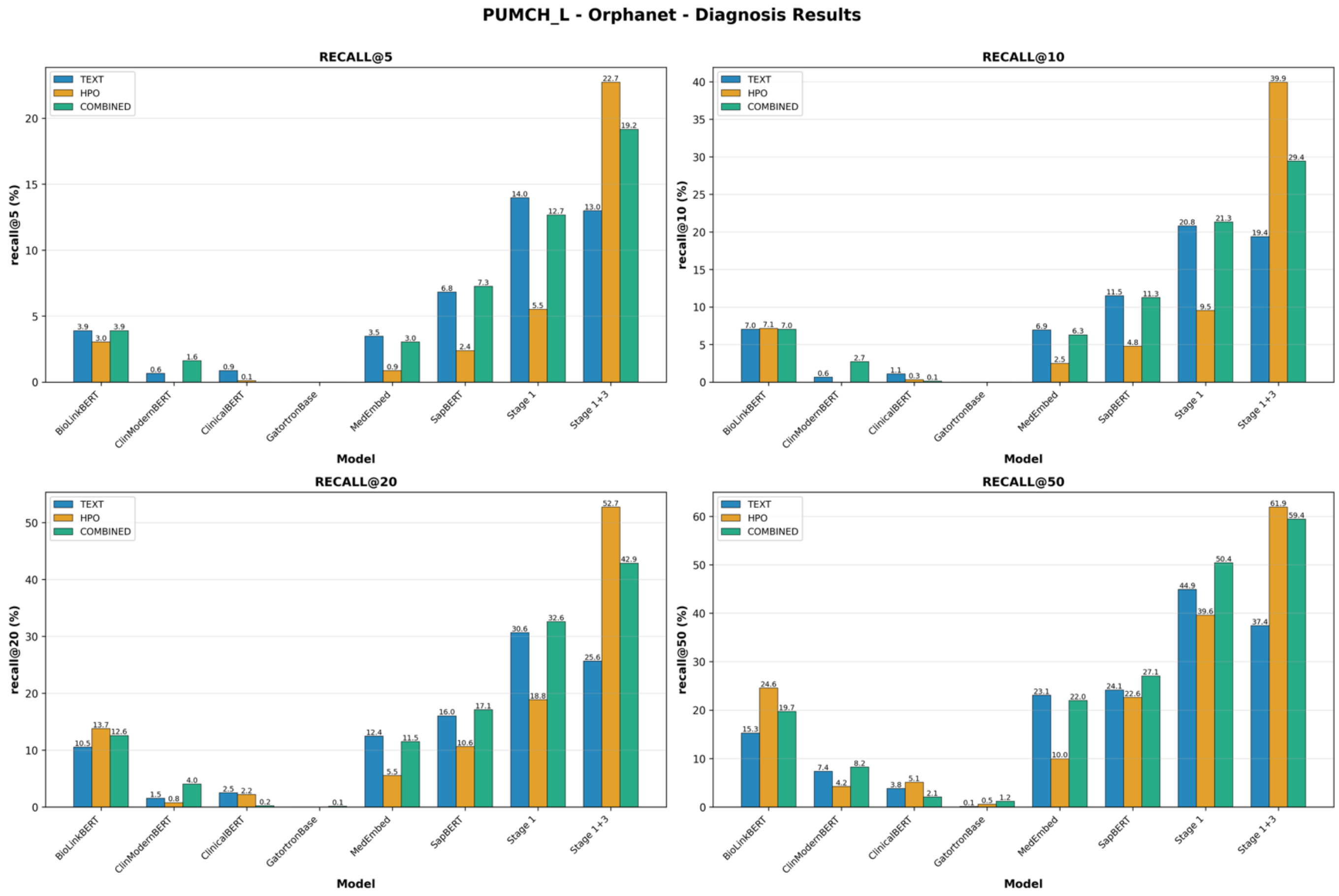


**Figure S19.** Orphanet-based disease ranking comparison across all modalities with a focus on the PUMCH-L corpus

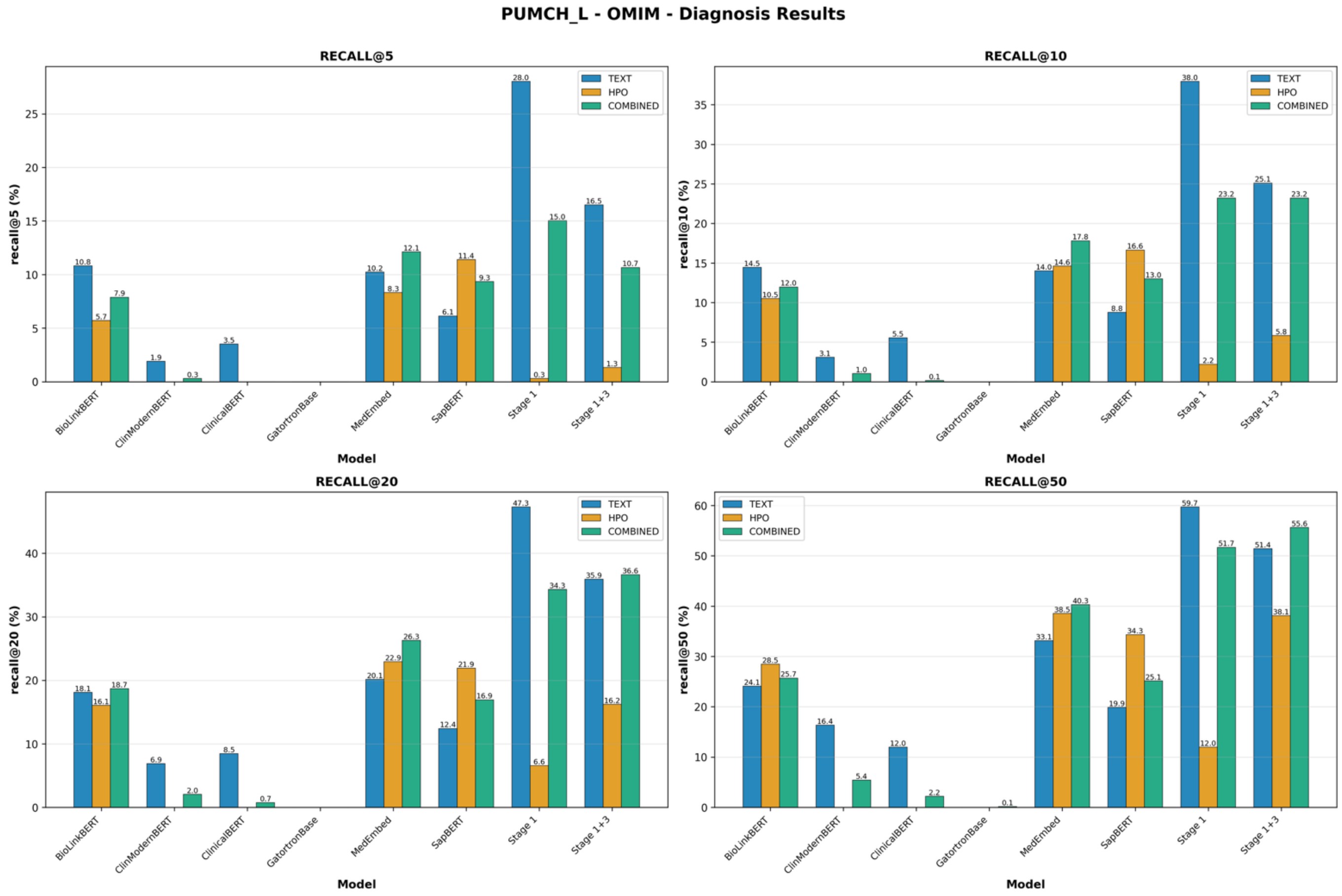


**Figure S20.** OMIM-based disease ranking comparison across all modalities with a focus on the PUMCH-L corpus

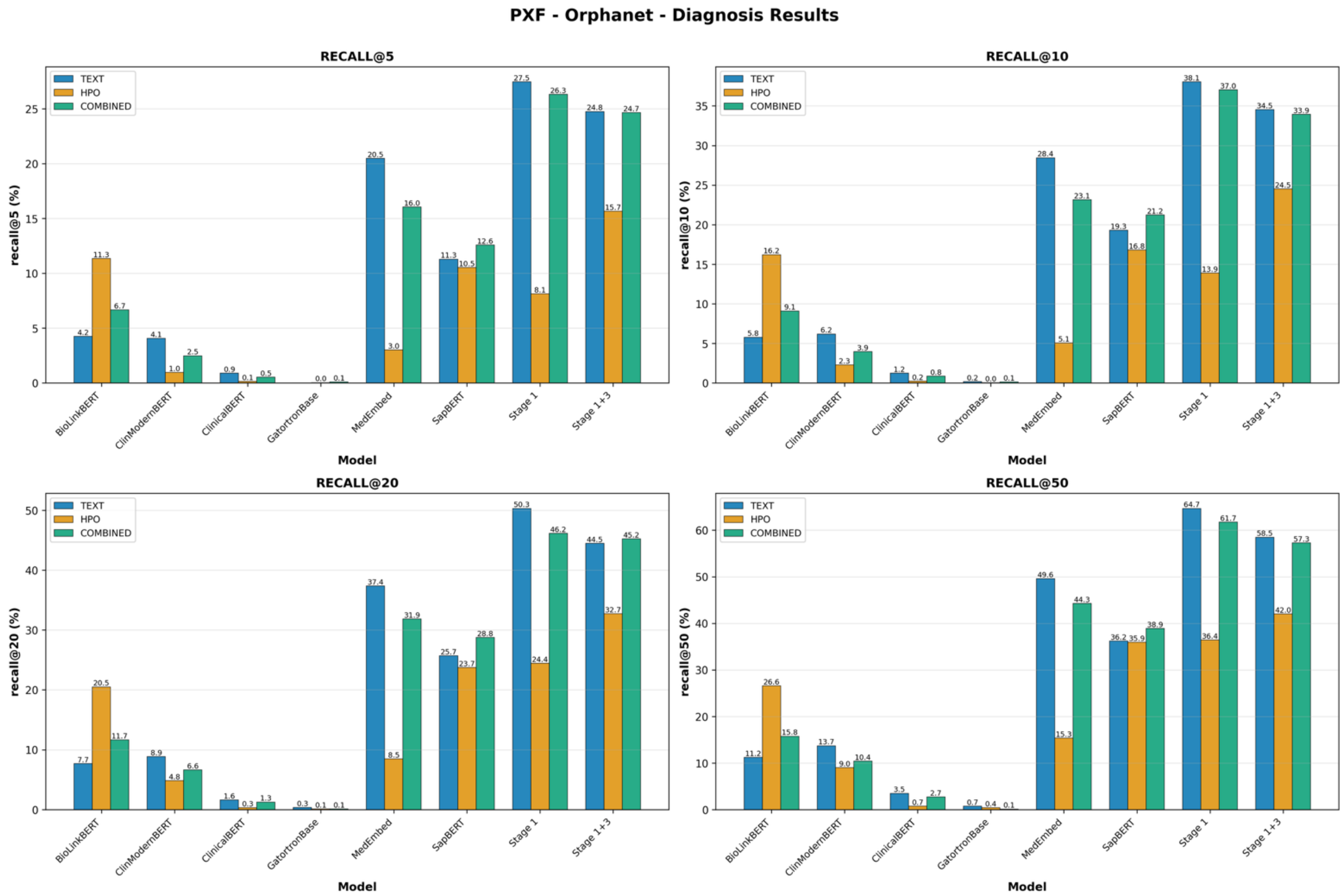


**Figure S21.** Orphanet-based disease ranking comparison across all modalities with a focus on the PXF corpus

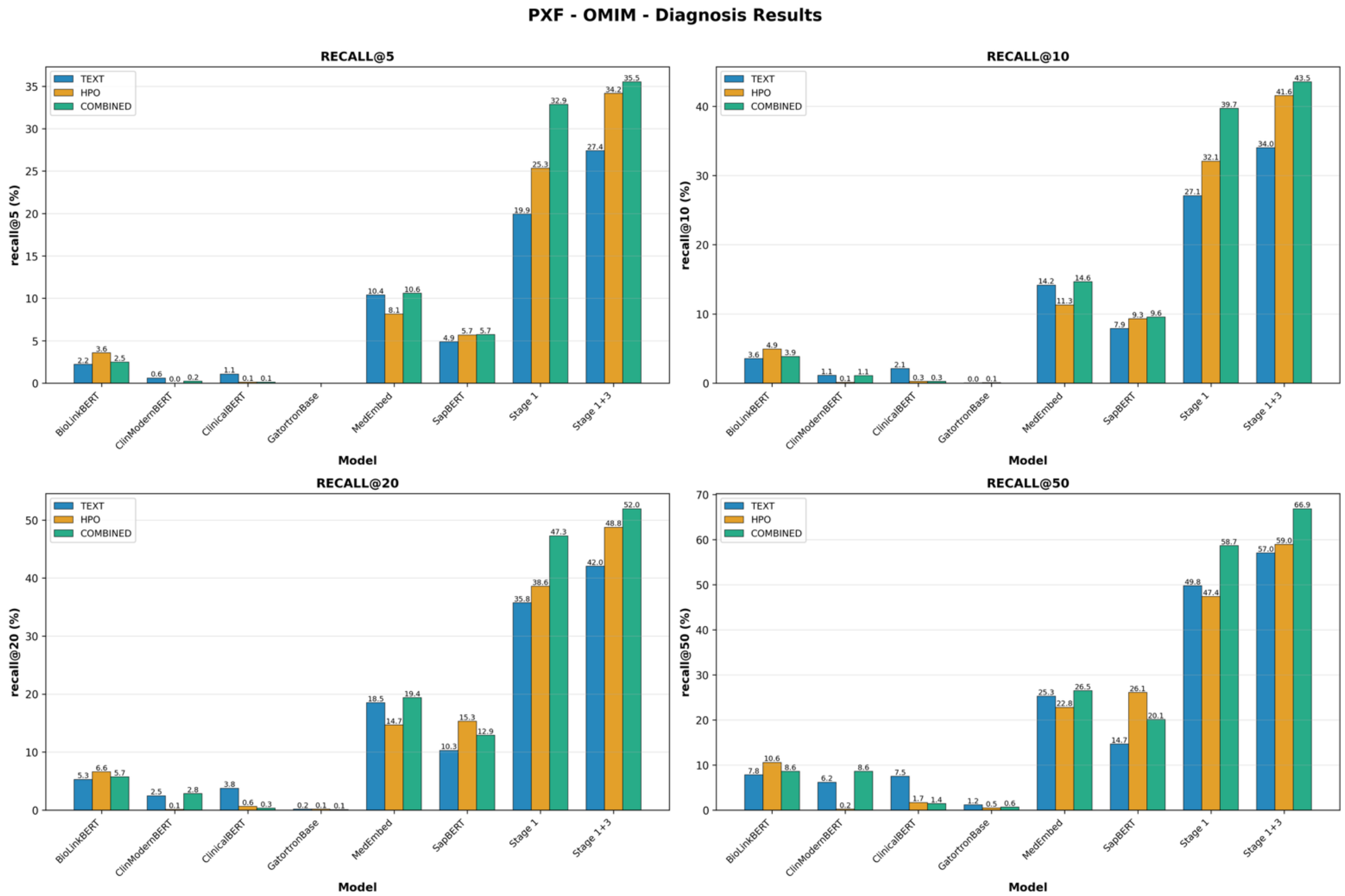


**Figure S22.** OMIM-based disease ranking comparison across all modalities with a focus on the PXF corpus

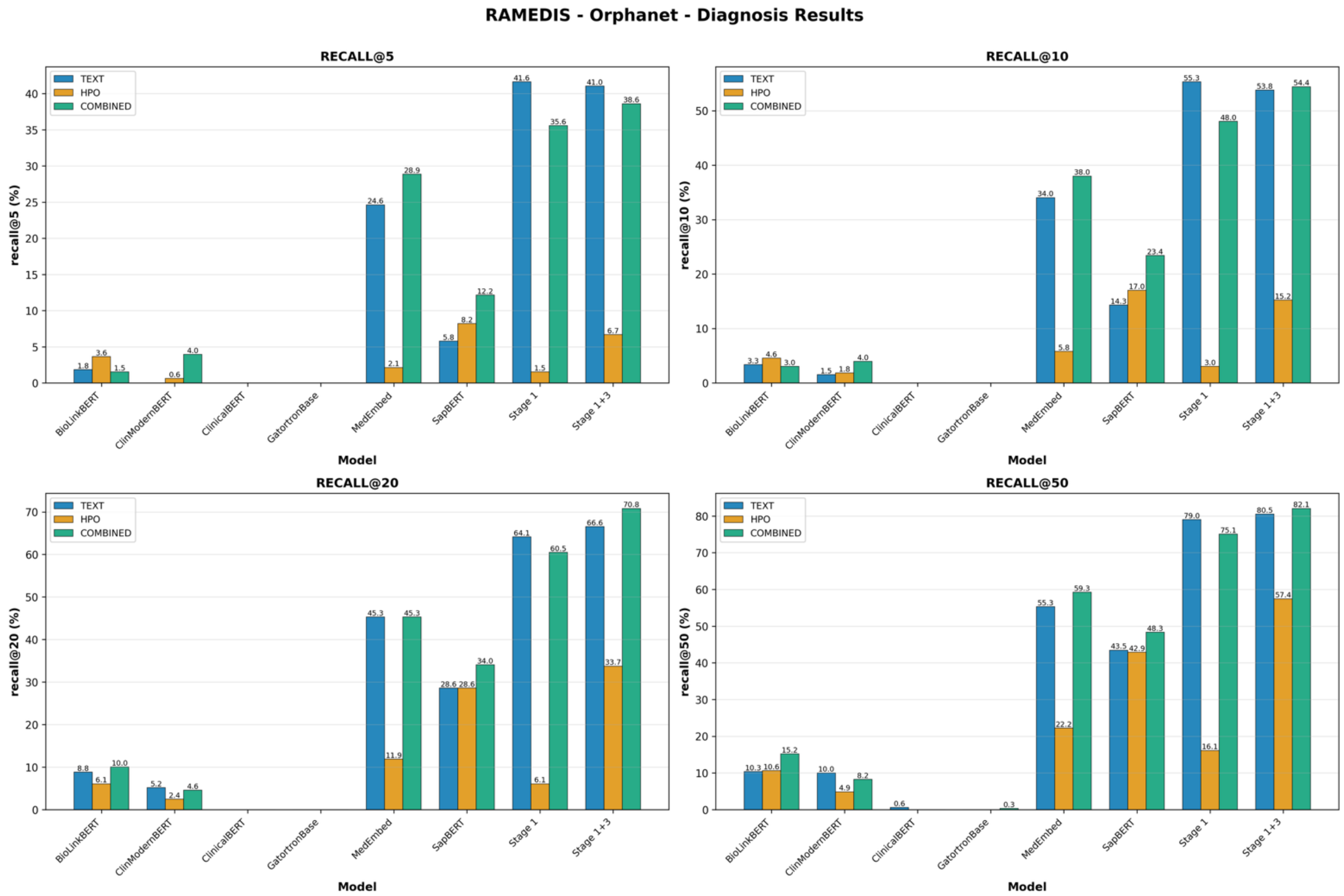


**Figure S23.** Orphanet-based disease ranking comparison across all modalities with a focus on the RAMEDIS corpus

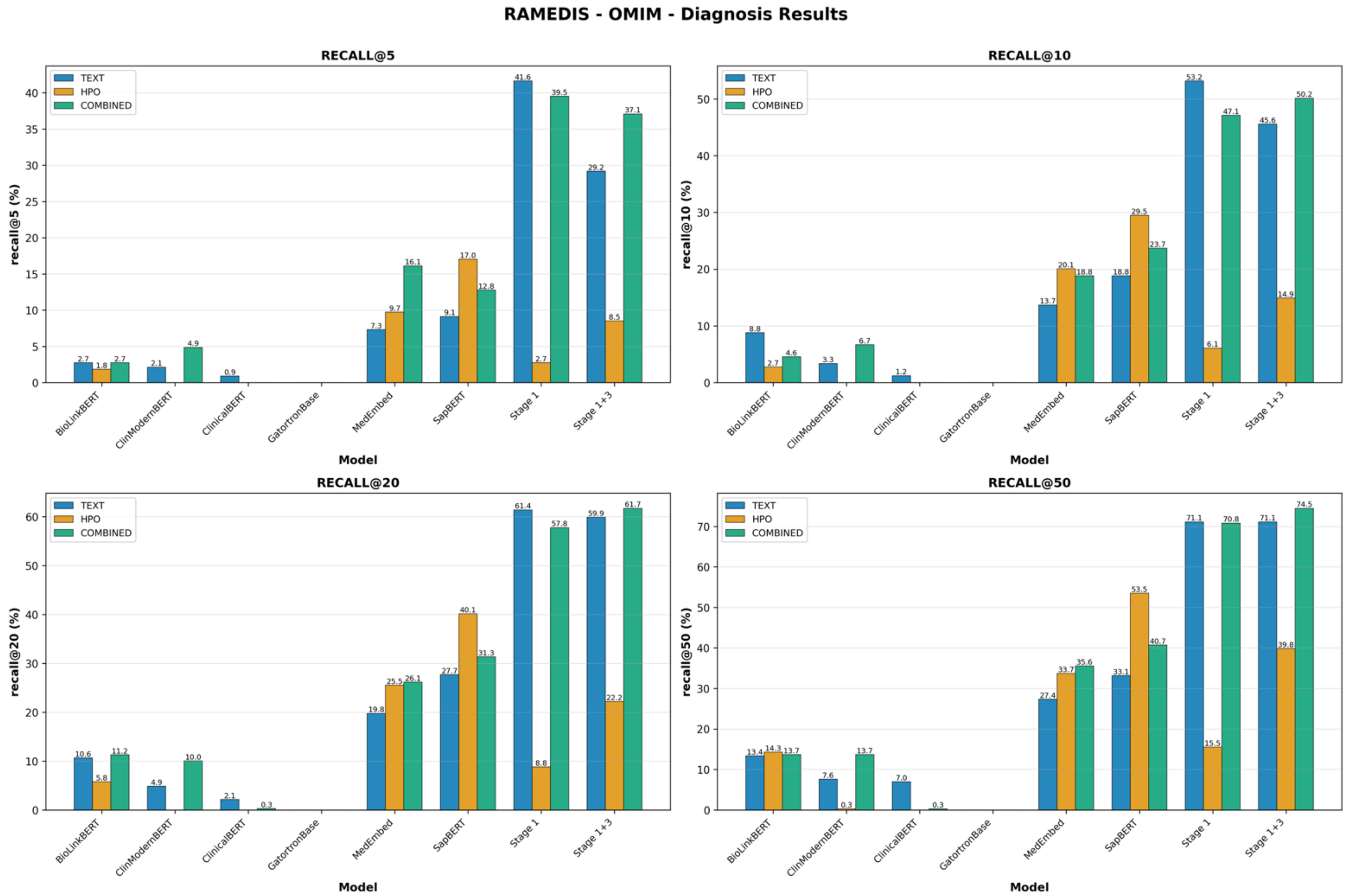


**Figure S24.** OMIM-based disease ranking comparison across all modalities with a focus on the RAMEDIS corpus

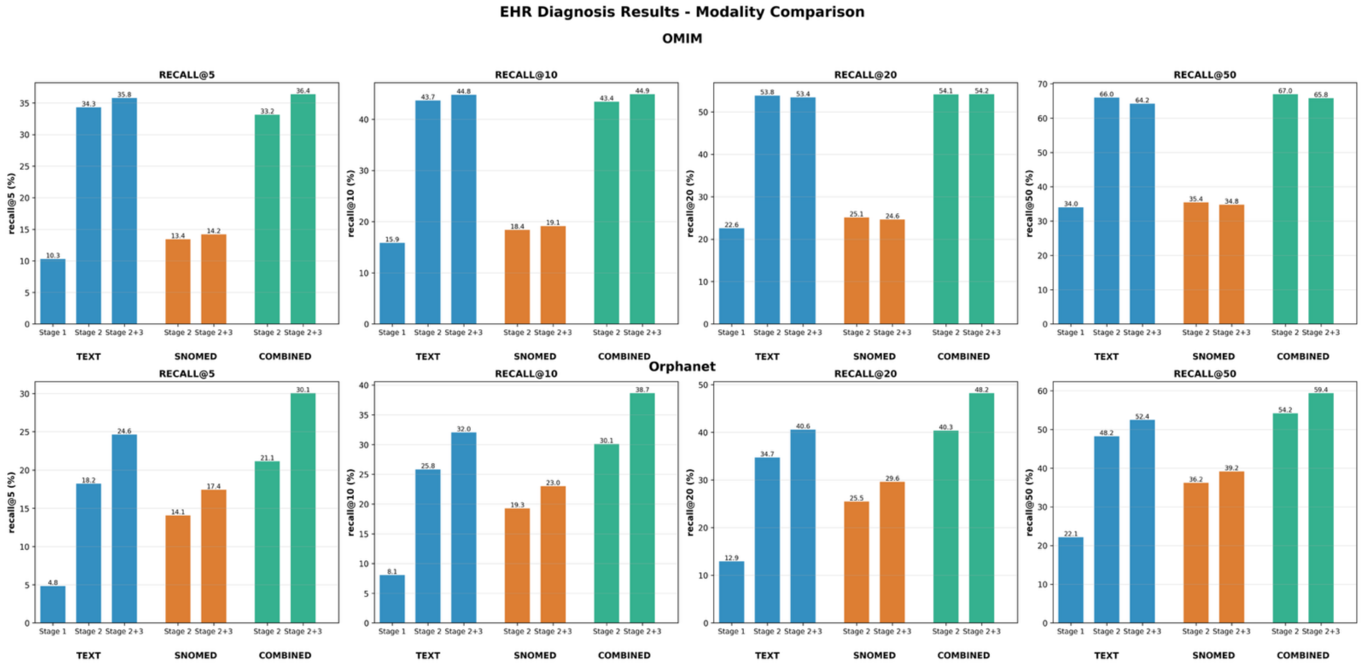


**Figure S25.** Orphanet and OMIM-based disease ranking comparison across all modalities with a focus on the EHR corpus

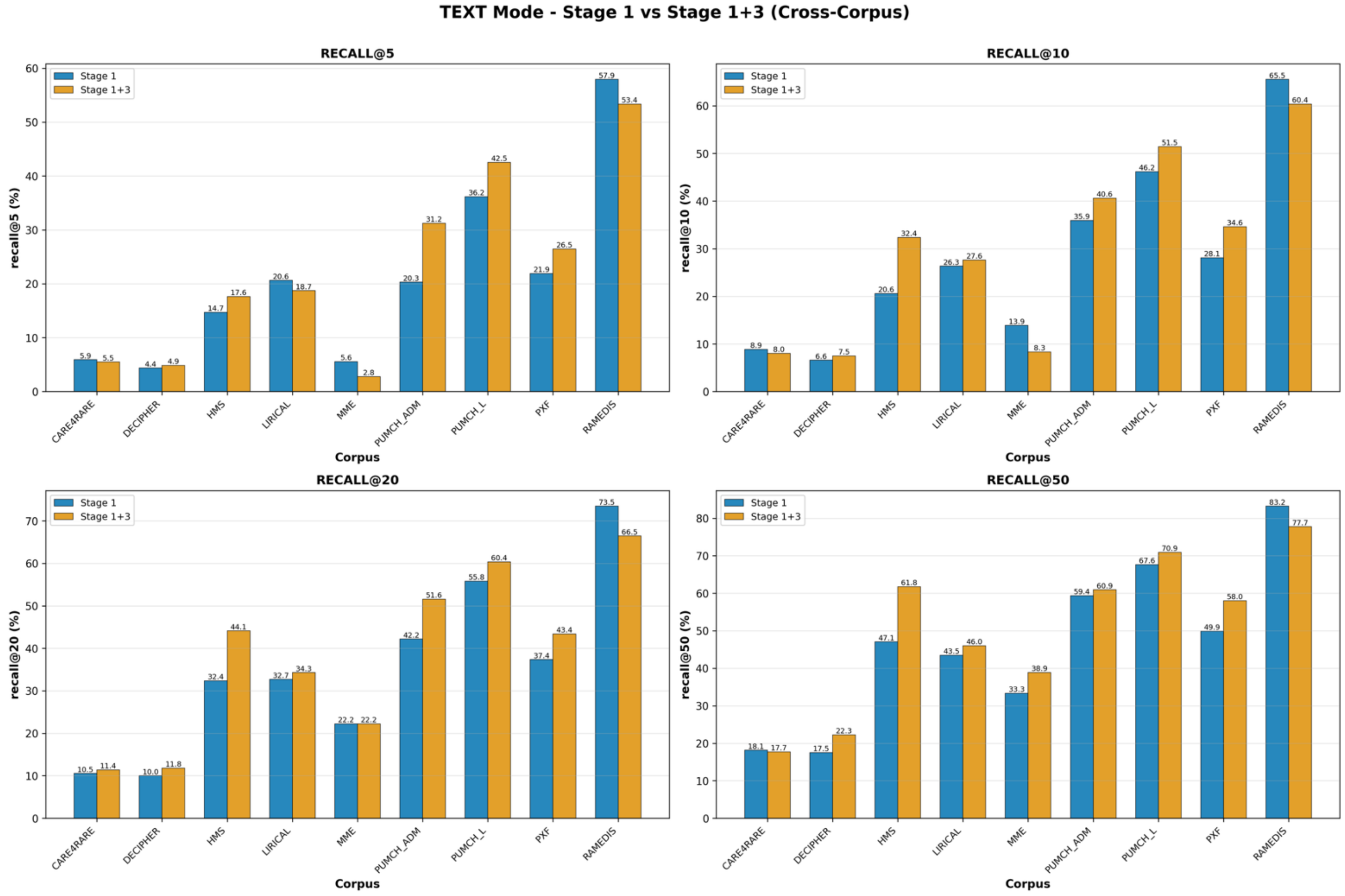


**Figure S26.** Gene identification, cross-corpus comparison using the TEXT modality

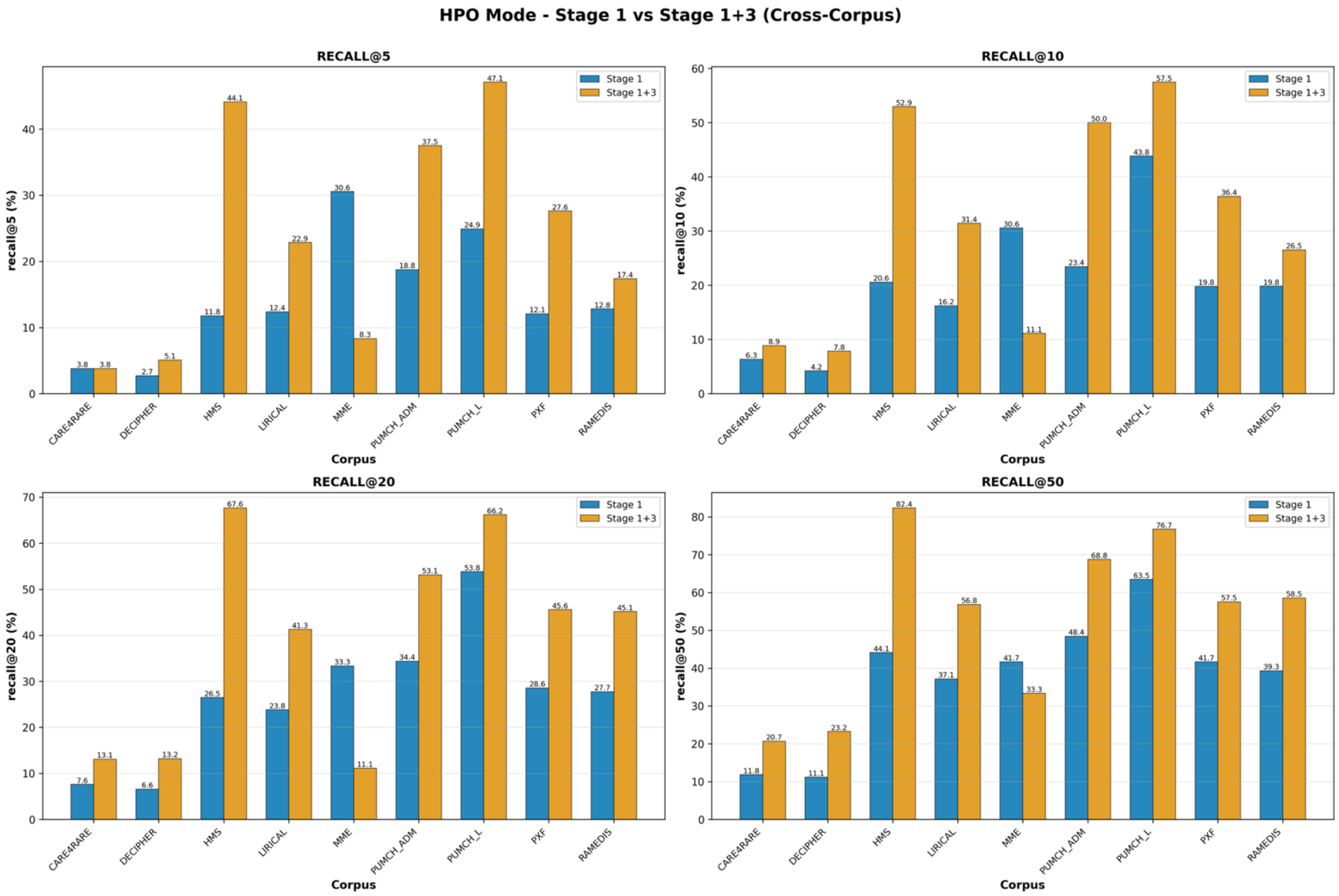


**Figure S27.** Gene identification, cross-corpus comparison using the HPO modality

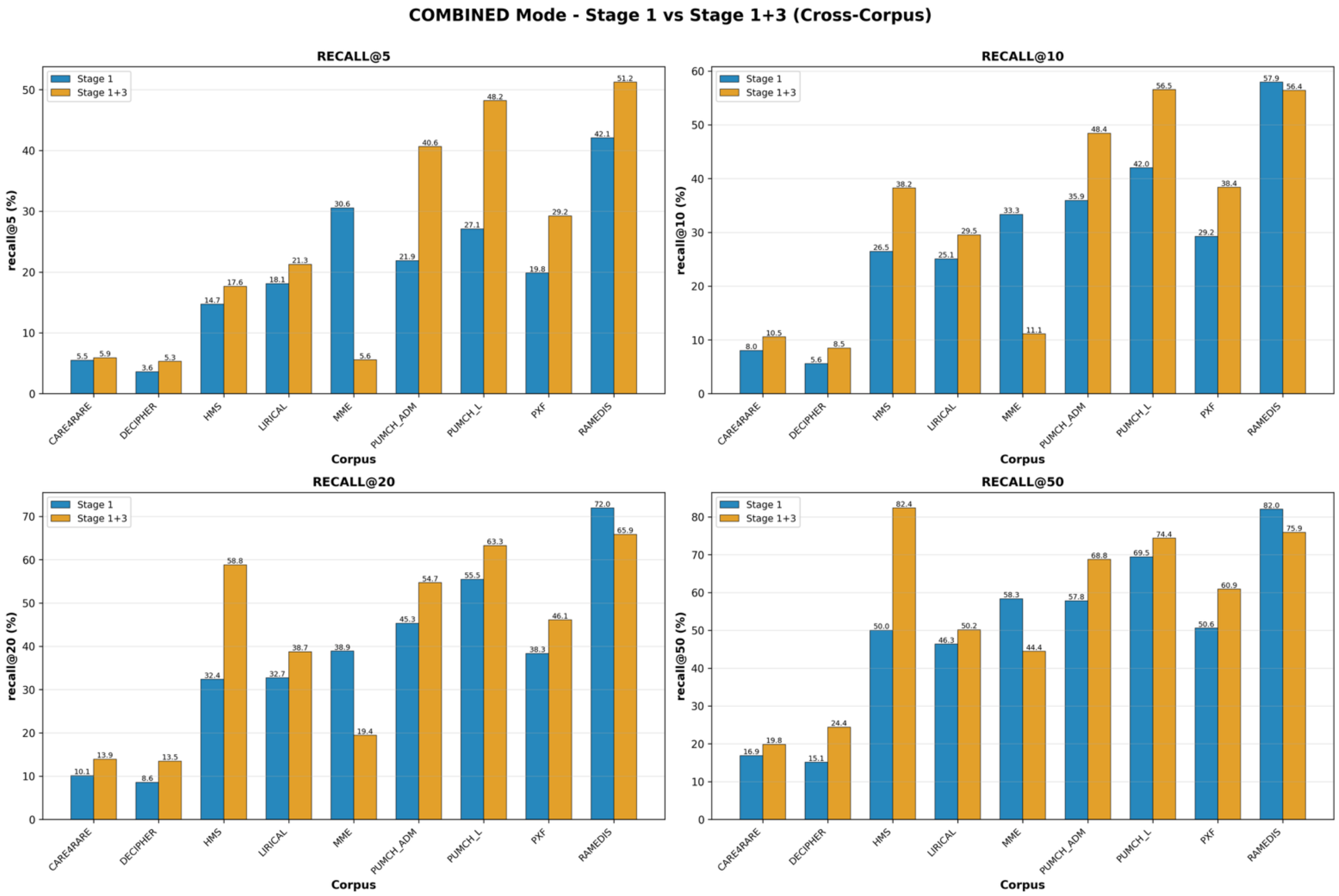


**Figure S28.** Gene identification, cross-corpus comparison using the COMBINED modality

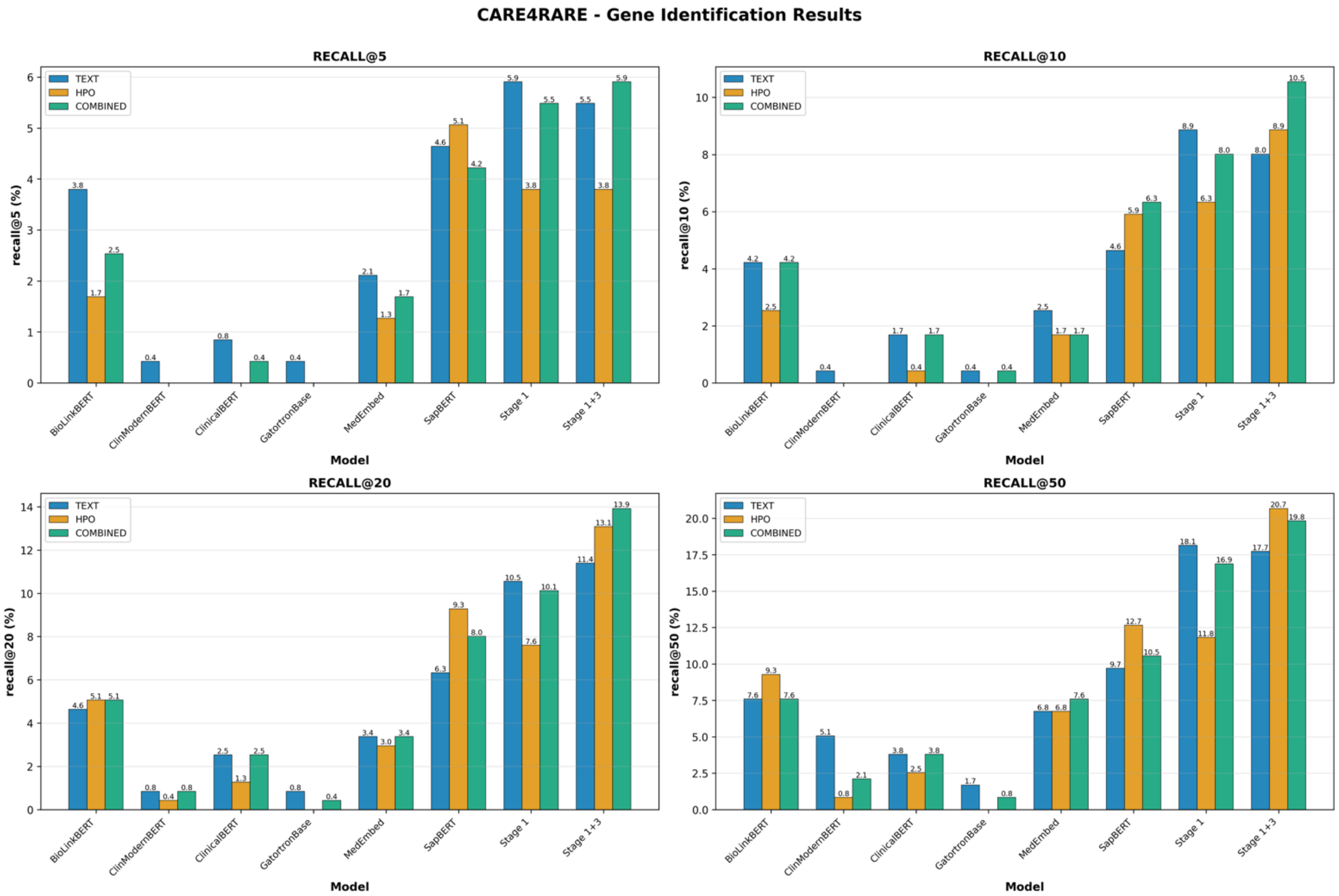


**Figure S29.** Gene identification comparison across all modalities with a focus on the Care4Rare corpus

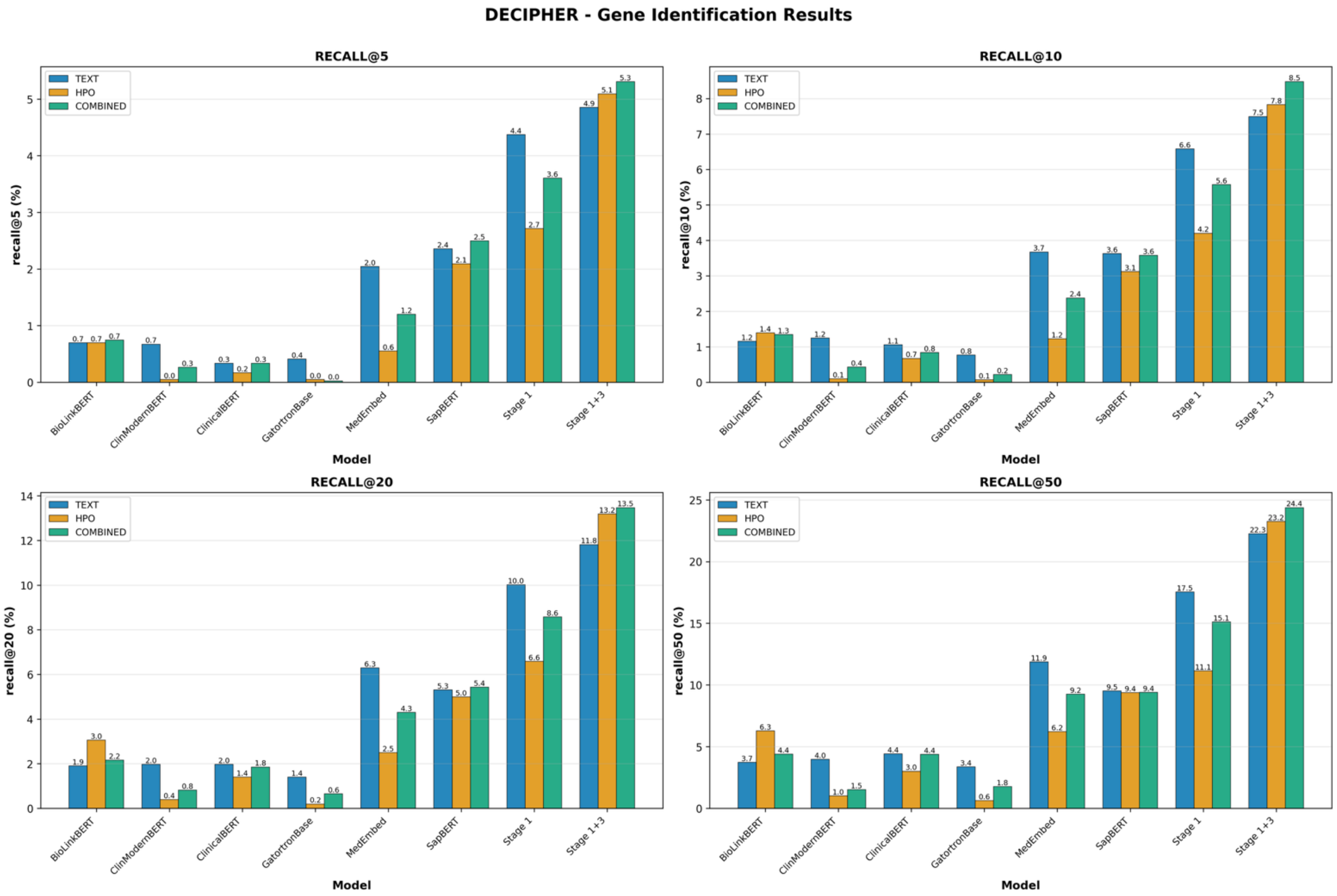


**Figure S30.** Gene identification comparison across all modalities with a focus on the DECIPHER corpus

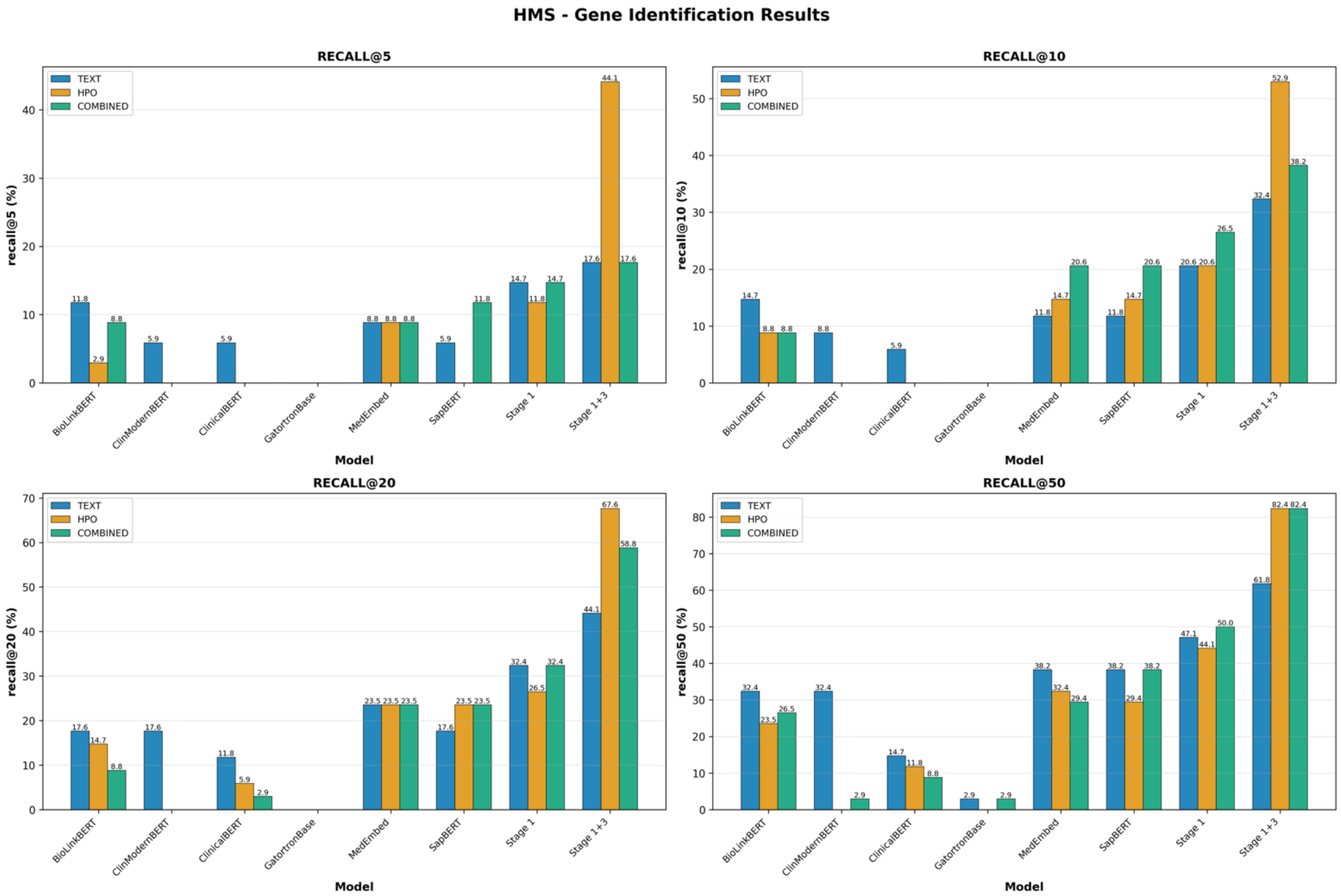


**Figure S31.** Gene identification comparison across all modalities with a focus on the HMS corpus

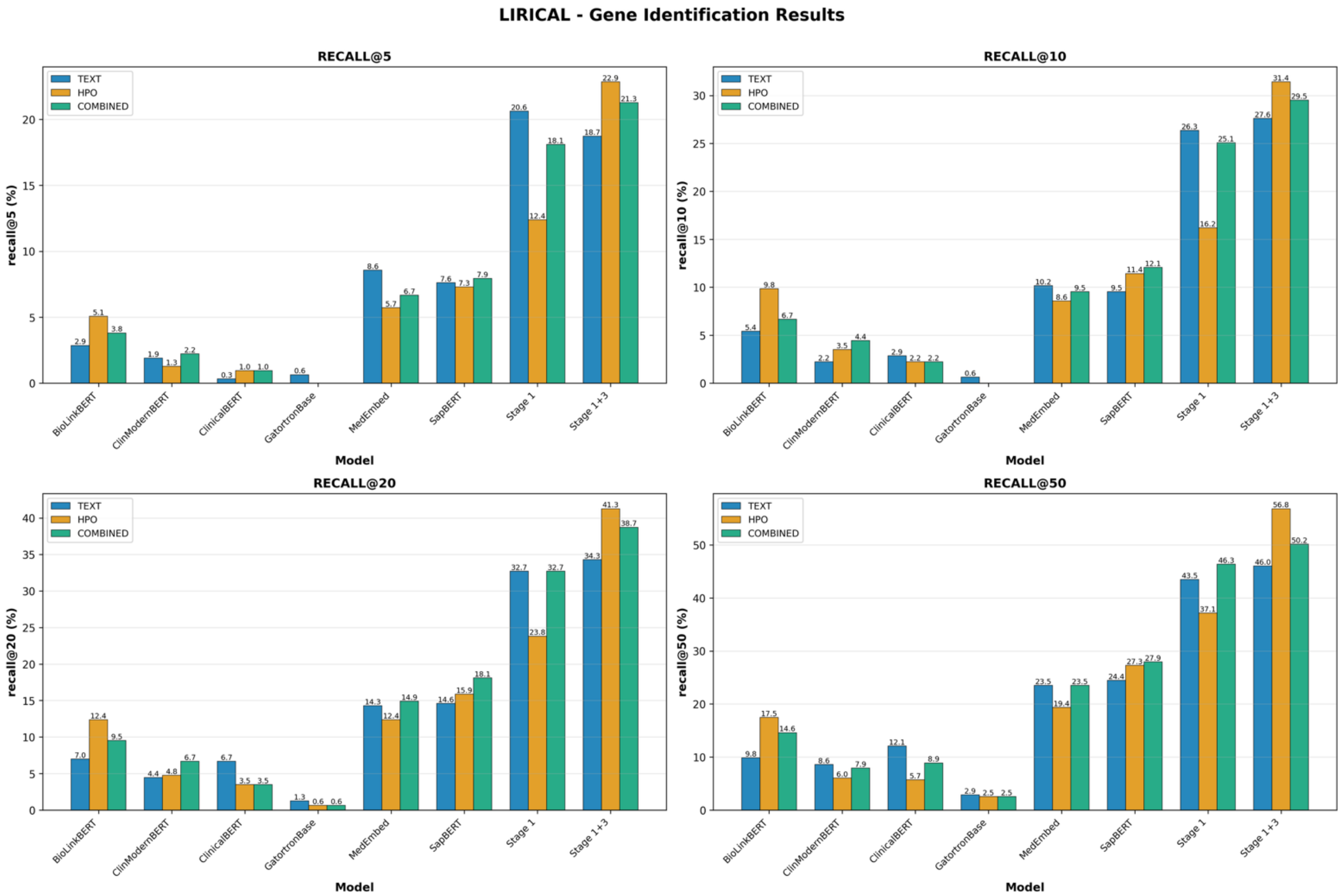


**Figure S32.** Gene identification comparison across all modalities with a focus on the LIRICAL corpus

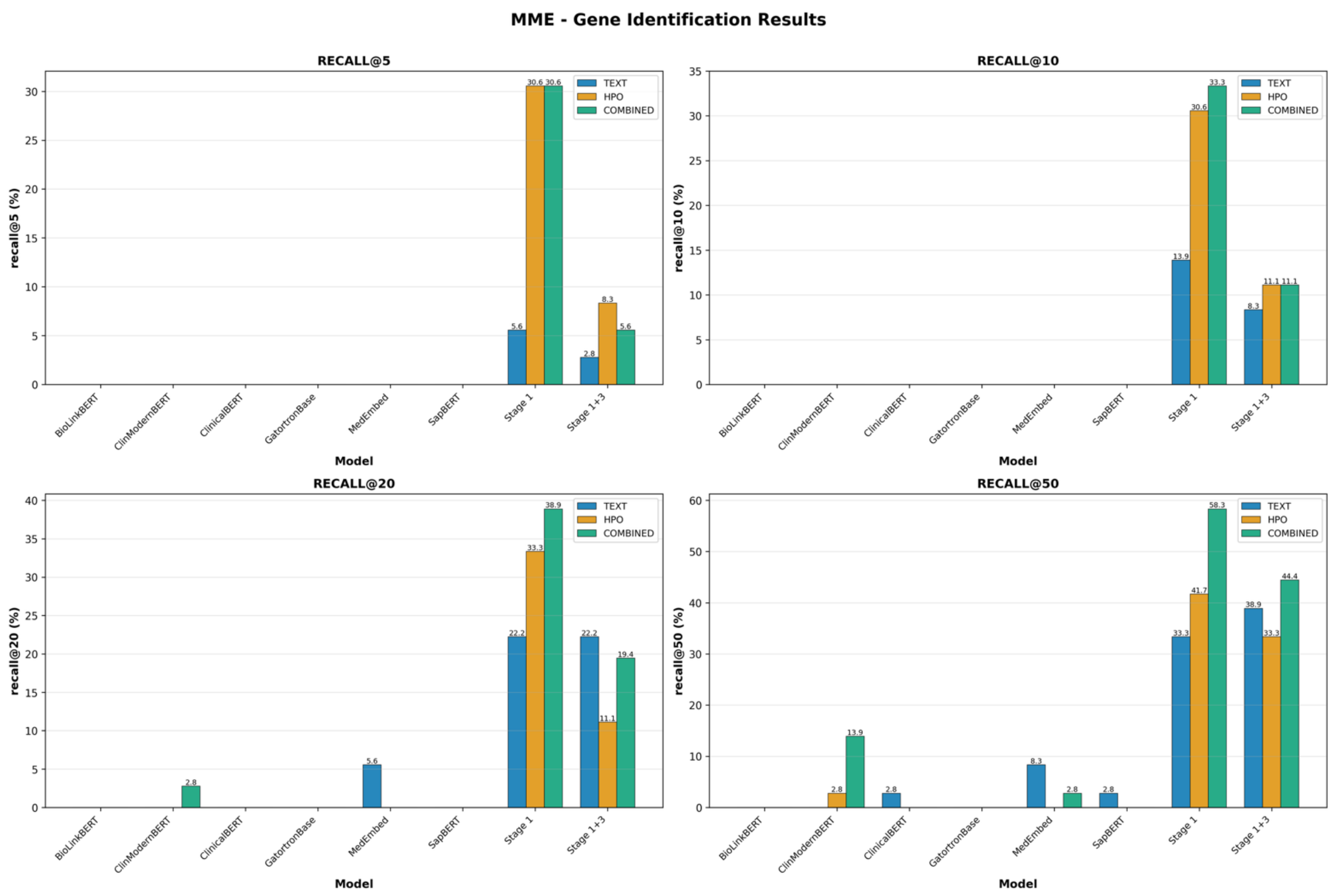

**Figure S33.** Gene identification comparison across all modalities with a focus on the MME corpus

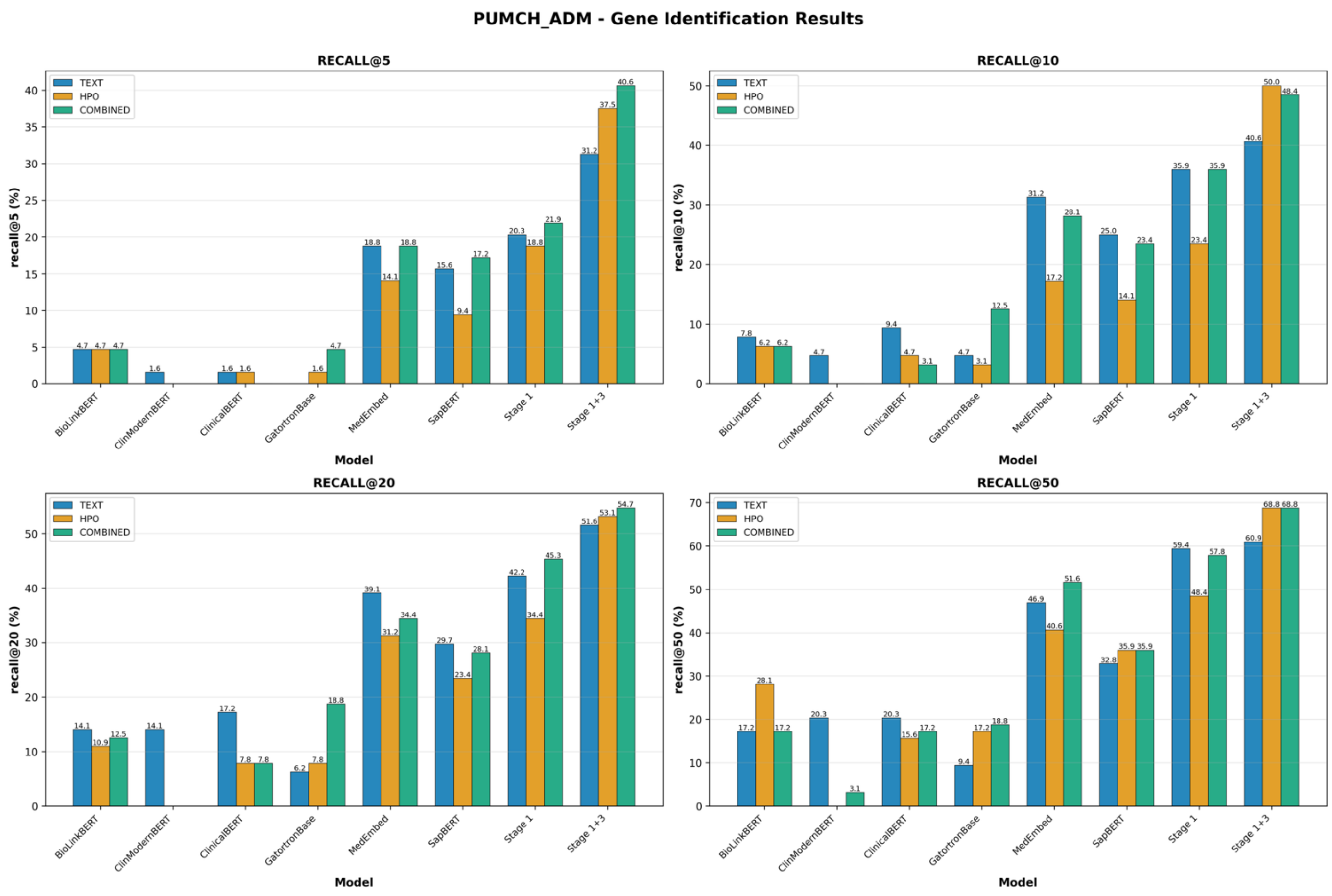

**Figure S34.** Gene identification comparison across all modalities with a focus on the PUMCH-ADM corpus

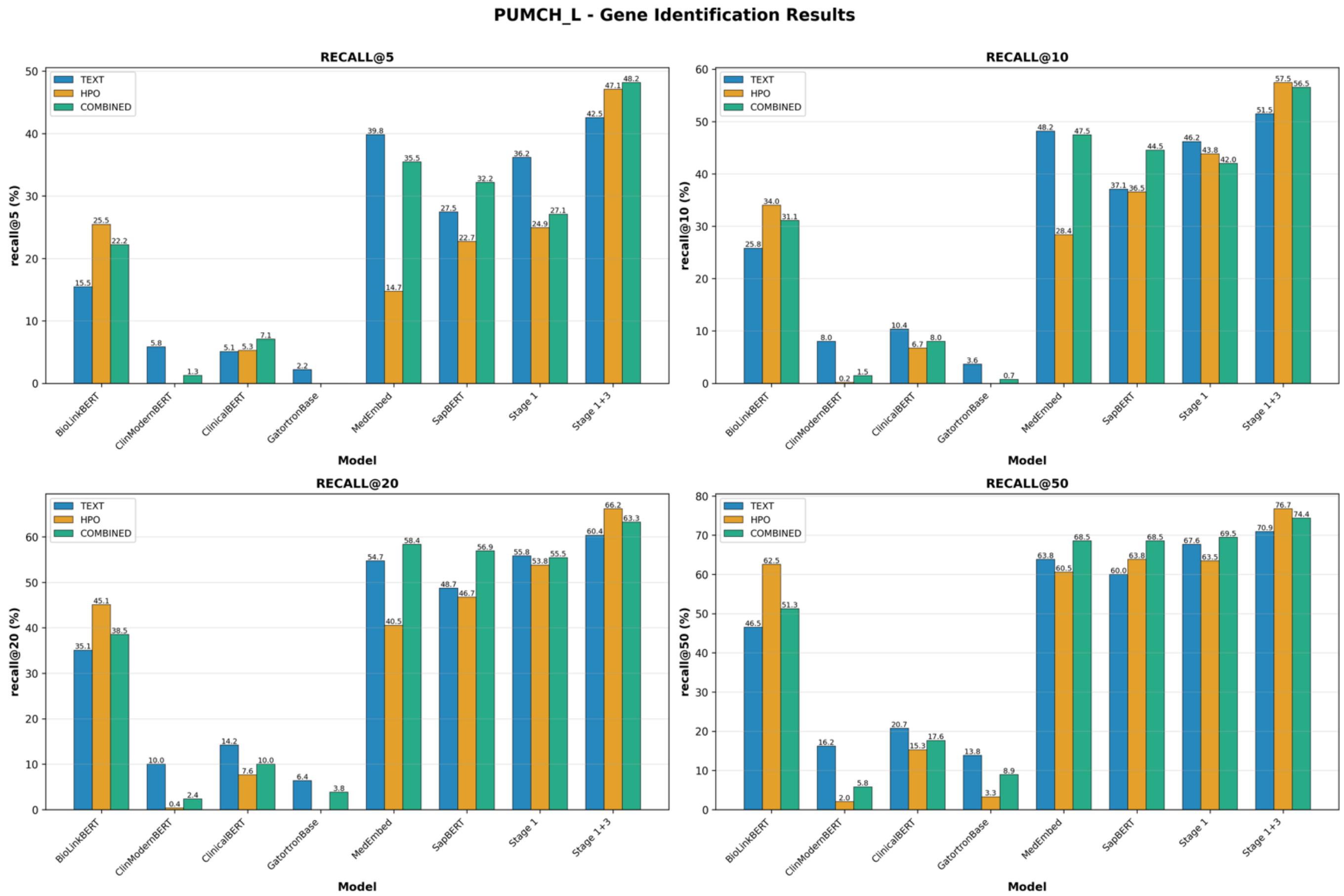


**Figure S35.** Gene identification comparison across all modalities with a focus on the PUMCH-L corpus

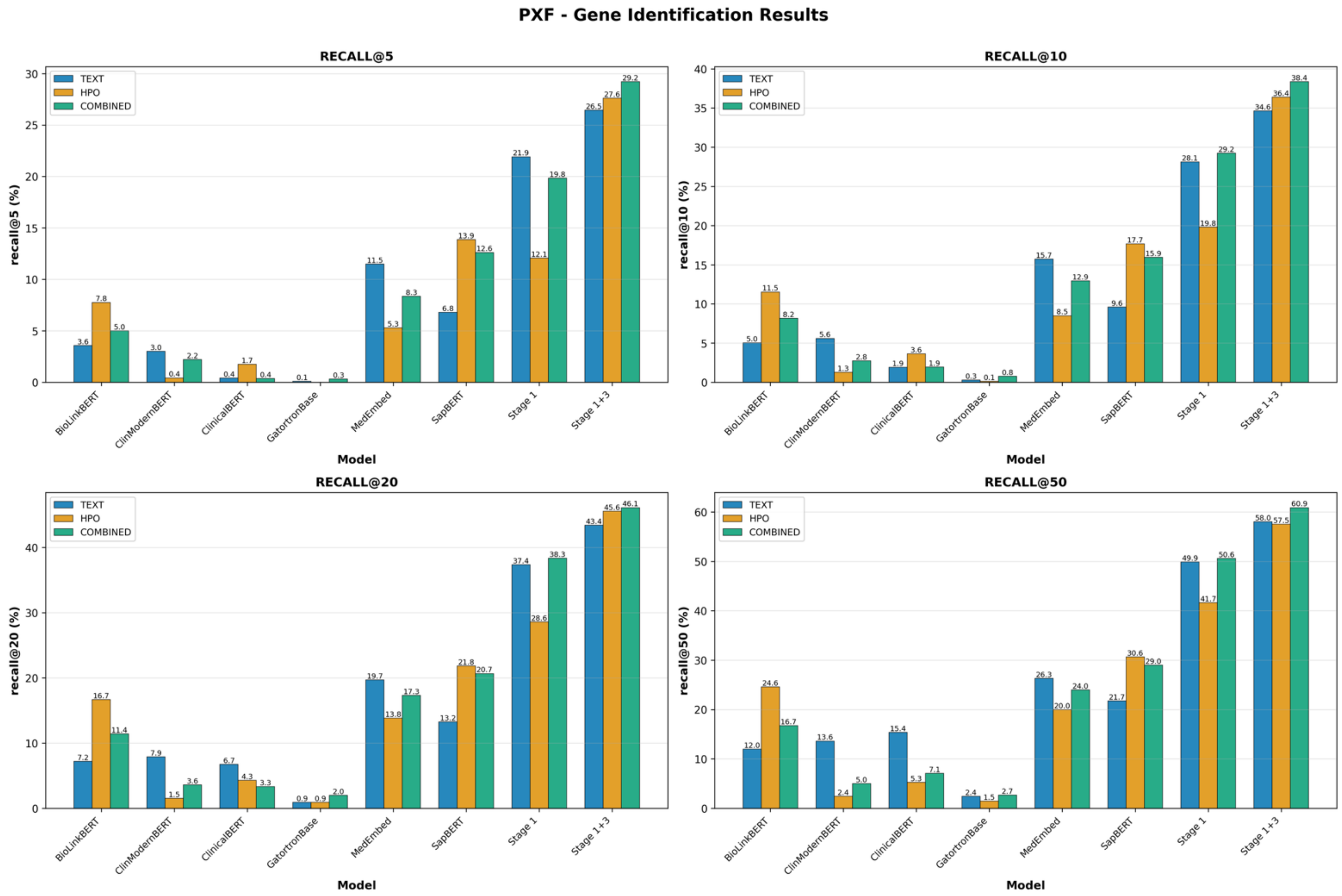


**Figure S36.** Gene identification comparison across all modalities with a focus on the PXF corpus

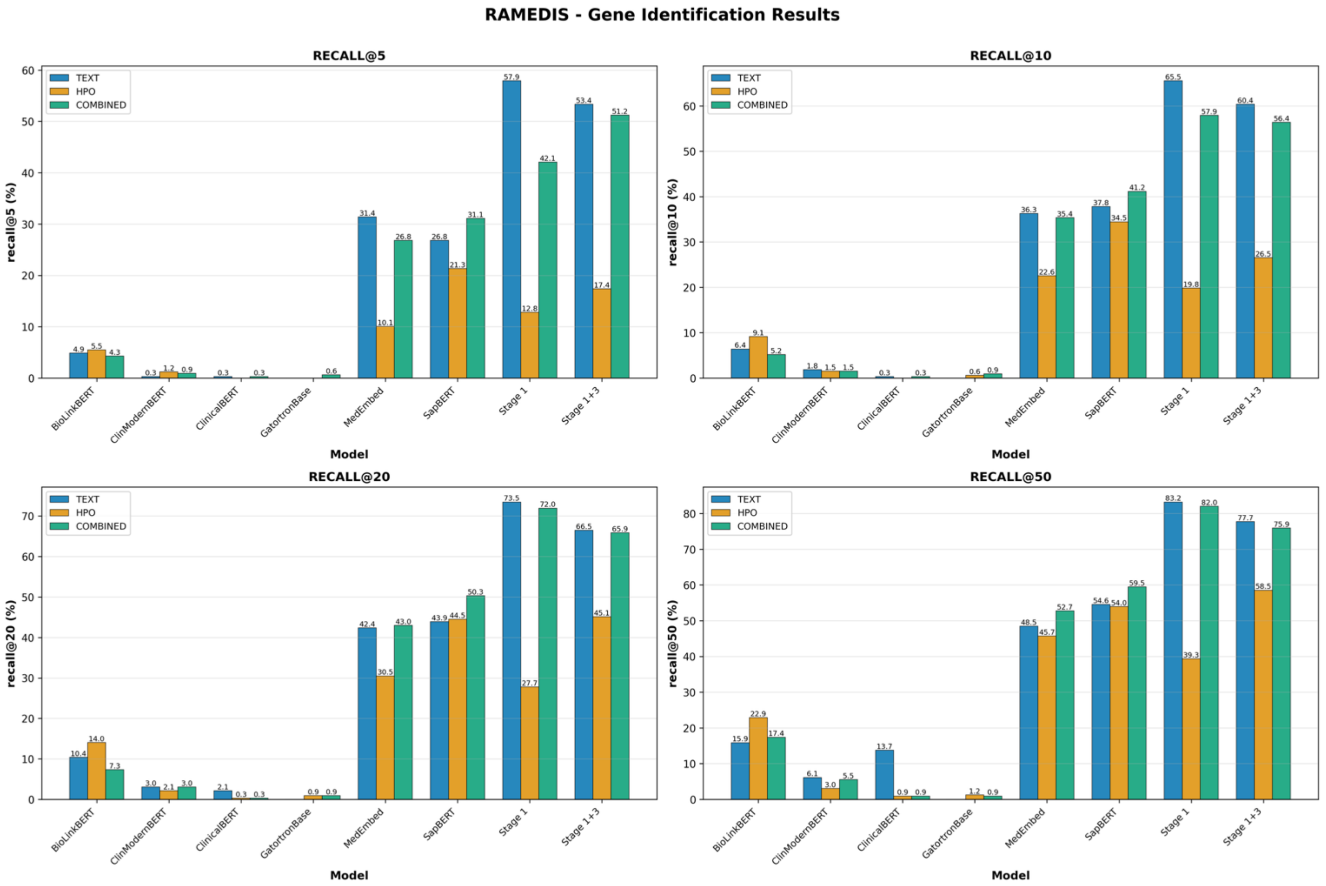


**Figure S37.** Gene identification comparison across all modalities with a focus on the RAMEDIS corpus

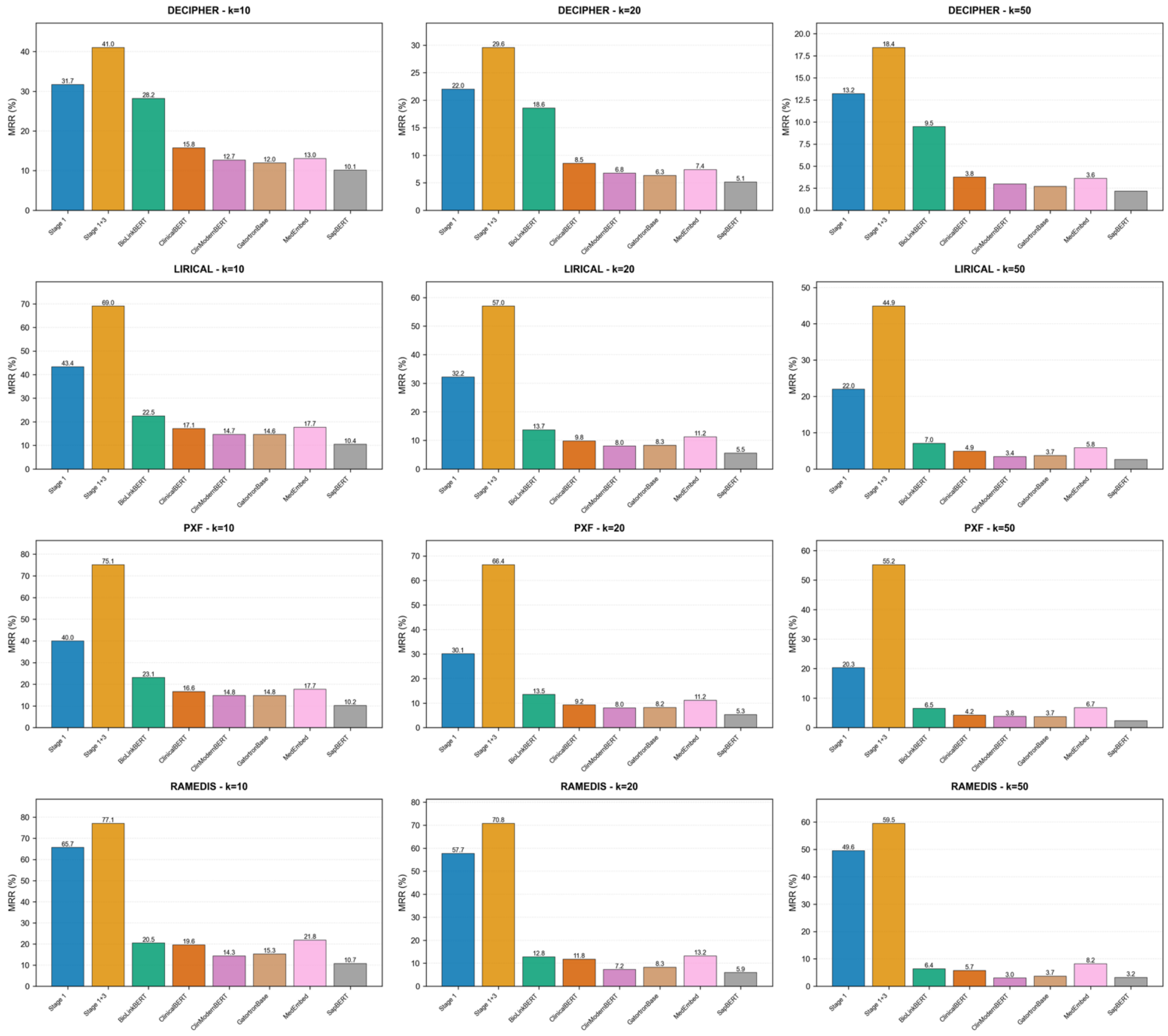


**Figure S38.** Differential ranking MRR results across all models and corpora at various k cut-offs

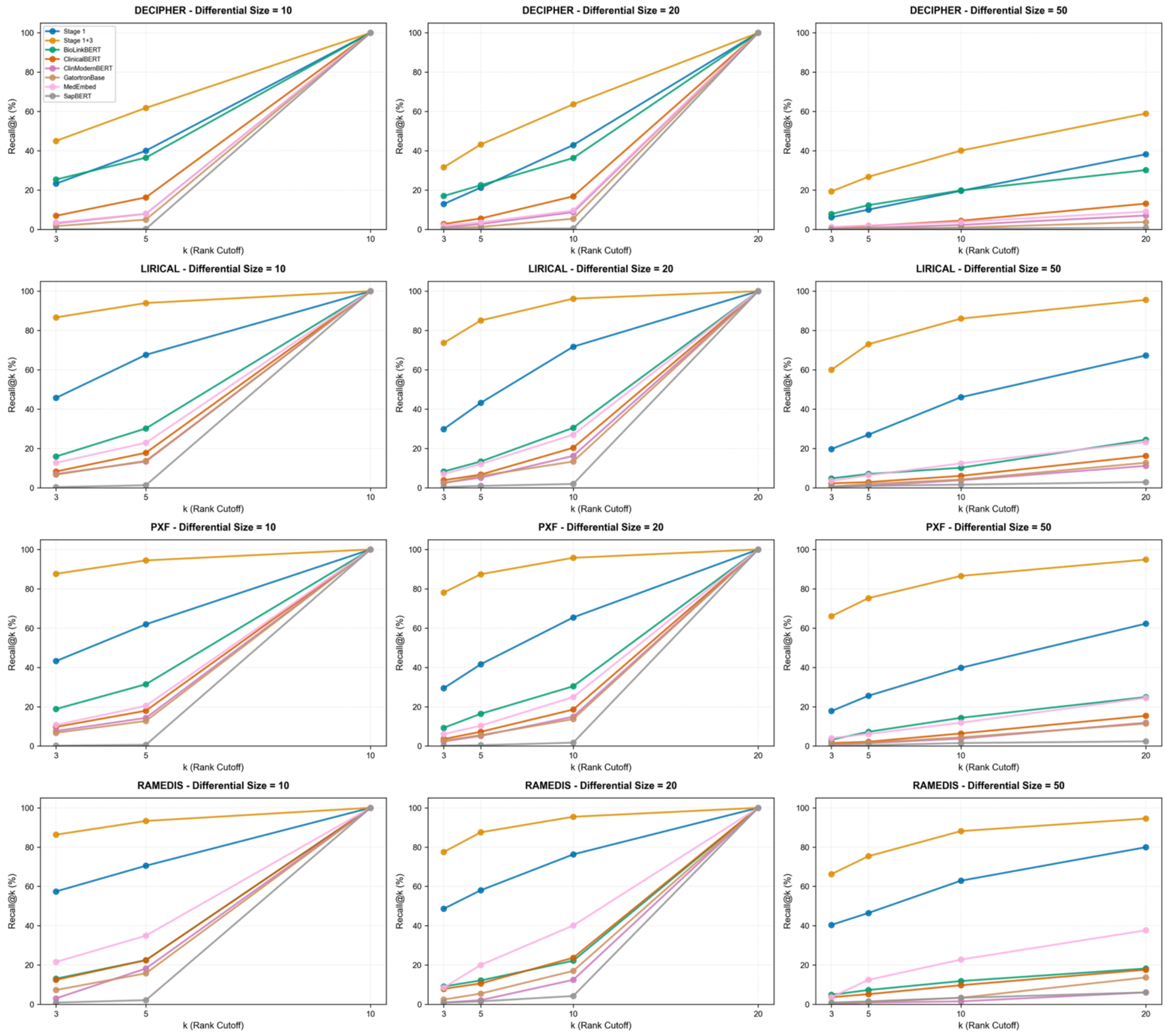


**Figure S39.** Differential ranking recall results across all models and corpora at various k cut-offs
